# Tuberculosis treatment outcome in Cameroon: a systematic review and meta-analysis

**DOI:** 10.64898/2026.11.07.26367494

**Authors:** Fabrice Zobel Lekeumo Cheuyem, Jude Tsafack Zefack, Rick Tchamani, Chabeja Achangwa, Clovis Josué Kachiwouo Mapouo, Armand Tiotsia Tsapi, Claude Axel Minkandi

## Abstract

**Background:** Tuberculosis (TB) remains a major public health challenge in Cameroon, yet national evidence on treatment outcomes across the full spectrum of routine care is limited. This systematic review and meta-analysis estimated pooled proportions of treatment success, mortality, treatment failure and loss to follow-up (LTFU) among TB patients in Cameroon, and identified predictors of unfavorable outcomes.

**Methods:** PubMed, Embase, Scopus, Web of Science, African Journals Online and the Cochrane Library were searched, supplemented by Google Scholar and reference-list screening. Observational studies conducted in Cameroon reporting at least one WHO-defined treatment outcome were eligible. Pooled proportions and adjusted odds ratios (AOR) were estimated with random-effects models meta-analysis. Data were analyzed using R Statistics version 4.6.1.

**Results:** Thirty-two studies (25 cohorts, 7 cross-sectional), conducted between 1998 and 2022 across six of Cameroon’s ten regions, were included. Pooled treatment success was 78.2% (95% CI: 73.8–82.1; 27 reports; 71,784 participants; *I²* = 98.2%), mortality 8.1% (95% CI: 5.5–11.7; 25 reports; 71,870 participants; *I²* = 97.8%), treatment failure 0.9% (95% CI: 0.5–1.4; 26 reports; 71,389 participants; *I²* = 89.1%) and LTFU 6.8% (95% CI: 5.0–9.1; 26 reports; 70,768 participants; *I²* = 98.6%). Mortality was highest in the South-West (16.2%) and among TB/HIV-coinfected patients (30.5%), and lowest among children (5.0%). Treatment success was lowest in the Littoral (71.1%) and among TB/HIV-coinfected patients (60.8%) and highest among multidrug-resistant TB (89.3%) and pediatric patients (86.8%). Treatment failure significantly declined from 2.9% before 2005 to 0.5% from 2020 onward (*p* < 0.001). Significant predictors of unfavorable outcomes were HIV positivity (AOR = 2.57; 95% CI: 1.86–3.55), previous TB treatment (AOR = 1.99; 95% CI: 1.40–2.84), male sex (AOR = 1.67; 95% CI: 1.45–1.92), age ≤40 years (AOR = 1.45; 95% CI: 1.06–1.98) and absence of antiretroviral therapy (AOR = 1.71; 95% CI: 1.15–2.54).

**Conclusions:** TB treatment outcomes in Cameroon remain suboptimal, with nearly one in five patients not achieving treatment success and high mortality among TB/HIV-coinfected individuals. Strengthening TB/HIV service integration, early antiretroviral therapy initiation, targeted support for previously treated patients and men, and improved retention in care are needed.

**Systematic review registration number:** CRD420261404490

## 1. Background

Tuberculosis (TB) remains one of the world’s leading infectious causes of illness and death, despite being preventable and curable when timely diagnosis and effective treatment are available. In 2024, tuberculosis was estimated to cause 10.7 million cases and 1.23 million deaths, including 150,000 among people living with HIV. Only 40% of patients with drug-resistant TB received treatment, even though major advances since 2000 have saved 83 million lives [1]. The burden is distributed unevenly, with low- and middle-income countries accounting for most cases and deaths. Sub-Saharan Africa continues to experience a particularly high burden because TB transmission and outcomes are shaped by delayed diagnosis, poverty, undernutrition, overcrowding, HIV infection, limited access to rapid diagnostics, and weaknesses in health-service delivery. Despite regional and global advancements, the rate of decline is still too slow to meet the objectives of the Sustainable Development Goals relating to tuberculosis and the World Health Organization’s (WHO) End TB Strategy [2, 3].

Cameroon remains an important setting for TB control in Central Africa. Available estimates indicate that TB incidence remained approximately 132 cases per 100,000 population in 2024, demonstrating continued transmission and a substantial need for diagnosis, treatment, and follow-up services [4]. The country’s response is implemented through the National Tuberculosis Control Programme and a network of diagnostic and treatment centres [2]. However, treatment coverage, equitable access to diagnostic services, continuity of care, and detection of drug-resistant disease remain important programmatic concerns [5]. Additionally, Cameroon’s 10 regions differ significantly in terms of geography, socioeconomic status, and health systems. As a result, significant differences between urban referral hospitals and decentralized treatment facilities, as well as between areas impacted differently by poverty, population mobility, insecurity, a lack of medical professionals, and disruptions in access to medications and laboratory services, may be hidden by national aggregate indicators.

Treatment-outcome monitoring is a core component of TB surveillance because it indicates whether patients who begin therapy are retained in care and achieve favorable clinical results. WHO reporting frameworks classify treatment outcomes using standardized categories, including cured, treatment completed, treatment failed, died, lost to follow-up, not evaluated, and treatment success, the latter generally combining cure and treatment completion [6, 7]. These definitions facilitate comparison across programmes and periods.

Treatment for tuberculosis that is successful lowers the chance of developing drug resistance as well as morbidity, mortality, recurrence, and continued transmission. On the other hand, the severity of the illness, delayed diagnosis, insufficient regimens, poor adherence, medication resistance, severe drug responses, or obstacles to continuity of care may be reflected in death, treatment failure, and loss to follow-up [7]. Age, socioeconomic position, prior TB treatment, HIV status, comorbidities, treatment support, and health-system capability all have an impact on these treatment outcomes [8].

The clinical form of TB is another potential source of outcome variation. Pulmonary TB (PTB), which affects the lungs, is the principal transmissible form and is usually evaluated through bacteriological examination of respiratory specimens, supported by clinical and radiological findings [9]. Extrapulmonary TB (EPTB) involves organs other than the lungs and may affect lymph nodes, pleura, bones and joints, the abdomen, genitourinary tract, or central nervous system. EPTB is often more difficult to diagnose because symptoms may be nonspecific and confirmation may require imaging, tissue biopsy, histopathology, or testing of paucibacillary non-respiratory specimens [10]. Evidence from Cameroon’s Littoral Region has shown that bacteriologically confirmed EPTB presents diagnostic and therapeutic challenges under routine programme conditions [11].

HIV infection is particularly relevant to TB presentation and prognosis in Cameroon. Immunosuppression increases the risk of progression from infection to active disease and is associated with disseminated or extrapulmonary forms, diagnostic difficulty, and increased mortality. Outcomes among people with TB/HIV coinfection may also depend on the timing and continuity of antiretroviral therapy, degree of immunosuppression, opportunistic infections, drug interactions, pill burden, and retention in care. A study in the Fako Division of Cameroon identified variation in TB treatment outcomes among people living with HIV and highlighted the importance of patient and health-service factors [12].

Drug-resistant TB further complicates treatment and threatens progress toward national and global targets [13, 14]. Multidrug-resistant TB (MDR-TB) is defined as disease caused by Mycobacterium tuberculosis resistant to at least isoniazid and rifampicin-resistant TB includes any rifampicin resistance, with or without resistance to other medicines [13]. MDR/Rifampicin resistant (RR)-TB has historically required longer, more toxic, and more expensive regimens than drug-susceptible TB, although WHO now prioritizes shorter all-oral regimens for eligible patients. Nevertheless, drug-resistant disease remains associated with adverse drug effects, demanding monitoring requirements, and a greater risk of unfavorable outcomes. An individual patient-data meta-analysis of 9,153 patients demonstrated the complexity and generally poorer outcomes of MDR-TB treatment, [15] while a comparative cohort study found a markedly higher risk of death among patients with MDR-TB than among those with drug-susceptible TB [16].

Cameroon-specific evidence similarly demonstrates that outcomes among patients with MDR/RR-TB are influenced by both clinical and programme factors. A study of rifampicin-resistant pulmonary TB in Yaoundé evaluated unfavorable outcomes among patients treated with a standardized shorter regimen and identified locally relevant predictors of poor outcome [17]. More recently, a Cameroon-focused systematic review and meta-analysis synthesized drug-resistant TB studies and reported pooled estimates for treatment success, death, loss to follow-up, and treatment failure [14]. That review provides an important synthesis of drug-resistant disease; however, it does not replace the need for a broader review that incorporates drug-susceptible TB and directly examines pulmonary and extrapulmonary disease across the full continuum of routine TB care.

A systematic review and meta-analysis is therefore warranted to identify, critically appraise, and quantitatively synthesize available evidence on TB treatment outcomes in Cameroon. The review will estimate pooled proportions of treatment success or recovery, death, loss to follow-up, and treatment failure. This synthesis will provide a clearer national evidence base, identify populations and contexts associated with unfavorable outcomes, reveal gaps in the published literature, and support the Cameroon National Tuberculosis Control Programme and other stakeholders in strengthening diagnosis, treatment delivery, patient retention, surveillance, and resource allocation.

## 2. Methods

### 2.1. Study design and registration

This systematic review and meta-analysis was conducted following the Preferred Reporting Items for Systematic Reviews and Meta-Analyses (PRISMA) 2020 guidelines [18]. The protocol was registered in the International Prospective Register of Systematic Reviews (PROSPERO identifier: CRD420261404490).

### 2.2. Eligibility criteria

#### Inclusion criteria

Studies were eligible if they: (1) were conducted in Cameroon; (2) enrolled patients of any age treated for tuberculosis, whether drug-susceptible or drug-resistant, pulmonary or extrapulmonary, new or previously treated, with or without HIV co-infection; (3) used an observational (cohort or cross-sectional for prevalence rate analysis), (4) reported the number of patients with at least one WHO-defined treatment outcome (treatment success [cure and treatment completion], death, treatment failure or loss to follow-up) among patients registered for treatment; (5) were published in English or French; and (6) involved human participants. Case-control studies were eligible for risk factor assessment among the included reports.

#### Exclusion criteria

Case reports, narrative reviews, systematic reviews, meta-analyses, editorials, opinion pieces, conference abstracts, animal studies and in vitro or laboratory-only studies were excluded. We also excluded studies that did not report patient-level treatment outcomes (for example, diagnostic accuracy or drug-resistance surveillance studies without outcome data), studies in which outcome counts for the population of interest could not be extracted (aggregated data or percentages without denominators), and studies with corrupted or incomplete outcome data. Duplicate publications of the same dataset were excluded, and overlapping cohorts were handled as described under data extraction.

### 2.3. Outcomes measurement and definition

The primary outcomes were the pooled proportions of treatment success, death, treatment failure and loss to follow-up (LTFU) among patients registered for treatment. Outcomes were classified according to the WHO definitions [7]. Treatment failure was defined as a patient whose treatment regimen needed to be terminated or permanently changed to a new regimen or treatment strategy. Cured was defined as a patient with pulmonary TB with bacteriologically confirmed TB at the beginning of treatment who completed treatment as recommended by the national policy, with evidence of bacteriologic response and no evidence of failure. Treatment completed was defined as a patient who completed treatment as recommended by the national policy but whose outcome did not meet the definition for cure or treatment failure. Mortality was defined as a patient who died before starting treatment or during the course of treatment. Lost to follow-up (LTFU) was defined as a patient who did not start treatment or whose treatment was interrupted for two consecutive months or more. Treatment success was defined as the sum of all patients cured and treatment completed. The proportion for each outcome was the number of patients with the outcome divided by the number of patients registered for treatment in the study. The secondary outcome was the association between patient characteristics and an unfavorable outcome, defined as death, treatment failure or LTFU [19–21].

### 2.4. Search strategy

A comprehensive search was conducted in PubMed, Embase, Scopus, Web of Science, African Journals Online (AJOL) and the Cochrane Library. Grey literature was searched on Google Scholar, and the reference lists of all included studies were hand-searched. The strategy combined controlled vocabulary (MeSH and Emtree) and free-text terms with Boolean operators around three concepts (tuberculosis, treatment outcome and Cameroon), without restriction on the type of tuberculosis. The PubMed strategy was: (“tuberculosis”[tiab] OR “TB”[tiab] OR “Mycobacterium tuberculosis”[tiab]) AND (“treatment outcome”[tiab] OR “treatment success”[tiab] OR “cure”[tiab] OR “treatment completion”[tiab] OR “mortality”[tiab] OR “death”[tiab] OR “loss to follow-up”[tiab] OR “default”[tiab] OR “treatment failure”[tiab]) AND (“Cameroon”[tiab]). The full strategy for each database is presented in the Supplementary Materials (**Table S1**). The last search was performed on April 15, 2026.

All retrieved records were deduplicated and then screened against the eligibility criteria. Full texts of all potentially eligible articles were independently screened by two reviewers (FZLC and RT). Disagreements were resolved through discussion or by consulting a third reviewer (JTZ).

### 2.5. Data extraction

Data were extracted independently by two reviewers (FZLC and JTZ) using a standardized form. For each study we extracted the first author, year of publication, year of study completion, design, region, participant group (unselected TB patients, previously treated patients, RR-TB, MDR-TB, TB/HIV co-infected patients or children), TB localization (pulmonary, extrapulmonary or mixed), treatment regimen (first-line or second-line), the number of patients registered for treatment, the number with each outcome (success, death, failure and LTFU) and the adjusted odds ratios with 95% confidence intervals for predictors of unfavorable outcomes. Studies are identified by first author last name and year of study completion, with a numeric suffix when an author contributed several studies in the same year. The year of study completion was grouped into five periods (<2005, 2005–2009, 2010– 2014, 2015–2019 and ≥2020) that follow the successive National Strategic Plans against tuberculosis, and studies were classified as single-region (by region) or multi-region. Disagreements were resolved through discussion or by a third reviewer (CA).

When several publications reported the same patients (same site and overlapping study period), or when one cohort was nested in a larger one, a single publication was retained, namely the one with the largest sample and the most complete outcome data, so that no patient contributed more than once to a pooled estimate.

### 2.6. Quality assessment of included reports

The methodological quality of the included studies was assessed independently by two reviewers (FZLC and CA) using the Joanna Briggs Institute (JBI) critical appraisal checklists matched to the study design: the 11-item checklist for cohort studies and the 9-item checklist for prevalence studies [22]. For cohort studies, the checklist contains eleven items assessing: the two groups were similar and recruited from the same population; exposures were measured similarly to assign people to exposed and unexposed groups; the exposure was measured in a valid and reliable way; confounding factors were identified and strategies to deal with them stated; whether the groups/participants were free of the outcome at the start of the study (or at the moment of exposure); outcomes were measured in a valid and reliable way; whether the follow-up time was sufficient for outcomes to occur; follow-up was complete, with reasons for loss to follow-up described and explored; strategies to address incomplete follow-up were utilized; and appropriate statistical analysis was used. Each item was scored 1 (yes) or 0 (no or unclear), and the overall risk of bias was classified as low (≥70% of items scored “yes”), moderate (50–69%) or high (<50%). Disagreements were resolved by discussion or by a third reviewer (RT).

### 2.7. Statistical analysis

Study-level proportions are presented with exact (Clopper–Pearson) 95% confidence intervals. Pooled proportions were estimated with a random-effects generalized linear mixed model (GLMM; random-intercept logistic regression with a logit link, fitted by maximum likelihood), implemented with the “metaprop” function of the “meta” package in R (method = “GLMM”, transformation “PLOGIT”) [23]. This approach models the binomial within-study variability directly and accommodates studies with 0% or 100% event rates without continuity corrections.

Heterogeneity was assessed with the I² statistic, classified as low (<25%), moderate (25–75%) or high (>75%). Because the between-study heterogeneity was high, the pooled proportion represented the mean of the distribution of true proportions across the heterogeneous studied settings and not a single common value.

Subgroup analyses were conducted for each outcome by study period, study design, region (each region or multi-region), participant group, TB localization and treatment regimen, and differences between subgroups were tested with the Q test for subgroup differences. Subgroup estimates were pooled when at least two reports were available, and categories with a single report are shown for completeness only. Subgroup analyses were exploratory and were not adjusted for multiple testing.

For predictors of unfavorable outcomes, adjusted odds ratios (AOR) and their 95% confidence intervals from multivariable models were pooled by inverse-variance random-effects meta-analysis when at least two independent studies reported the same predictor. Reference categories were harmonized across studies (male vs. female; HIV-positive vs. HIV-negative; previous TB treatment vs. new case; age ≤40 vs. >40 years; extrapulmonary vs. pulmonary TB; no vs. yes for co-trimoxazole prophylaxis and for antiretroviral therapy), and odds ratios reported in the opposite direction were inverted. Crude odds ratios and predictors reported by a single study were not pooled. Statistical significance was defined as a *p*-value <0.05. All analyses were conducted in R version 4.6.1 [24].

### 2.8. Publication bias and sensitivity analysis

Small-study effects were examined with funnel plots and with Egger’s regression and Begg’s rank correlation tests for outcomes with at least 10 reports. Because the dependence between a proportion and its variance can produce funnel-plot asymmetry unrelated to publication bias, these tests were interpreted cautiously [9,10], and a *p*-value <0.05 was considered evidence of asymmetry. Robustness was assessed with a leave-one-out analysis.

## 3. Results

### 3.1. Study selection and inclusion

The search yielded 780 records from databases (PubMed: 92; Scopus: 156; Web of Science: 222; Embase: 239; Cochrane: 69; AJOL: 2). After deduplication (163 removed) and screening (617 screened; 551 excluded), 66 full-text reports were sought, of which 66 were assessed and 34 excluded (outcome of interest not reported, n = 26; duplicate data, n = 5; aggregated data, n = 3). From other methods, 47 reports were sought and 43 excluded (outcome not reported). Thirty-two studies were finally included in the systematic review and meta-analysis (**Fig. 1**).

**Fig. 1.**
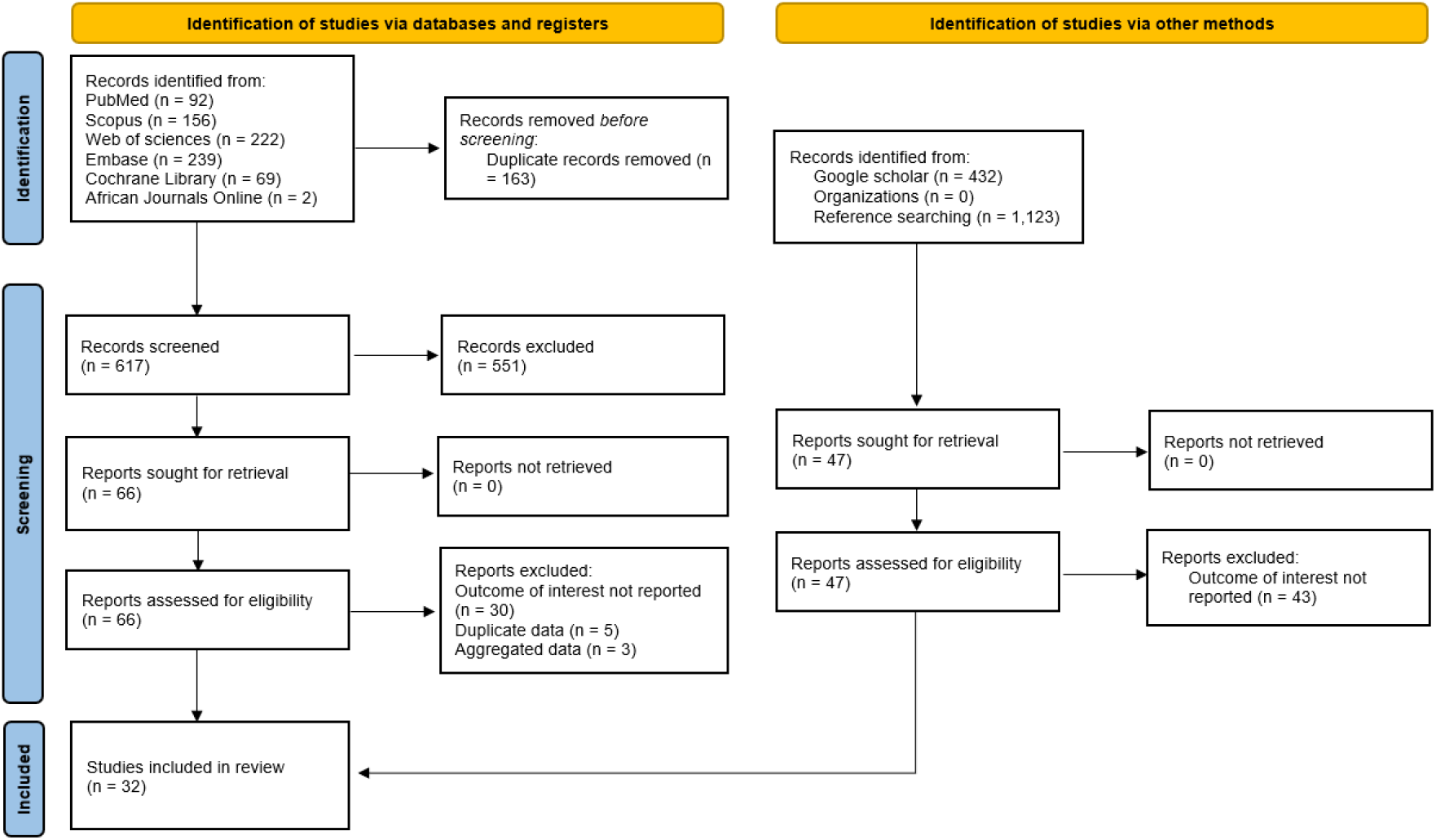
PRISMA diagram flow from study identification to inclusion in the systematic review and meta-analysis [18].

### 3.2. Characteristics of included studies

The 32 studies were conducted among Six out of regions of the country. These included the Centre, Littoral, North-West, South-West, West, North. Six studies were conducted in multi-regional settings (more than one region). Designs were predominantly cohort (n = 25), with cross-sectional studies (n = 7). Most studies enrolled unspecific TB patients (n = 22), with smaller numbers focusing on retreated TB (n = 3), RR-TB (n = 3), MDR-TB (n = 2), TB/HIV-coinfected (n = 3), and pediatric patients (n = 3). Sample sizes ranged from 54 to 26,195 participants. Most studies (n = 27) reported patients on standard first-line regimens and five evaluated second-line regimens. Risk of bias was low in 27 studies, moderate in four, and high in one (**Table 1**).

**Table 1.** Synthesis of included studies characteristics.

| **Author** | **Ref** | **Publication year** | **Study year<sup>1</sup>** | **Design** | **Region** | **Participant** | **Type of TB case** | **Total treated** | **Treatment** | **n (%)** |  |  |  | **Risk of bias** |
|---|---|---|---|---|---|---|---|---|---|---|---|---|---|---|
|  |  |  |  |  |  |  |  |  |  | Death | Success | Failure | Lost to follow-up |  |
| Noeske,1998 | [25] | 2002 | 1998 | Cohort | West | Unspecific TB patient | PTB | 560 | Standard regimen | 47 | 441 | 16 | 50 | Low |
| Kuaban,2007 | [26] | 2009 | 2007 | Cohort | Centre | Unspecific TB patient | PTB | 413 | Standard regimen | 40 | 330 | 10 | 68 | Low |
| Luma,2009 | [27] | 2013 | 2009 | Cross-sectional | Littoral | Unspecific TB patient | EPTB | 54 | Standard regimen | 43 | NR | NR | NR | High |
| Noeske,2009 | [28] | 2012 | 2009 | Cohort | Littoral | Unspecific TB patient | PTB | 216 | Standard regimen | 5 | 216 | 14 | 18 | Low |
| Pefura-Yone,2009_1 | [29] | 2011 | 2009 | Cohort | Centre | Unspecific TB patient | EPTB | 196 | Standard regimen | 7 | 151 | 1 | 27 | Moderate |
| Pefura-Yone,2009_2 | [30] | 2012 | 2009 | Cohort | Centre | Unspecific TB patient | Mixed | 1,647 | Standard regimen | 86 | 1121 | 6 | 331 | Low |
| Kuaban,2011 | [31] | 2015 | 2011 | Cohort | Littoral and Centre | MDR-TB patient | PTB | 150 | Second-line regimen | 10 | 134 | 1 | 5 | Low |
| Atekem,2012 | [32] | 2018 | 2012 | Cross-sectional | South-West | Unspecific TB patient | Mixed | 895 | Standard regimen | NR | 726 | 0 | NR | Low |
| Pefura-Yone,2012 | [33] | 2014 | 2012 | Cohort | Centre | Unspecific TB patient | PTB | 78 | Standard regimen | 1 | 56 | 14 | 7 | Low |
| Agbor,2013 | [34] | 2014 | 2013 | Cohort | Centre | TB/HIV coinfected patients | Mixed | 337 | Standard regimen | 99 | 205 | 1 | 14 | Low |
| Bigna,2013 | [35] | 2015 | 2013 | Cohort | Centre | TB/HIV coinfected patients | Mixed | 337 | Standard regimen | 99 | NR | NR | NR | Low |
| Essomba,2013 | [36] | 2017 | 2013 | Cross-sectional | Littoral | Unspecific TB patient | Mixed | 359 | Standard regimen | 14 | 244 | 3 | 83 | Low |
| Ngahane,2013 | [37] | 2016 | 2013 | Cohort | Littoral | Unspecific TB patient | Mixed | 8,902 | Standard regimen | 722 | 6,684 | 77 | 911 | Low |
| Balkissou,2015 | [38] | 2022 | 2015 | Cohort | West and North | Unspecific TB patient | Mixed | 19,681 | Standard regimen | 1,565 | 16,930 | 222 | 560 | Low |
| Essomba,2015 | [36] | 2017 | 2015 | Cross-sectional | Littoral | Unspecific TB patient | Mixed | 359 | Standard regimen | 14 | 244 | 3 | 83 | Low |
| Kuaban,2015 | [39] | 2020 | 2015 | Cohort | North-West, South-West, West, and Littoral | Retreated TB patient | PTB | 753 | Standard regimen | 63 | 573 | 8 | 89 | Low |
| Merker,2015 | [40] | 2021 | 2015 | Cohort | North-West | RR-TB patient | PTB | 195 | Second-line regimen | 15 | 126 | 3 | 5 | Low |
| Balkissou,2016 | [38] | 2022 | 2016 | Cohort | North | Pediatric patients | Mixed | 668 | Standard regimen | 14 | 641 | 1 | 12 | Low |
| Endale,2016 | [41] | 2022 | 2016 | Cohort | Centre, West, and North | Unspecific TB patient | Mixed | 26,195 | Standard regimen | 2,171 | 22,312 | 285 | 1,427 | Low |
| Mbuh,2017 | [42] | 2019 | 2017 | Cross-sectional | Littoral | Unspecific TB patient | EPTB | 109 | Standard regimen | 5 | 38 | 0 | 2 | Low |
| Tanue,2017 | [8] | 2019 | 2017 | Cohort | South-West | Unspecific TB patient | NR | 1041 | Standard regimen | 72 | 818 | 16 | 99 | Moderate |
| Foncha,2019 | [43] | 2021 | 2019 | Cohort | Littoral | Unspecific TB patient | Mixed | 3,321 | Standard regimen | 183 | 2,759 | 22 | 164 | Low |
| Jouego,2019 | [44] | 2022 | 2019 | Cohort | Multiregion | RR-TB patient | PTB | 709 | Second-line regimen | 79 | 602 | 2 | 26 | Moderate |
| Kuaban,2019 | [17] | 2021 | 2019 | Cohort | Centre | RR-TB patient | Mixed | 242 | Second-line regimen | 31 | 193 | 10 | 8 | Moderate |
| Meriki,2014 | [45] | 2019 | 2019 | Cohort | North-West and South-West | TB/HIV coinfected patients | NR | 209 | Standard regimen | 71 | NR | NR | NR | Low |
| Mpoh,2019 | [46] | 2023 | 2019 | Cohort | Centre | MDR-TB patient | Mixed | 107 | Second-line regimen | 3 | NR | NR | NR | Low |
| Ngangue,2020 | [47] | 2022 | 2020 | Cross-sectional | North-West, South-West, and Littoral | Unspecific TB patient | PTB | 274 | Standard regimen | 7 | NR | NR | 10 | Low |
| Poka-Mayap,2020 | [48] | 2025 | 2020 | Cohort | Centre | Pediatric patients | Mixed | 881 | Standard regimen | 40 | 665 | 2 | 174 | Low |
| Vukugah,2020 | [49] | 2022 | 2020 | Cohort | Centre | Pediatric patients | Mixed | 610 | Standard regimen | 73 | 488 | 4 | 25 | Low |
| Abessolo,2022 | [50] | 2023 | 2022 | Cross-sectional | Centre | Unspecific TB patient | Mixed | 395 | Standard regimen | 75 | 311 | NR | NR | Low |
| Chethkwo,2022 | [51] | 2024 | 2022 | Cohort | North-West | Unspecific TB patient | Mixed | 2,428 | Standard regimen | 200 | 2,119 | 10 | 99 | Low |
| Mbuh,2022 | [52] | 2024 | 2022 | Cohort | Littoral | Unspecific TB patient | Mixed | 444 | Second-line regimen | 32 | 386 | 6 | 19 | Low |

*1: Year of study completion; Ref: Reference; TB: Mycobacterium tuberculosis; FQR-Mtr: Fluoroquinolone-resistant modified treatment regimen; SLIr-Mtr: Second-line injectable-resistant modified treatment regimen; RR-TB: Rifampicin-resistant Mycobacterium tuberculosis; MDR-TB: Multidrug-resistant Mycobacterium tuberculosis; NR; Not reported; PTB: Pulmonary Mycobacterium tuberculosis; EPTB: Extra-Pulmonary Mycobacterium tuberculosis; The sum of all outcomes in some studies does not reach 100% because some patients were transferred out during treatment.*

### 3.3. Tuberculosis mortality

The pooled mortality rate was 8.1% (95% CI: 5.7–11.3) among 71,870 participants from 31 reports, with high heterogeneity (I² = 96.4%; *p* < 0.001) (**Fig 2**). The pooled estimate was robust in the sensitivity analysis, and no evidence of publication bias was observed on the funnel plot or according to Egger’s (p = 0.543) and Begg’s (p = 0.424) tests (**Figs. S1-2**).

**Fig. 2.**
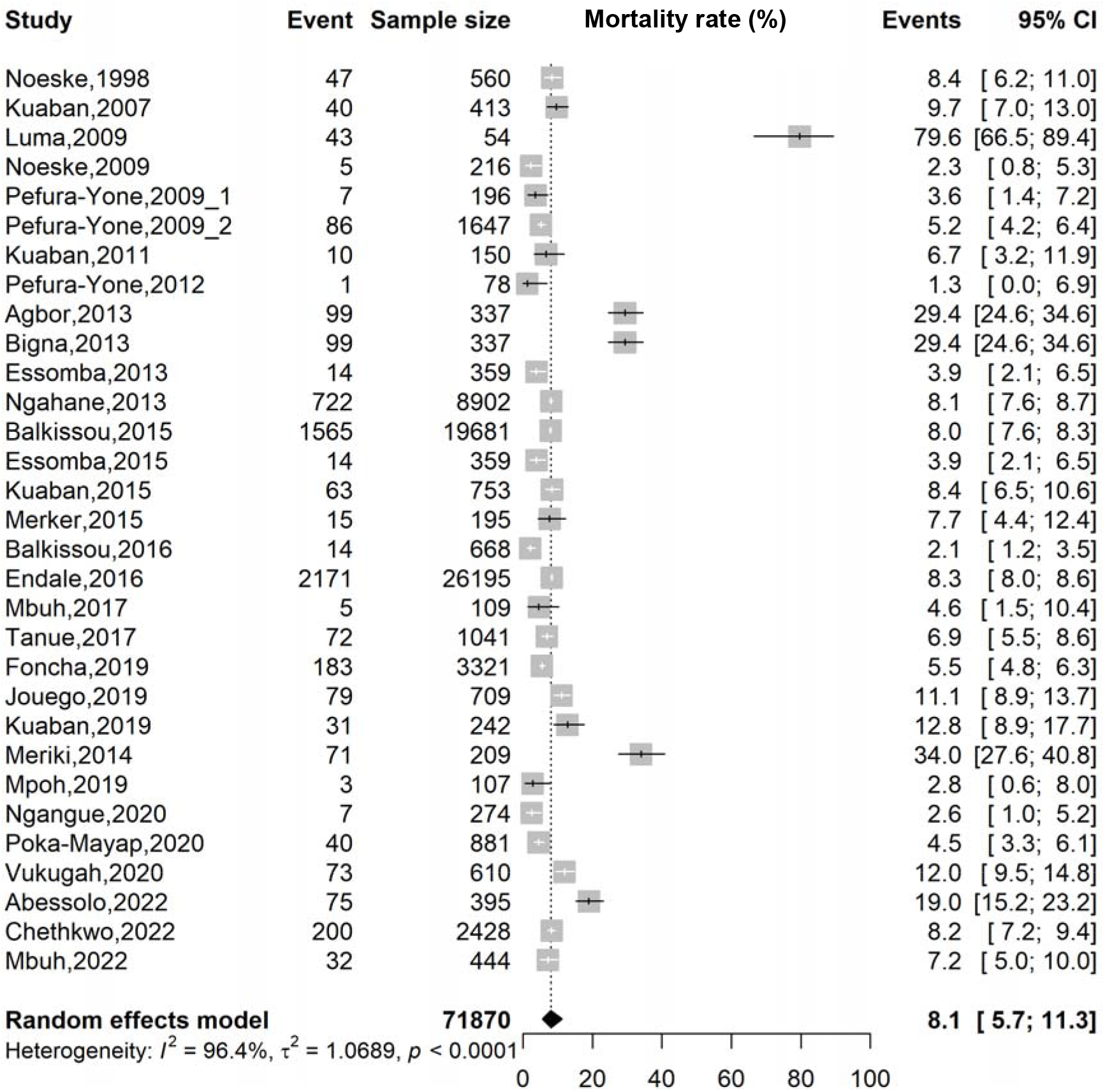
Forest plot of the pooled mortality rate among tuberculosis patients in Cameroon, 1998-2022.

Subgroup analyses showed significant differences by region (*p* < 0.001) and participant type (p < 0.001). Mortality was highest in the South-West (16.2%; 95% CI: 4.8– 42.6; 2 studies; n = 1,250) and among TB/HIV-coinfected patients (30.5%; 95% CI: 27.5– 33.6; 3 studies; n = 883), and lowest in the North and West regions and among pediatric patients (5.0%; 95%CI: 2.2-11.0; 3 studies; n = 2,159). The regional pattern should be interpreted with caution, as the North and West regions were each represented by only one study, warranting cautious interpretation (**Table 2 and Figs S3-8**).

**Table 2.**
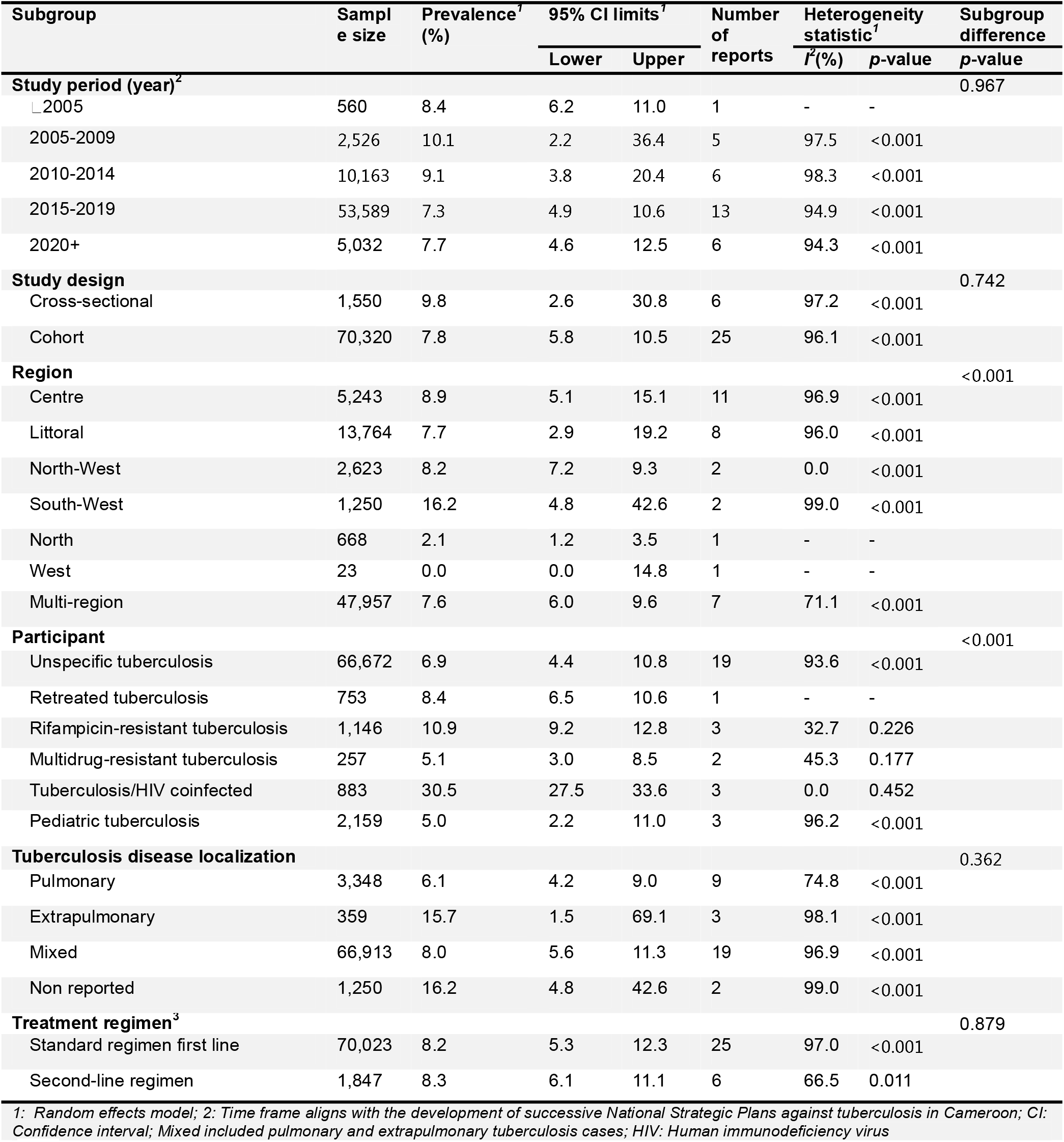
Subgroup meta-analysis of mortality rate among tuberculosis patients in Cameroon, 1998–2022.

| Subgroup | Sample size | Prevalence<sup>1</sup> (%) | 95% CI limits<sup>1</sup> |  | Number of reports | Heterogeneity statistic<sup>1</sup> |  | Subgroup difference |
|---|---|---|---|---|---|---|---|---|
|  |  |  | Lower | Upper |  | *I*<sup>2</sup>(%) | *p*-value | *p*-value |
| **Study period (year)<sup>2</sup>** |  |  |  |  |  |  |  | 0.967 |
| ⎿2005 | 560 | 8.4 | 6.2 | 11.0 | 1 | - | - |  |
| 2005-2009 | 2,526 | 10.1 | 2.2 | 36.4 | 5 | 97.5 | <0.001 |  |
| 2010-2014 | 10,163 | 9.1 | 3.8 | 20.4 | 6 | 98.3 | <0.001 |  |
| 2015-2019 | 53,589 | 7.3 | 4.9 | 10.6 | 13 | 94.9 | <0.001 |  |
| 2020+ | 5,032 | 7.7 | 4.6 | 12.5 | 6 | 94.3 | <0.001 |  |
| **Study design** |  |  |  |  |  |  |  | 0.742 |
| Cross-sectional | 1,550 | 9.8 | 2.6 | 30.8 | 6 | 97.2 | <0.001 |  |
| Cohort | 70,320 | 7.8 | 5.8 | 10.5 | 25 | 96.1 | <0.001 |  |
| **Region** |  |  |  |  |  |  |  | <0.001 |
| Centre | 5,243 | 8.9 | 5.1 | 15.1 | 11 | 96.9 | <0.001 |  |
| Littoral | 13,764 | 7.7 | 2.9 | 19.2 | 8 | 96.0 | <0.001 |  |
| North-West | 2,623 | 8.2 | 7.2 | 9.3 | 2 | 0.0 | <0.001 |  |
| South-West | 1,250 | 16.2 | 4.8 | 42.6 | 2 | 99.0 | <0.001 |  |
| North | 668 | 2.1 | 1.2 | 3.5 | 1 | - | - |  |
| West | 23 | 0.0 | 0.0 | 14.8 | 1 | - | - |  |
| Multi-region | 47,957 | 7.6 | 6.0 | 9.6 | 7 | 71.1 | <0.001 |  |
| **Participant** |  |  |  |  |  |  |  | <0.001 |
| Unspecific tuberculosis | 66,672 | 6.9 | 4.4 | 10.8 | 19 | 93.6 | <0.001 |  |
| Retreated tuberculosis | 753 | 8.4 | 6.5 | 10.6 | 1 | - | - |  |
| Rifampicin-resistant tuberculosis | 1,146 | 10.9 | 9.2 | 12.8 | 3 | 32.7 | 0.226 |  |
| Multidrug-resistant tuberculosis | 257 | 5.1 | 3.0 | 8.5 | 2 | 45.3 | 0.177 |  |
| Tuberculosis/HIV coinfected | 883 | 30.5 | 27.5 | 33.6 | 3 | 0.0 | 0.452 |  |
| Pediatric tuberculosis | 2,159 | 5.0 | 2.2 | 11.0 | 3 | 96.2 | <0.001 |  |
| **Tuberculosis disease localization** |  |  |  |  |  |  |  | 0.362 |
| Pulmonary | 3,348 | 6.1 | 4.2 | 9.0 | 9 | 74.8 | <0.001 |  |
| Extrapulmonary | 359 | 15.7 | 1.5 | 69.1 | 3 | 98.1 | <0.001 |  |
| Mixed | 66,913 | 8.0 | 5.6 | 11.3 | 19 | 96.9 | <0.001 |  |
| Non reported | 1,250 | 16.2 | 4.8 | 42.6 | 2 | 99.0 | <0.001 |  |
| **Treatment regimen<sup>3</sup>** |  |  |  |  |  |  |  | 0.879 |
| Standard regimen first line | 70,023 | 8.2 | 5.3 | 12.3 | 25 | 97.0 | <0.001 |  |
| Second-line regimen | 1,847 | 8.3 | 6.1 | 11.1 | 6 | 66.5 | 0.011 |  |

*1:  Random effects model; 2: Time frame aligns with the development of successive National Strategic Plans against tuberculosis in Cameroon; CI: Confidence interval; Mixed included pulmonary and extrapulmonary tuberculosis cases; HIV: Human immunodeficiency virus*

### 3.4. Tuberculosis treatment success

The pooled treatment success rate was 78.2% (95% CI: 73.8–82.1) among 71,784 participants from 27 reports, with high heterogeneity (I² = 98.2%; *p* < 0.001) (**Fig. 3**). Sensitivity analysis revealed that the overall pooled estimate was robust and not influenced by any single study. However, Egger’s test identified a possible risk of publication bias (*p* = 0.036), associated with an asymmetrical funnel plot. To adjust the pooled estimate, the traditional Duval and Tweedie trim-and-fill analysis imputed eight potentially missing studies, resulting in an adjusted pooled treatment success rate of 83.1% (95% CI: 78.8–86.6; I² = 98.4%; *p* < 0.001) (**Figs. S10–11**).

**Fig. 3.**
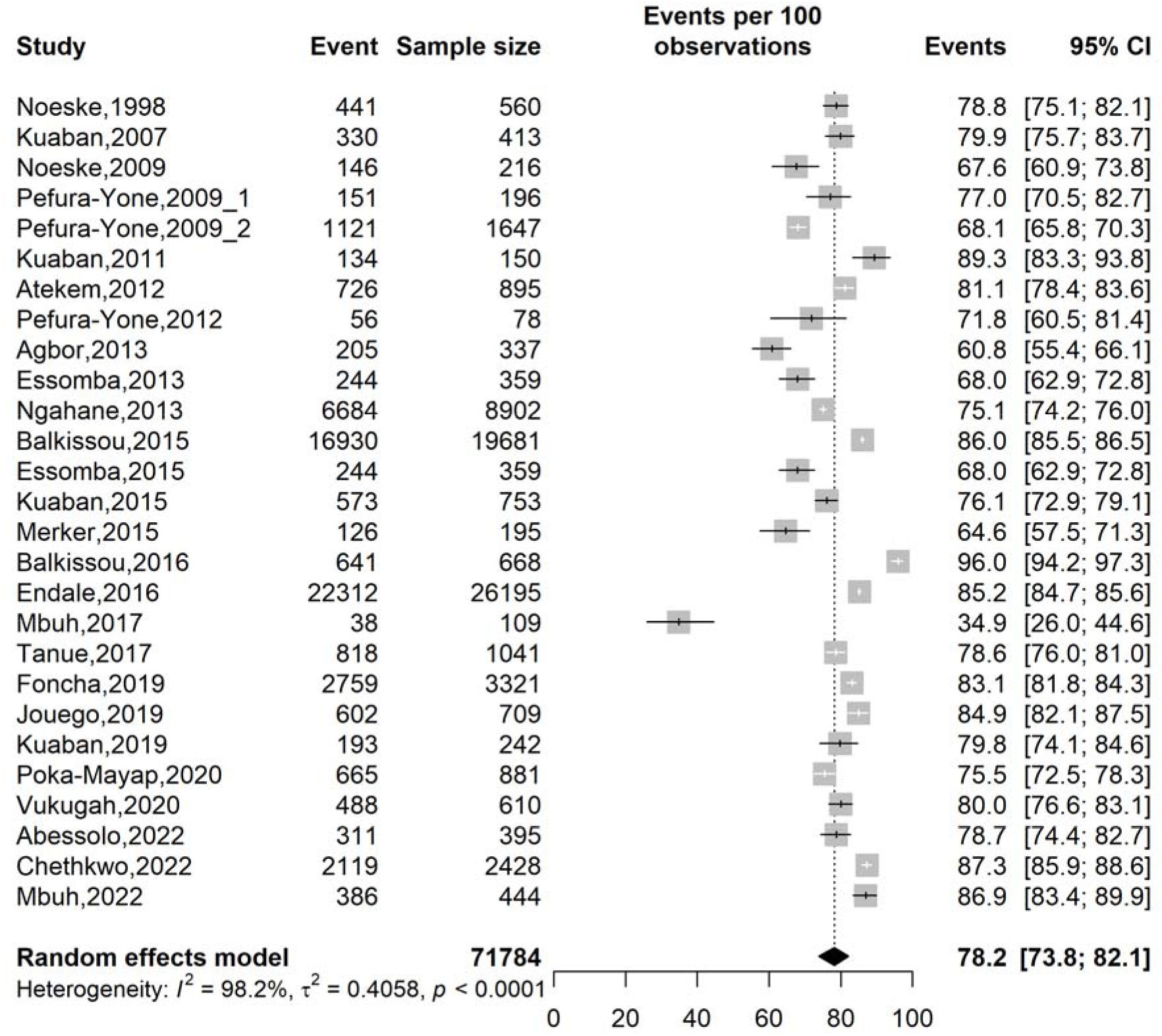
Forest plot of the pooled treatment success rate among tuberculosis patients in Cameroon, 1998-2022.

The region of study conduction and the type of study participants significantly influenced the heterogeneity of the pooled estimates (p < 0.001). By participant type, treatment success was highest among MDR-TB patients (89.3%; 95% CI: 83.3–93.8; 1 study; n = 150) and pediatric patients (86.8%; 95% CI: 70.3–94.8; 3 studies; n = 2,159), and lowest among TB/HIV-coinfected patients (60.8%; 95% CI: 55.4–66.1; 1 study; n = 337). By region, treatment success was highest in the North (96.0%; 95% CI: 94.2–97.3; 1 study; n = 668) and North-West (87.3%; 95% CI: 85.9–88.6; 1 study; n = 2,428), and lowest in the Littoral (71.1%; 95% CI: 58.9–80.9; 7 studies; n = 13,710). However, these findings should be interpreted with caution because of the high heterogeneity observed and the fact that some of the subgroups were derived from only one study (**Table 3 and Figs. S12–17**).

**Table 3.** Subgroup meta-analysis of treatment success rate among tuberculosis patients in Cameroon, 1998–2022.

| **Subgroup** | **Sample size** | **Prevalence<sup>1</sup> (%)** | **95% CI limits<sup>1</sup>** |  | **Number of reports** | **Heterogeneity statistic<sup>1</sup>** |  | **Subgroup difference** |
|  |  |  | **Lower** | **Upper** |  | ***I*<sup>2</sup>(%)** | ***p*-value** | ***p*-value** |
|---|---|---|---|---|---|---|---|---|
| **Study period (year)**<sup>2</sup> |  |  |  |  |  |  |  | 0.131 |
| ∟2005 | 560 | 78.8 | 75.1 | 82.1 | 1 | - | - |  |
| 2005-2009 | 2,472 | 73.3 | 67.5 | 78.5 | 4 | 88.8 | <0.001 |  |
| 2010-2014 | 10,721 | 75.4 | 66.9 | 82.2 | 6 | 93.5 | <0.001 |  |
| 2015-2019 | 53,273 | 79.2 | 69.4 | 86.4 | 11 |  |  |  |
| 2020+ | 4,758 | 82.2 | 77.6 | 86.0 | 5 | 95.0 | <0.001 |  |
| **Study design** |  |  |  |  |  |  |  | 0.057 |
| Cross-sectional | 2,117 | 67.8 | 52.8 | 79.8 | 5 | 96.4 | <0.001 |  |
| Cohort | 69,667 | 80.2 | 76.2 | 83.7 | 22 | 98.3 | <0.001 |  |
| **Region** |  |  |  |  |  |  |  | <0.001 |
| Centre | 4,799 | 75.1 | 70.9 | 78.8 | 9 | 90.7 | <0.001 |  |
| Littoral | 13,710 | 71.1 | 58.9 | 80.9 | 7 | 97.4 | <0.001 |  |
| North-West | 2,428 | 87.3 | 85.9 | 88.6 | 1 | - | - |  |
| South-West | 1,936 | 79.8 | 77.9 | 81.5 | 2 | 47.9 | 0.166 |  |
| North | 668 | 96.0 | 94.2 | 97.3 | 1 | - | - |  |
| West | 560 | 78.8 | 75.1 | 82.1 | 1 | - | - |  |
| Multi-region | 47,683 | 82.1 | 75.7 | 87.1 | 6 | 95.8 | <0.001 |  |
| **Participant** |  |  |  |  |  |  |  | <0.001 |
| Unspecific tuberculosis | 67,239 | 76.9 | 71.9 | 81.2 | 18 | 76.9 | <0.001 |  |
| Retreated tuberculosis | 753 | 76.1 | 72.9 | 79.1 | 1 | - | - |  |
| Rifampicin-resistant tuberculosis | 1,146 | 77.7 | 67.1 | 85.6 | 3 | - | - |  |
| Multidrug-resistant tuberculosis | 150 | 89.3 | 83.3 | 93.8 | 1 | - | - |  |
| Tuberculosis/HIV coinfected | 337 | 60.8 | 55.4 | 66.1 | 1 | - | - |  |
| Pediatric tuberculosis | 2,159 | 86.8 | 70.3 | 94.8 | 3 | 97.9 | <0.001 |  |
| **Tuberculosis disease localization** |  |  |  |  |  |  |  | 0.395 |
| Pulmonary | 3,074 | 77.6 | 71.7 | 82.6 | 8 | 89.6 | <0.001 |  |
| Extrapulmonary | 305 | 57.4 | 27.3 | 82.9 | 2 | 97.9 | <0.001 |  |
| Mixed | 67,364 | 80.4 | 75.2 | 84.7 | 16 | 98.8 | <0.001 |  |
| Not reported | 1,041 | 78.6 | 76.0 | 81.0 | 1 | - | - |  |
| **Treatment regimen**<sup>3</sup> |  |  |  |  |  |  |  | 0.970 |
| Standard regimen first line | 70,044 | 77.2 | 72.0 | 81.7 | 22 | 98.7 | <0.001 |  |
| Second-line regimen | 1,740 | 82.3 | 74.6 | 88.1 | 5 | 92.7 | <0.001 |  |

*1: Random effects model; 2: Time frame aligns with the development of successive National Strategic Plans against tuberculosis in Cameroon; CI: Confidence interval; Mixed included pulmonary and extrapulmonary tuberculosis cases; HIV: Human immunodeficiency virus*

### 3.5. Tuberculosis treatment failure

The pooled treatment failure rate was 0.9% (95% CI: 0.5–1.4) among 71,389 participants from 26 reports, with high heterogeneity (I² = 89.1%; *p* < 0.001) (**Fig. 4**). The pooled estimate was robust at sensitivity analysis and showed no sign of publication bias at funnel plot, nor for the Egger’s (*p* = 0.957), Begg’s tests (*p* = 0.378) (**Figs. S18–19**).

**Fig. 4.**
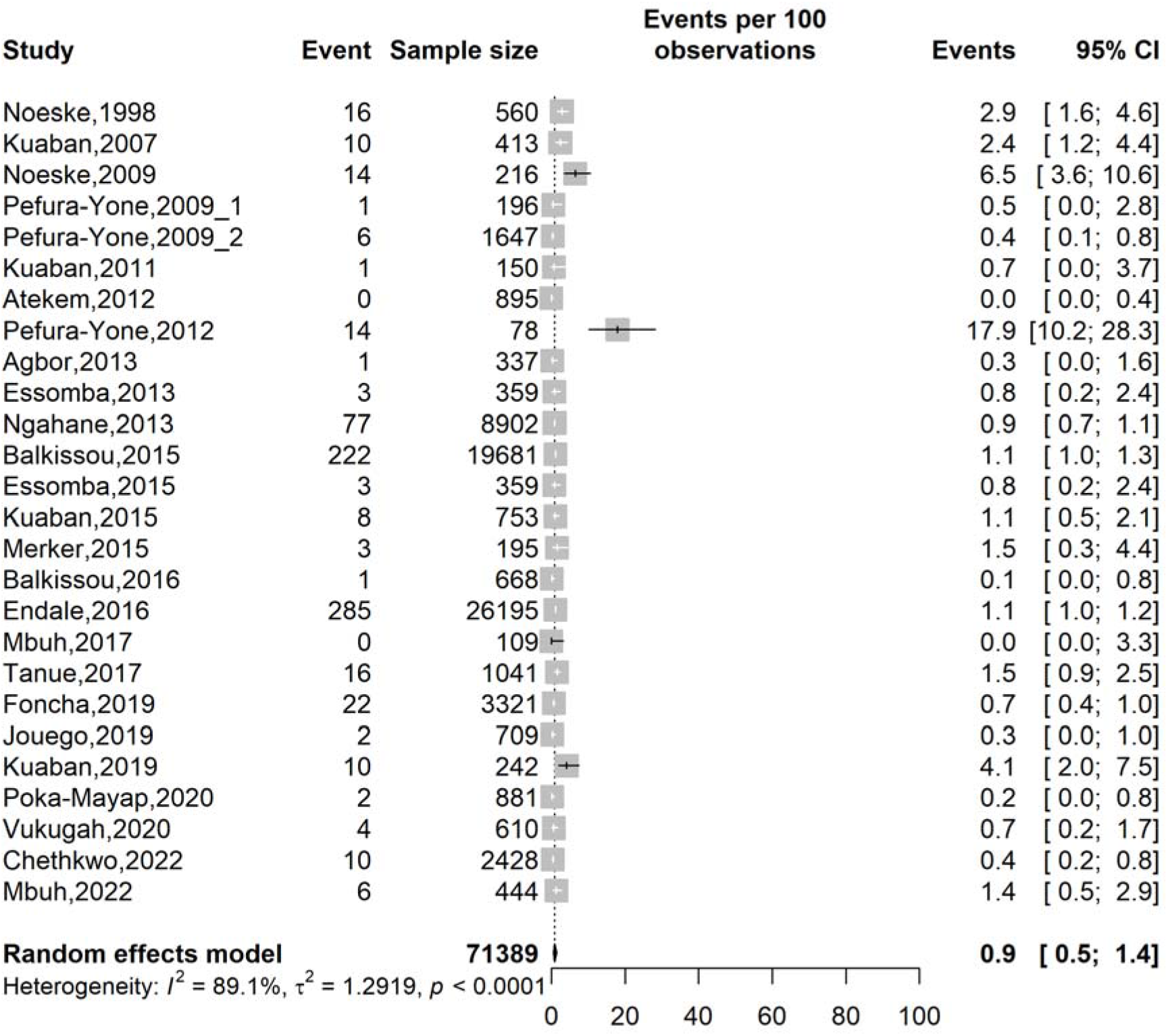
Forest plot of the pooled treatment failure rate among tuberculosis patients in Cameroon, 1998-2022

The subgroup analysis revealed that the failure rate declined from 2.9% (95% CI: 1.6– 4.6; 1 study; n = 560) before 2005 to 1.4% (2005–2009), 0.6% (2010–2014), 0.9% (2015–2019), and 0.5% (95% CI: 0.3–0.9; 4 studies; n = 4,363) from 2020 onward, with a significant period effect (p < 0.001). By localization, pulmonary TB had a higher failure rate (2.1%; 95% CI: 0.9–5.0; 8 studies; n = 3,074) than extrapulmonary TB (0.3%; 95% CI: 0.0–2.3; 2 studies; n = 305). There was a significant regional disparity in tuberculosis treatment failure (p < 0.001): the highest rate was observed in the West region (2.9%; 95% CI: 1.6–4.6; 1 study; n = 560), while the lowest rates were recorded in the South-West (0.1%; 95% CI: 0.0–14.2; 2 studies; n = 1,936) and North region (0.1%; 95% CI: 0.0–0.8; 1 study; n = 668). These subgroup differences should be interpreted with caution, as some were derived from only one study (**Table 4 and Figs. S20-25**).

**Table 4.**
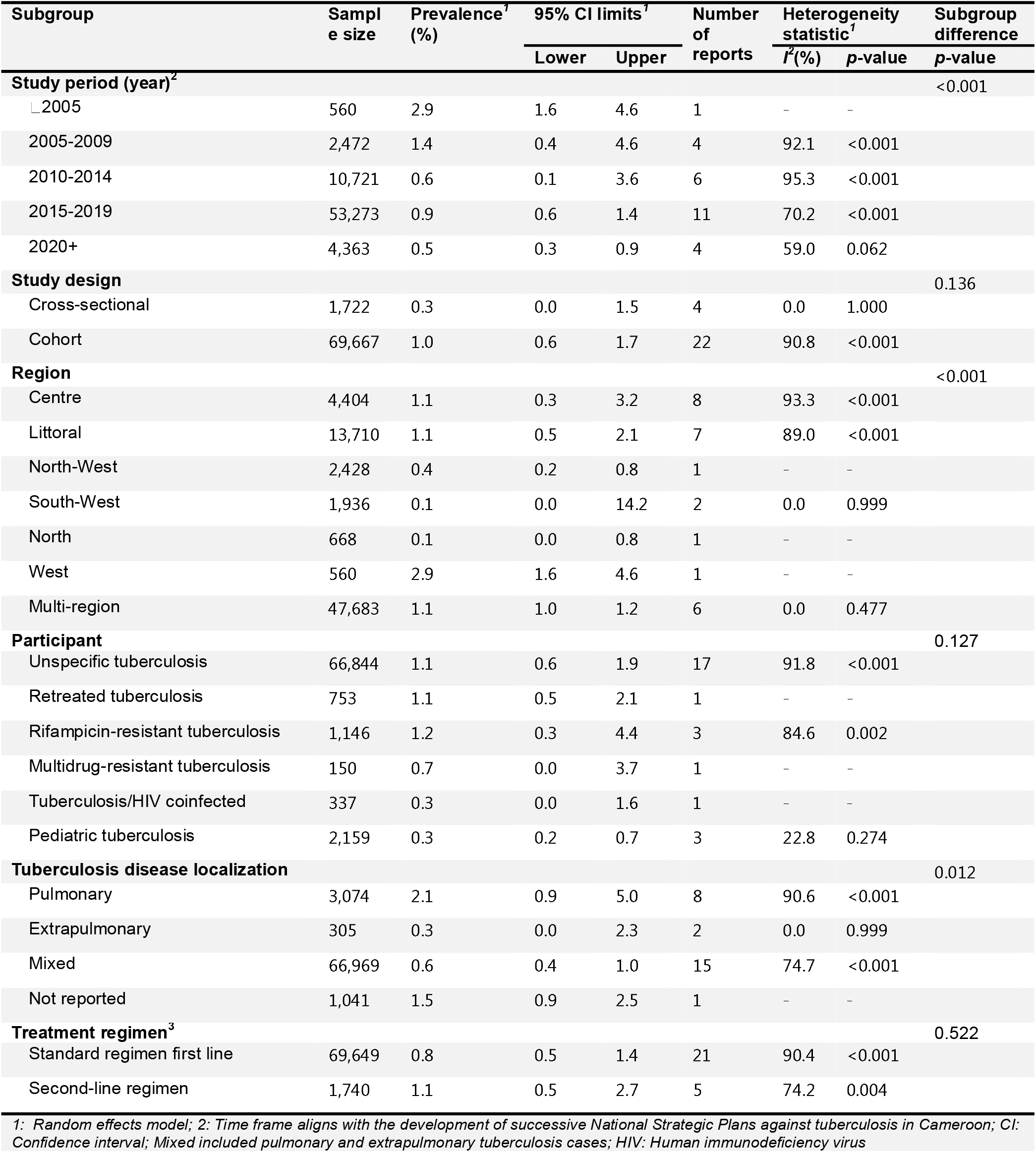
Subgroup meta-analysis of treatment failure rate among tuberculosis patients in Cameroon, 1998–2022.

| **Subgroup** | **Sample size** | **Prevalence<sup>1</sup> (%)** | **95% CI limits<sup>1</sup>** |  | **Number of reports** | **Heterogeneity statistic<sup>1</sup>** |  | **Subgroup difference** |
|---|---|---|---|---|---|---|---|---|
|  |  |  | **Lower** | **Upper** |  | ***I***<sup>2</sup>**(%)** | ***p*-value** | ***p*-value** |
| **Study period (year)<sup>2</sup>** |  |  |  |  |  |  |  | <0.001 |
| ∟2005 | 560 | 2.9 | 1.6 | 4.6 | 1 | - | - |  |
| 2005-2009 | 2,472 | 1.4 | 0.4 | 4.6 | 4 | 92.1 | <0.001 |  |
| 2010-2014 | 10,721 | 0.6 | 0.1 | 3.6 | 6 | 95.3 | <0.001 |  |
| 2015-2019 | 53,273 | 0.9 | 0.6 | 1.4 | 11 | 70.2 | <0.001 |  |
| 2020+ | 4,363 | 0.5 | 0.3 | 0.9 | 4 | 59.0 | 0.062 |  |
| **Study design** |  |  |  |  |  |  |  | 0.136 |
| Cross-sectional | 1,722 | 0.3 | 0.0 | 1.5 | 4 | 0.0 | 1.000 |  |
| Cohort | 69,667 | 1.0 | 0.6 | 1.7 | 22 | 90.8 | <0.001 |  |
| **Region** |  |  |  |  |  |  |  | <0.001 |
| Centre | 4,404 | 1.1 | 0.3 | 3.2 | 8 | 93.3 | <0.001 |  |
| Littoral | 13,710 | 1.1 | 0.5 | 2.1 | 7 | 89.0 | <0.001 |  |
| North-West | 2,428 | 0.4 | 0.2 | 0.8 | 1 | - | - |  |
| South-West | 1,936 | 0.1 | 0.0 | 14.2 | 2 | 0.0 | 0.999 |  |
| North | 668 | 0.1 | 0.0 | 0.8 | 1 | - | - |  |
| West | 560 | 2.9 | 1.6 | 4.6 | 1 | - | - |  |
| Multi-region | 47,683 | 1.1 | 1.0 | 1.2 | 6 | 0.0 | 0.477 |  |
| **Participant** |  |  |  |  |  |  |  | 0.127 |
| Unspecific tuberculosis | 66,844 | 1.1 | 0.6 | 1.9 | 17 | 91.8 | <0.001 |  |
| Retreated tuberculosis | 753 | 1.1 | 0.5 | 2.1 | 1 | - | - |  |
| Rifampicin-resistant tuberculosis | 1,146 | 1.2 | 0.3 | 4.4 | 3 | 84.6 | 0.002 |  |
| Multidrug-resistant tuberculosis | 150 | 0.7 | 0.0 | 3.7 | 1 | - | - |  |
| Tuberculosis/HIV coinfected | 337 | 0.3 | 0.0 | 1.6 | 1 | - | - |  |
| Pediatric tuberculosis | 2,159 | 0.3 | 0.2 | 0.7 | 3 | 22.8 | 0.274 |  |
| **Tuberculosis disease localization** |  |  |  |  |  |  |  | 0.012 |
| Pulmonary | 3,074 | 2.1 | 0.9 | 5.0 | 8 | 90.6 | <0.001 |  |
| Extrapulmonary | 305 | 0.3 | 0.0 | 2.3 | 2 | 0.0 | 0.999 |  |
| Mixed | 66,969 | 0.6 | 0.4 | 1.0 | 15 | 74.7 | <0.001 |  |
| Not reported | 1,041 | 1.5 | 0.9 | 2.5 | 1 | - | - |  |
| **Treatment regimen<sup>3</sup>** |  |  |  |  |  |  |  | 0.522 |
| Standard regimen first line | 69,649 | 0.8 | 0.5 | 1.4 | 21 | 90.4 | <0.001 |  |
| Second-line regimen | 1,740 | 1.1 | 0.5 | 2.7 | 5 | 74.2 | 0.004 |  |

*1:  Random effects model; 2: Time frame aligns with the development of successive National Strategic Plans against tuberculosis in Cameroon; CI: Confidence interval; Mixed included pulmonary and extrapulmonary tuberculosis cases; HIV: Human immunodeficiency virus*

### 3.6. Tuberculosis loss to follow-up

The pooled LTFU rate was 6.8% (95% CI: 5.0–9.1) among 70,768 participants from 26 reports, with high heterogeneity (I² = 98.6%; *p* < 0.001). LTFU rates varied across study periods but were generally lower in more recent periods (**Fig. 5**). No single study significantly influenced the pooled estimates in the sensitivity analysis. Moreover, no evidence of publication bias was detected (Egger’s test, *p* = 0.575; Begg’s test, *p* = 0.378) (**Figs. S26-27**).

**Fig. 5.**
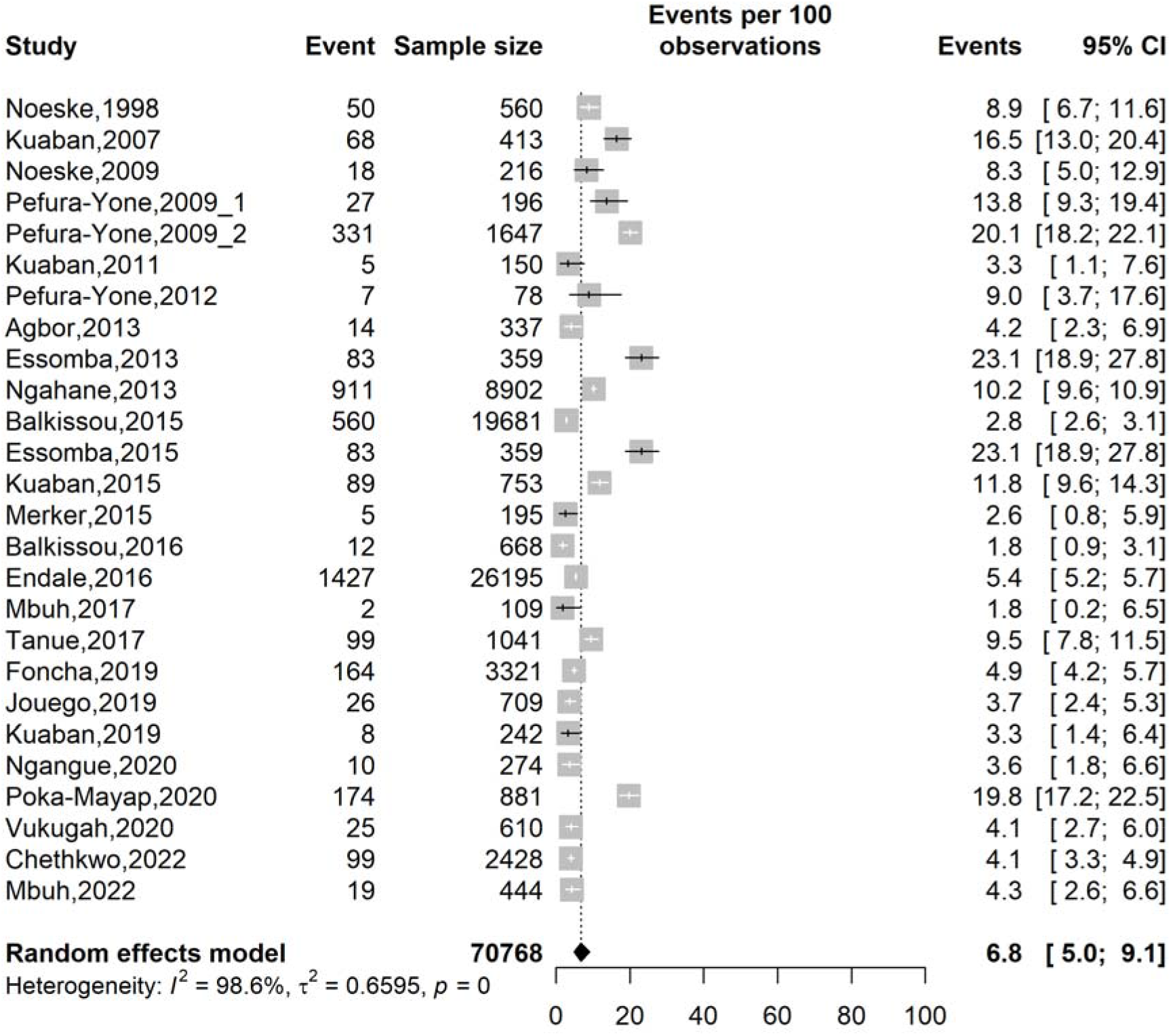
Forest plot of the pooled loss to follow-up rate among tuberculosis patients in Cameroon, 1998-2022.

The study timeframe, the region, type of participants and treatment regimen were significant sources of heterogeneity (*p* < 0.001). By participant type, LTFU was highest among retreated TB patients (11.8%; 95% CI: 9.6–14.3; 17 studies; n = 753) and lowest among RR-TB (3.4%; 95% CI: 2.5–4.6; 3 studies; n = 1,146) and MDR-TB (3.3%; 95% CI: 1.1–7.6; 1 study; n = 150). By regimen, second-line regimens had lower LTFU (3.6%; 95% CI: 2.8–4.6; 5 studies; n = 1,740) than first-line regimens (7.9%; 95% CI: 5.6–10.9; 21 studies; n = 69,028) (**Table 5 and Figs. S28-33**).

**Table 5.**
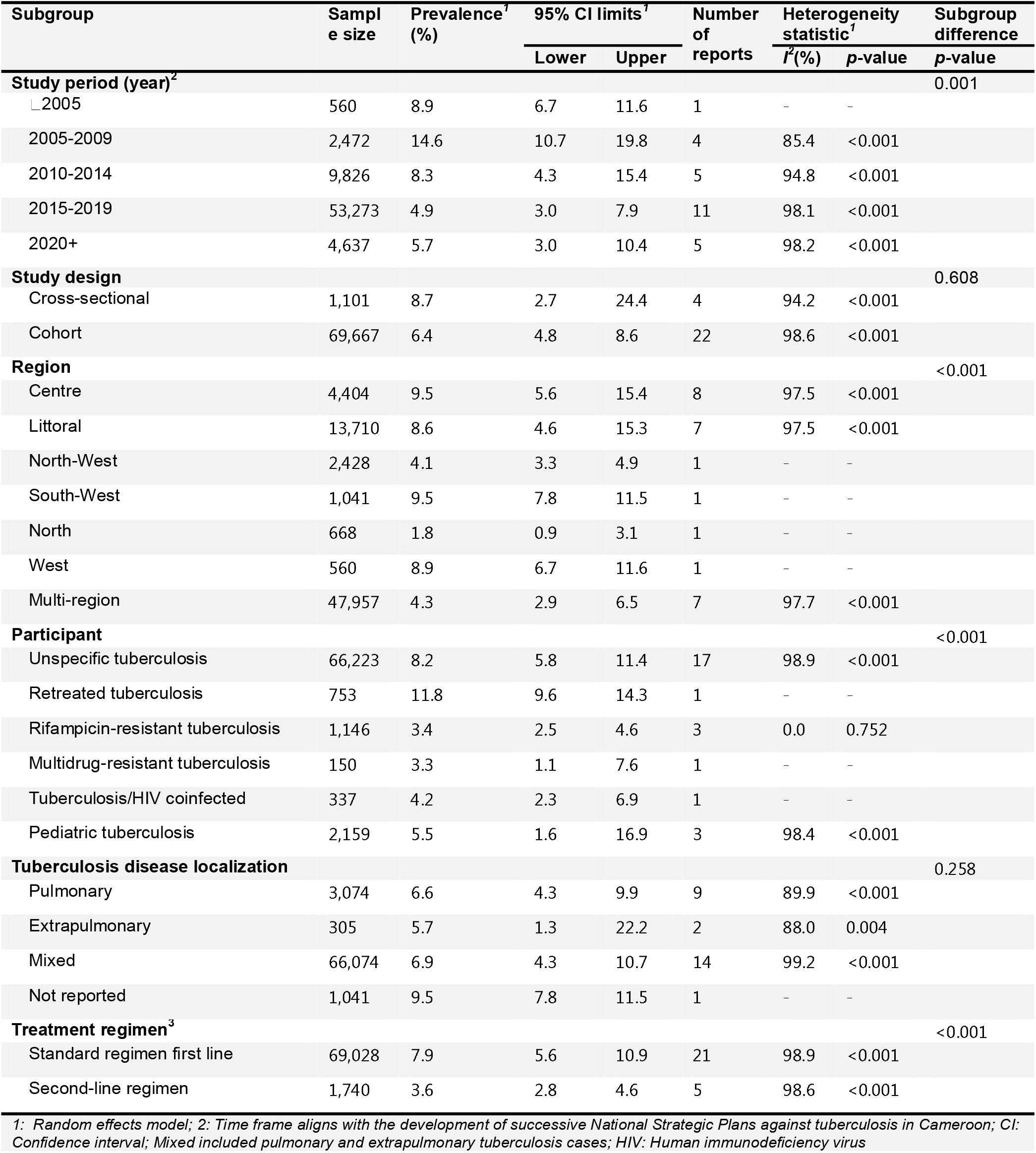
Subgroup meta-analysis of loss to follow-up rate among tuberculosis patients in Cameroon, 1998–2022.

| **Subgroup** | **Sample size** | **Prevalence<sup>1</sup> (%)** | **95% CI limits<sup>1</sup>** |  | **Number of reports** | **Heterogeneity statistic<sup>1</sup>** |  | **Subgroup difference** |
|---|---|---|---|---|---|---|---|---|
|  |  |  | **Lower** | **Upper** |  | ***I*<sup>2</sup>(%)** | ***p*-value** | ***p*-value** |
| **Study period (year)<sup>2</sup>** |  |  |  |  |  |  |  | 0.001 |
| ∟2005 | 560 | 8.9 | 6.7 | 11.6 | 1 | - | - |  |
| 2005-2009 | 2,472 | 14.6 | 10.7 | 19.8 | 4 | 85.4 | <0.001 |  |
| 2010-2014 | 9,826 | 8.3 | 4.3 | 15.4 | 5 | 94.8 | <0.001 |  |
| 2015-2019 | 53,273 | 4.9 | 3.0 | 7.9 | 11 | 98.1 | <0.001 |  |
| 2020+ | 4,637 | 5.7 | 3.0 | 10.4 | 5 | 98.2 | <0.001 |  |
| **Study design** |  |  |  |  |  |  |  | 0.608 |
| Cross-sectional | 1,101 | 8.7 | 2.7 | 24.4 | 4 | 94.2 | <0.001 |  |
| Cohort | 69,667 | 6.4 | 4.8 | 8.6 | 22 | 98.6 | <0.001 |  |
| **Region** |  |  |  |  |  |  |  | <0.001 |
| Centre | 4,404 | 9.5 | 5.6 | 15.4 | 8 | 97.5 | <0.001 |  |
| Littoral | 13,710 | 8.6 | 4.6 | 15.3 | 7 | 97.5 | <0.001 |  |
| North-West | 2,428 | 4.1 | 3.3 | 4.9 | 1 | - | - |  |
| South-West | 1,041 | 9.5 | 7.8 | 11.5 | 1 | - | - |  |
| North | 668 | 1.8 | 0.9 | 3.1 | 1 | - | - |  |
| West | 560 | 8.9 | 6.7 | 11.6 | 1 | - | - |  |
| Multi-region | 47,957 | 4.3 | 2.9 | 6.5 | 7 | 97.7 | <0.001 |  |
| **Participant** |  |  |  |  |  |  |  | <0.001 |
| Unspecific tuberculosis | 66,223 | 8.2 | 5.8 | 11.4 | 17 | 98.9 | <0.001 |  |
| Retreated tuberculosis | 753 | 11.8 | 9.6 | 14.3 | 1 | - | - |  |
| Rifampicin-resistant tuberculosis | 1,146 | 3.4 | 2.5 | 4.6 | 3 | 0.0 | 0.752 |  |
| Multidrug-resistant tuberculosis | 150 | 3.3 | 1.1 | 7.6 | 1 | - | - |  |
| Tuberculosis/HIV coinfected | 337 | 4.2 | 2.3 | 6.9 | 1 | - | - |  |
| Pediatric tuberculosis | 2,159 | 5.5 | 1.6 | 16.9 | 3 | 98.4 | <0.001 |  |
| **Tuberculosis disease localization** |  |  |  |  |  |  |  | 0.258 |
| Pulmonary | 3,074 | 6.6 | 4.3 | 9.9 | 9 | 89.9 | <0.001 |  |
| Extrapulmonary | 305 | 5.7 | 1.3 | 22.2 | 2 | 88.0 | 0.004 |  |
| Mixed | 66,074 | 6.9 | 4.3 | 10.7 | 14 | 99.2 | <0.001 |  |
| Not reported | 1,041 | 9.5 | 7.8 | 11.5 | 1 | - | - |  |
| **Treatment regimen<sup>3</sup>** |  |  |  |  |  |  |  | <0.001 |
| Standard regimen first line | 69,028 | 7.9 | 5.6 | 10.9 | 21 | 98.9 | <0.001 |  |
| Second-line regimen | 1,740 | 3.6 | 2.8 | 4.6 | 5 | 98.6 | <0.001 |  |

*1:  Random effects model; 2: Time frame aligns with the development of successive National Strategic Plans against tuberculosis in Cameroon; CI: Confidence interval; Mixed included pulmonary and extrapulmonary tuberculosis cases; HIV: Human immunodeficiency virus*

#### HIV status

People living with HIV were significantly more likely to have unfavorable treatment outcomes (AOR = 2.57; 95% CI: 1.86–3.55; 18 reports).

#### Antiretroviral therapy

Patients lacking antiretroviral therapy were at significantly higher risk of an unfavorable outcome than those receiving it (AOR = 1.71; 95% CI: 1.15–2.54; 3 reports).

#### Sex

Male sex was associated with increased odds of an unfavorable treatment outcome (AOR = 1.67; 95% CI: 1.45–1.92; 13 reports).

#### Age

Patients aged ≤40 years had significantly higher odds of unfavorable treatment outcomes compared with those aged >40 years (AOR = 1.45; 95% CI: 1.06–1.98; 3 reports).

#### Past history of TB treatment

Retreated TB patients were more likely to have an unfavorable outcome than new TB patients (AOR = 1.99; 95% CI: 1.40–2.84; 9 reports) (**Table 6 and Figs. S34-40**)

**Table 6.**
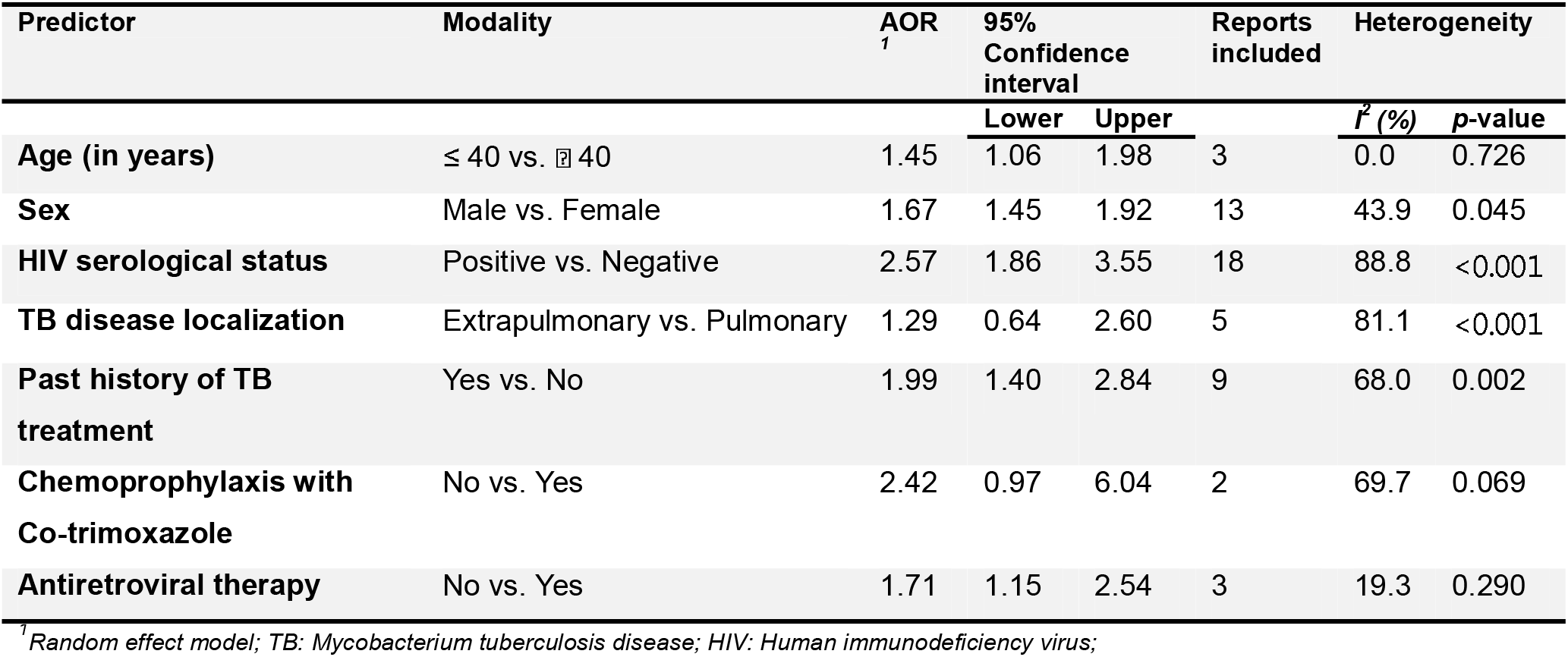
Predictors of unfavorable outcome among patients treated for drug-resistant tuberculosis in Cameroon, 1998–2022.

| **Predictor** | **Modality** | **AOR**<sup>*1*</sup> | **95% Confidence interval** |  | **Reports included** | **Heterogeneity** |  |
|---|---|---|---|---|---|---|---|
|  |  |  | **Lower** | **Upper** |  | ***I***<sup>***2***</sup> ***(%)*** | ***p*-value** |
| **Age (in years)** | ≤ 40 vs. ⍰ 40 | 1.45 | 1.06 | 1.98 | 3 | 0.0 | 0.726 |
| **Sex** | Male vs. Female | 1.67 | 1.45 | 1.92 | 13 | 43.9 | 0.045 |
| **HIV serological status** | Positive vs. Negative | 2.57 | 1.86 | 3.55 | 18 | 88.8 | <0.001 |
| **TB disease localization** | Extrapulmonary vs. Pulmonary | 1.29 | 0.64 | 2.60 | 5 | 81.1 | <0.001 |
| **Past history of TB treatment** | Yes vs. No | 1.99 | 1.40 | 2.84 | 9 | 68.0 | 0.002 |
| **Chemoprophylaxis with Co-trimoxazole** | No vs. Yes | 2.42 | 0.97 | 6.04 | 2 | 69.7 | 0.069 |
| **Antiretroviral therapy** | No vs. Yes | 1.71 | 1.15 | 2.54 | 3 | 19.3 | 0.290 |

<sup>*1*</sup>*Random effect model; TB: Mycobacterium tuberculosis disease; HIV: Human immunodeficiency virus;*

## 4. Discussion

This systematic review and meta-analysis brought together evidence from 32 studies conducted in Cameroon between 1998 and 2022 to examine tuberculosis treatment outcomes and their associated factors. The pooled treatment success rate was 78.2%, while mortality, treatment failure and loss to follow-up were 8.1%, 0.9% and 6.8%, respectively. Although some improvement was observed over the years, the findings suggest that a considerable proportion of patients in Cameroon still experience poor treatment outcomes. Important differences were also observed across regions and patient groups, with HIV coinfection, previous tuberculosis treatment, male sex and absence of antiretroviral therapy emerging as significant factors associated with unfavorable outcomes.

Our finding of a pooled treatment success rate of 78.2% was consistent with findings from previous studies conducted across Africa. For instance, a systematic review and meta-analysis involving 31 studies and 18,194 participants from sub-Saharan Africa reported a treatment success rate of 76.2% [53]. Similar findings were also documented in a meta-analysis of 26 African studies [54], which reported a pooled treatment success rate of 79.0%. Our findings suggest that the challenges affecting tuberculosis treatment in Cameroon are also being experienced in other African countries. However, our estimate was lower than the 83.7% reported in Ethiopia [53] and the global treatment success rate of 88% for drug-susceptible tuberculosis reported by the World Health Organization [55]. Our findings should be interpreted with caution, given differences in study populations and treatment regimens. The results suggest that there is still room to improve tuberculosis treatment outcomes in Cameroon. Differences in access to healthcare, patient follow-up, treatment adherence and the organization of tuberculosis services may help explain some of these variations.

Mortality remains an important concern in this review, with a pooled mortality rate of 8.1% indicating that a substantial proportion of patients die during tuberculosis treatment. The burden was particularly high among patients coinfected with TB and HIV, for whom mortality reached 30.5%. This was considerably higher than the 6.9% observed among patients in the general tuberculosis category. The finding is consistent with previous studies showing that HIV infection remains a major contributor to poor tuberculosis treatment outcomes in sub-Saharan Africa. An Ethiopian study reported that HIV-coinfected patients had nearly twice the odds of unsuccessful treatment compared with HIV-negative patients (OR = 1.98; 95% CI: 1.56–2.52) [56]. Similarly, a more recent meta-analysis found that HIV coinfection increased the odds of unsuccessful tuberculosis treatment by approximately 2.65 times (95% CI: 2.1–3.3) [57]. The consistency of these findings with our results highlights the continuing challenge of managing tuberculosis among people living with HIV and highlights the importance of integrating TB management into routine HIV care.

Several factors may contribute to the high mortality observed among TB/HIV-coinfected patients, particularly advanced immunosuppression, opportunistic infections and delayed diagnosis. A systematic review and meta-analysis conducted in Ethiopia found that patients with CD4 counts below 50 cells/mm³ had a significantly higher risk of mortality (HR = 3.37; 95% CI: 2.18–5.22), while the presence of opportunistic infections was also associated with increased mortality (HR = 1.75; 95% CI: 1.30–2.34) [58]. Similarly, WHO has highlighted the increased risk of death among patients with advanced HIV disease, particularly those presenting with tuberculosis and other opportunistic infections [59]. Patients presenting with advanced HIV disease may already have severe tuberculosis or other infections by the time treatment is initiated, further complicating their management. Although our review could not establish the contribution of these individual factors, the findings highlight the importance of early identification of TB/HIV coinfection and timely initiation of appropriate treatment. In particular, the absence of cotrimoxazole chemoprophylaxis initiation emerged as a significant predictor of poor outcomes among TB-treated patients. Better integration of tuberculosis and HIV services, regular clinical monitoring, and continued access to antiretroviral therapy could therefore help improve survival among these patients, in line with WHO recommendations [59].

Furthermore, the Mortality rate in our study also varied according to the type of tuberculosis. Patients with rifampicin-resistant tuberculosis had a pooled mortality rate of significantly higher than those with multidrug-resistant tuberculosis. Although this difference may seem unexpected, it should be interpreted with caution, as the estimates were derived from a small number of studies involving different patient populations. Patients receiving treatment for multidrug-resistant tuberculosis may benefit from more specialized care and closer clinical follow-up, while differences in disease severity and treatment history may also influence outcomes. These findings should therefore not be taken to suggest that multidrug-resistant tuberculosis carries a lower risk of death. Rather, they highlight the importance of considering patient characteristics and treatment settings when interpreting subgroup differences.

The pooled treatment failure rate was relatively low at 0.9% (95% CI: 0.5–1.4). A significant difference was observed across study periods, with treatment failure declining from 2.9% before 2005 to 0.5% after 2020. This finding is encouraging and may reflect improvements in tuberculosis management, particularly in diagnosis, treatment monitoring and the identification of patients who do not respond adequately to treatment. However, a low recorded failure rate does not necessarily mean that treatment outcomes are optimal. Patients who die or are lost to follow-up may not complete the assessments required to determine treatment failure, potentially leading to underestimation. The higher failure rate observed among patients with pulmonary tuberculosis compared with those with extrapulmonary tuberculosis should also be interpreted cautiously, as monitoring treatment response in extrapulmonary disease can be more challenging. Differences in bacteriological confirmation and outcome assessment may therefore have influenced the reported rates.

Loss to follow-up was another important finding, affecting an estimated 6.8% (95% CI: 5.0–9.1) of patients. Although the rate declined from 14.6% in 2005–2009 to 4.9% in 2015– 2019, it increased slightly to 5.7% from 2020 onwards. The overall pattern suggests that retaining patients in treatment remains an important challenge. Tuberculosis treatment requires sustained engagement with healthcare services, and patients may face difficulties related to transportation, treatment-related costs, competing responsibilities, and the burden of prolonged treatment. These challenges may be particularly important in settings where access to healthcare is limited.

Loss to follow-up was highest among previously treated patients, reaching 11.8%. This finding suggests that patients with a history of tuberculosis treatment may face additional difficulties completing subsequent treatment episodes. Such patients may have previously experienced treatment interruption, drug-related adverse effects, or challenges accessing healthcare. The findings are consistent with those observed in an African study who reported that previously treated tuberculosis patients had a higher risk of unsuccessful treatment than newly diagnosed patients (RR = 1.48; 95% CI: 1.14–1.94) [54]. Our analysis also found lower loss-to-follow-up rates among patients receiving second-line treatment (3.6%) than among those receiving first-line regimens (7.9%). One possible explanation is that patients receiving second-line treatment are more likely to be managed in specialized centres where monitoring and adherence support may be more intensive. However, differences in patient characteristics and treatment settings make it difficult to attribute this finding to the treatment regimens themselves.

Beyond the individual treatment outcomes in our study, the analysis identified several factors associated with unfavorable outcomes. HIV coinfection was one of the strongest, with HIV-positive patients having more than twice the odds of an unfavorable outcome compared with HIV-negative patients (AOR = 2.57; 95% CI: 1.86–3.55). This estimate was close to the findings from Ethiopia, where HIV coinfection was associated with an odds ratio of 2.65 [57]. The similarity between these findings strengthens the evidence that HIV remains an important determinant of tuberculosis treatment outcomes across African settings. In addition, patients who were not receiving antiretroviral therapy had higher odds of unfavorable outcomes (AOR = 1.71; 95% CI: 1.15–2.54). Together, these results reinforce the importance of early HIV diagnosis, timely initiation of antiretroviral therapy and closer follow-up of patients with TB/HIV coinfection.

Previous tuberculosis treatment was also significantly associated with unfavorable outcomes (AOR = 1.99; 95% CI: 1.40–2.84). This finding corroborated findings from an Ethiopian meta-analysis which reported approximately twice the odds of unsuccessful outcomes among previously treated patients (OR = 2.17; 95% CI: 1.55–3.03) [56]. Patients with a previous history of tuberculosis may be more likely to have underlying drug resistance, recurrent disease or difficulties with treatment adherence [13]. Although these explanations require further investigation, the findings suggest that previously treated patients should receive particular attention during treatment assessment and follow-up.

Male sex was another factor associated with unfavorable outcomes (AOR = 1.67; 95% CI: 1.45–1.92). Differences in health-seeking behavior, work-related mobility, social circumstances and treatment adherence may partly explain this association. In some settings, men may delay seeking care or encounter difficulties attending regular treatment appointments because of occupational or financial responsibilities. However, these explanations were not directly assessed in the present review and should be explored in future studies. Understanding the barriers experienced by different patient groups could help tuberculosis programmes provide more appropriate and targeted support.

Treatment outcomes also varied across geographical regions, with significant differences observed in treatment success and mortality. However, these findings should be interpreted with caution, as some regions were represented by only one or two studies, which limits the reliability of regional comparisons. More geographically representative studies are needed to better understand variations in tuberculosis treatment outcomes across Cameroon.

Finally, substantial heterogeneity was observed across the pooled estimates. Similar findings have been reported in previous African systematic reviews [53, 54], suggesting that tuberculosis treatment outcomes vary considerably between populations and healthcare settings. Differences in study design, treatment regimens, HIV status, disease presentation and outcome reporting may have contributed to this variation. Although the pooled estimates provide an overview of the available evidence, they should be interpreted in light of these differences rather than as uniform outcomes across all tuberculosis treatment settings in Cameroon.

## 5. Limitations

This study has several limitations. First, considerable heterogeneity was observed across the included studies, probably reflecting differences in patient characteristics, study settings, treatment regimens and periods of data collection. Although subgroup analyses were conducted to explore these differences, they did not fully explain the observed variation. The inclusion of different tuberculosis populations, including patients with drug-resistant TB and those coinfected with HIV, may also have influenced the pooled estimates. Another limitation was the uneven geographical distribution of the included studies. Some regions of Cameroon were represented by only one study, while others had no available data. This limits the extent to which the findings can be generalized to the entire country. Similarly, some subgroup analyses were based on relatively few studies, making their estimates less precise.

Most of the included studies were observational, which limits our ability to draw causal conclusions about the factors associated with treatment outcomes. Differences in how treatment outcomes were reported, along with incomplete data in some studies, may also have affected the results. Furthermore, funnel plot asymmetry and the significant Egger’s test for treatment success suggest possible small-study effects or publication bias. Despite these limitations, this review brings together available evidence on tuberculosis treatment outcomes in Cameroon and identifies important areas where patient care and follow-up could be improved.

## 6. Conclusions

This systematic review and meta-analysis found that tuberculosis treatment outcomes in Cameroon remain suboptimal, with a pooled treatment success rate of 78.2%, mortality of 8.1%, treatment failure of 0.9% and loss to follow-up of 6.8%. Although some improvements were observed across study periods, considerable differences remained between patient groups and study settings. HIV coinfection, previous tuberculosis treatment, male sex and absence of antiretroviral therapy were associated with increased odds of unfavorable outcomes. These findings highlight the need to strengthen tuberculosis care in Cameroon, particularly through better integration of TB and HIV services, closer patient follow-up and improved treatment support for those at greater risk of poor outcomes. Addressing barriers to treatment completion, strengthening routine surveillance and improving the quality of care across different regions could help reduce preventable deaths and improve tuberculosis treatment outcomes in Cameroon.

## Supplementary Information

Supplementary Material

## Acknowledgments

Not applicable.

## Authors’ contributions

FZLC conceived the original idea of the study. FZLC, RT, JTZ conducted the literature search. FZLC, RT, JTZ, JTZ, CA selected the studies, extracted the relevant information, critically assessed included reports and synthesized the data. FZLC performed the analyses. FZLC, JTZ, and CA wrote the first draft of the manuscript. FZLC, JTZ, CA, RT, CJKM, ATT, and CAM critically reviewed and revised successive drafts of the manuscript. FZLC, JTZ, CA, RT, CJKM, ATT, and CAM read and approved the final version of the manuscript.

## Data availability

All data generated in this study are available in the main manuscript and its supplementary material.

## Declarations

### Ethical approval and consent to participate

Not applicable.

### Consent for publication

Not applicable.

### Clinical trial number

Not applicable.

## Authors’ details

^1^Department of Public Health, Faculty of Medicine and Biomedical Sciences, The University of Yaoundé 1, Yaoundé, Cameroon

^2^Regional Delegation of Public Health, West Region, Bafoussam, Cameroon

^3^Department of Global Health and Bioethics, Euclid University, Bangui, Central African Republic

^4^Social Epidemiology Lab e.V, Wuppertal, Germany

^5^Central Africa University Institute of Excellence, Yaoundé, Cameroon

^6^Department of Public Health, Faculty of Medical Sciences, University of West Indies, Bridgetown, Barbados

^7^School of Biosciences, University of Kent, Canterbury, UK

^8^Faculty of Sciences and Technologies, Evangelical University Institute of Cameroon, Bandjoun, Cameroon

^9^Department of Public Health and Social Sciences, Faculty of Medicine and Pharmaceutical Sciences of Sangmelima, University of Ebolowa, Ebolowa, Cameroon

## Abbreviations

AOR: Adjusted odds ratio
AJOL: African Journals Online
CI: Confidence interval
EPTB: Extrapulmonary tuberculosis
GLMM: Generalized linear mixed model
HIV: Human immunodeficiency virus
JBI: Joanna Briggs Institute
LTFU: Loss to follow-up
MDR-TB: Multidrug-resistant tuberculosis
MeSH: Medical Subject Headings
PRISMA: Preferred Reporting Items for Systematic Reviews and Meta-Analyses
PROSPERO: International Prospective Register of Systematic Reviews
PTB: Pulmonary tuberculosis
RR-TB: Rifampicin-resistant tuberculosis
TB: Tuberculosis
WHO: World Health Organization

