## Supplementary Material for "Tuberculosis treatment outcome in Cameroon: a systematic review and meta-analysis"

**Tables S1** Search strategy by databases

| **Database** | **Search string** | **Number of entries** |
| --- | --- | --- |
| **Pubmed** | ("tuberculosis"[tiab] OR "TB"[tiab] OR "Mycobacterium tuberculosis"[tiab]) AND ("treatment outcome"[tiab] OR "treatment success"[tiab] OR "cure"[tiab] OR "treatment completion"[tiab] OR "mortality"[tiab] OR "death"[tiab] OR "loss to follow-up"[tiab] OR "default"[tiab] OR "treatment failure"[tiab]) AND ("Cameroon"[tiab]) | 92 |
| **Scopus** | TITLE-ABS-KEY("tuberculosis" OR "TB" OR "Mycobacterium tuberculosis") AND TITLE-ABS-KEY("treatment outcome" OR "treatment success" OR "cure" OR "treatment completion" OR "mortality" OR "death" OR "loss to follow-up" OR "treatment failure") AND TITLE-ABS-KEY("Cameroon") | 156 |
| **Web of sciences** | (((((TS = tuberculosis) OR (TS = TB)) OR (TS = Mycobacterium tuberculosis)) AND (((((((((TS = treatment outcome) OR (TS = treatment success)) OR (TS = cure)) OR (TS = treatment completion)) OR (TS = mortality)) OR (TS = death)) OR (TS = loss to follow-up)) OR (TS = default)) OR (TS = treatment failure))) AND (TS = Cameroon)) | 222 |
| **Embase** | ('tuberculosis'/exp OR'Mycobacterium tuberculosis'/exp OR  tuberculosis:ti,ab,kw OR TB:ti,ab,kw OR'Mycobacterium tuberculosis':ti,ab,kw) AND('treatment outcome'/exp OR  'treatment success'/exp OR 'treatment failure'/exp OR  'cure'/exp OR'mortality'/exp OR'death'/exp OR'lost to follow up'/exp OR 'treatment outcome*':ti,ab,kw OR 'treatment success*':ti,ab,kw OR cure*:ti,ab,kw OR 'treatment completion*':ti,ab,kw OR  'treatment failure*':ti,ab,kw OR'lost to follow-up':ti,ab,kw OR  'lost to follow up':ti,ab,kw ORdefault*:ti,ab,kw OR  mortality:ti,ab,kw OR death*:ti,ab,kw)  AND ('cameroon'/exp OR Cameroon:ti,ab,kw OR  Cameroun:ti,ab,kw OR Cameroonian*:ti,ab,kw) | 239 |
| **Cochrane Library** | (tuberculosis OR TB OR Mycobacterium tuberculosis) AND (treatment outcome OR treatment success OR cure OR treatment completion OR mortality OR death OR loss to follow-up OR default OR treatment failure) AND (Cameroon) | 69 |
| **AJOL** | (tuberculosis OR TB OR Mycobacterium tuberculosis) AND (treatment outcome OR treatment success OR cure OR treatment completion OR mortality OR death OR loss to follow-up OR default OR treatment failure) AND (Cameroon) | 2 |

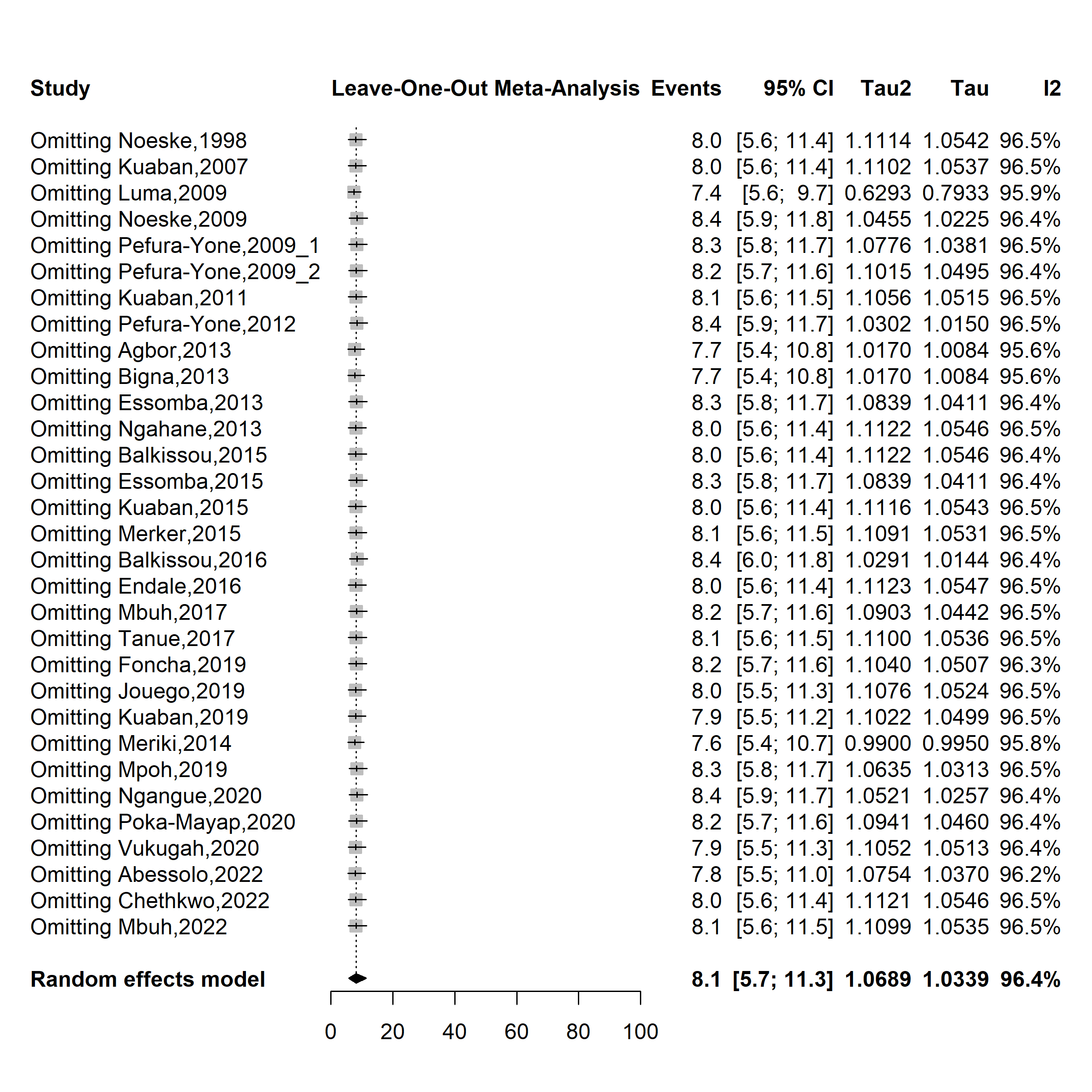

**Fig. S1** Forest plot displaying the sensitivity analysis of the pooled mortality rate among tuberculosis patients in Cameroon, 1998-2022

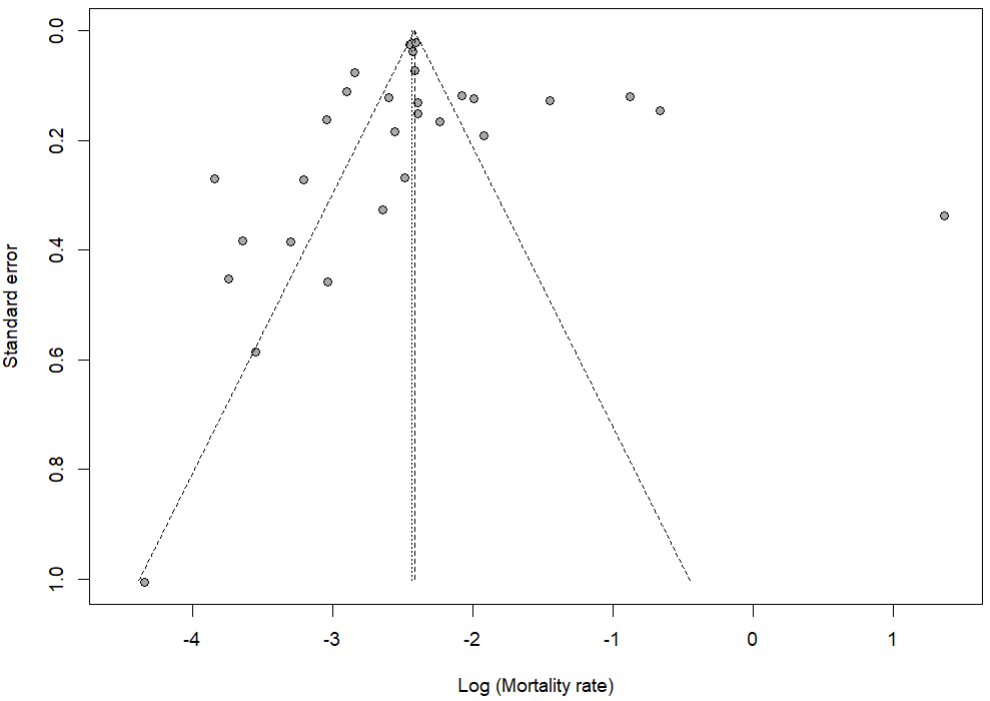

Egger’s test *p*-value = 0.543

Begg’s test *p*-value = 0.424

**Fig. S2** Funnel plot assessing publication bias among studies reporting mortality rates among tuberculosis patients in Cameroon, 1998–2022

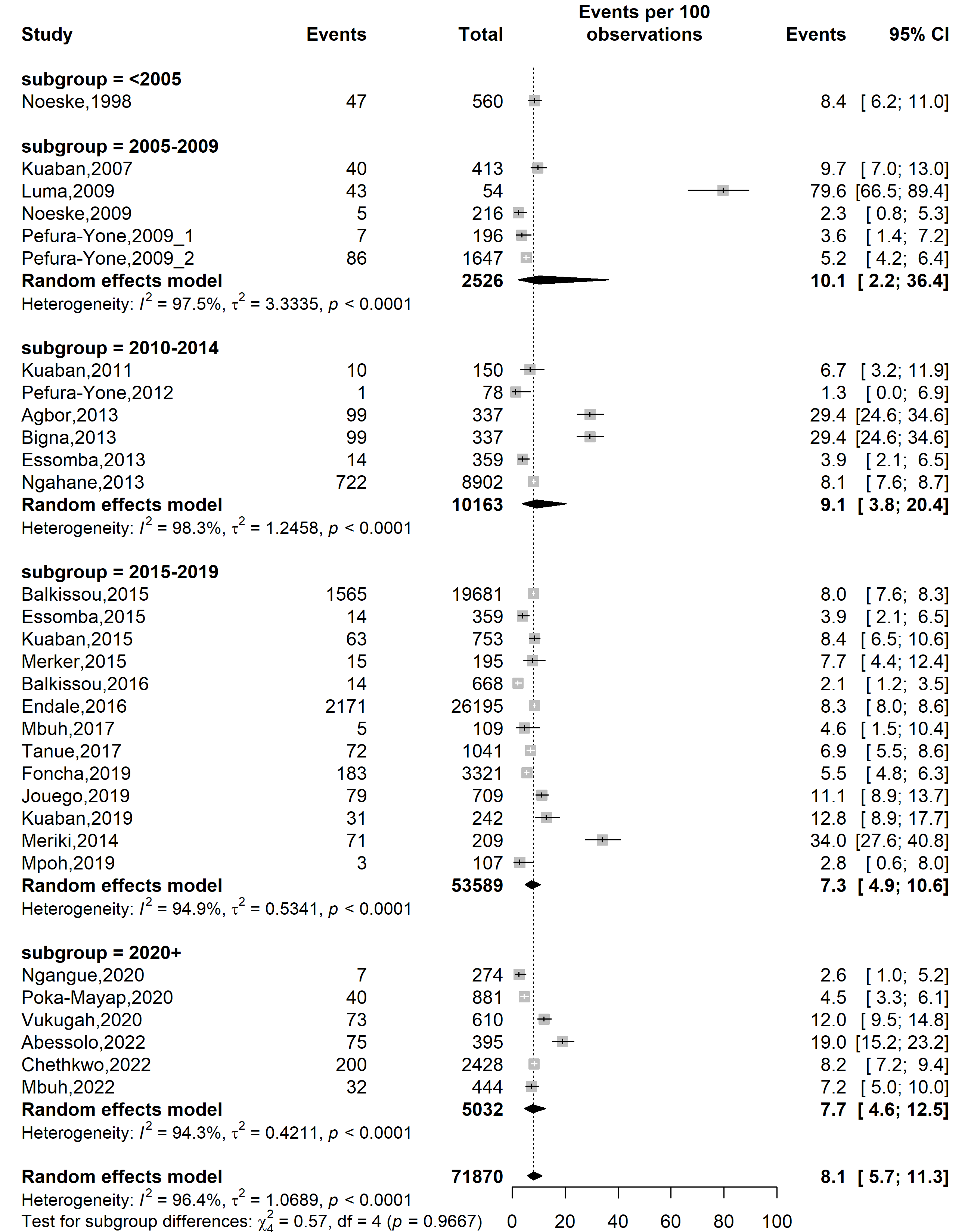

**Fig. S3** Forest plot displaying the subgroup meta-analysis of the pooled mortality rate among tuberculosis patients in Cameroon by study timeframe, 1998-2022

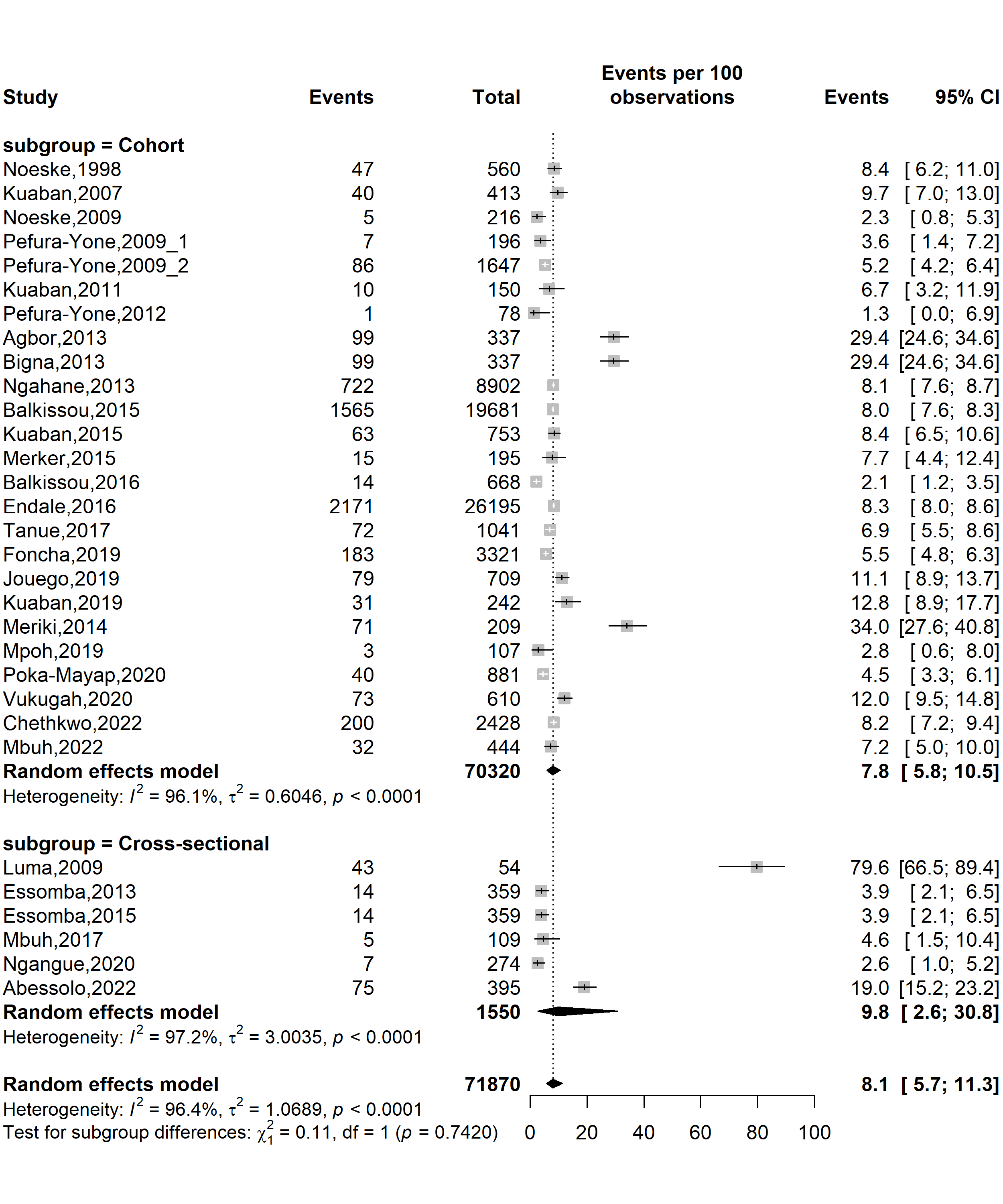

**Fig. S4** Forest plot displaying the subgroup meta-analysis of the pooled mortality rate among tuberculosis patients in Cameroon by study designs, 1998-2022

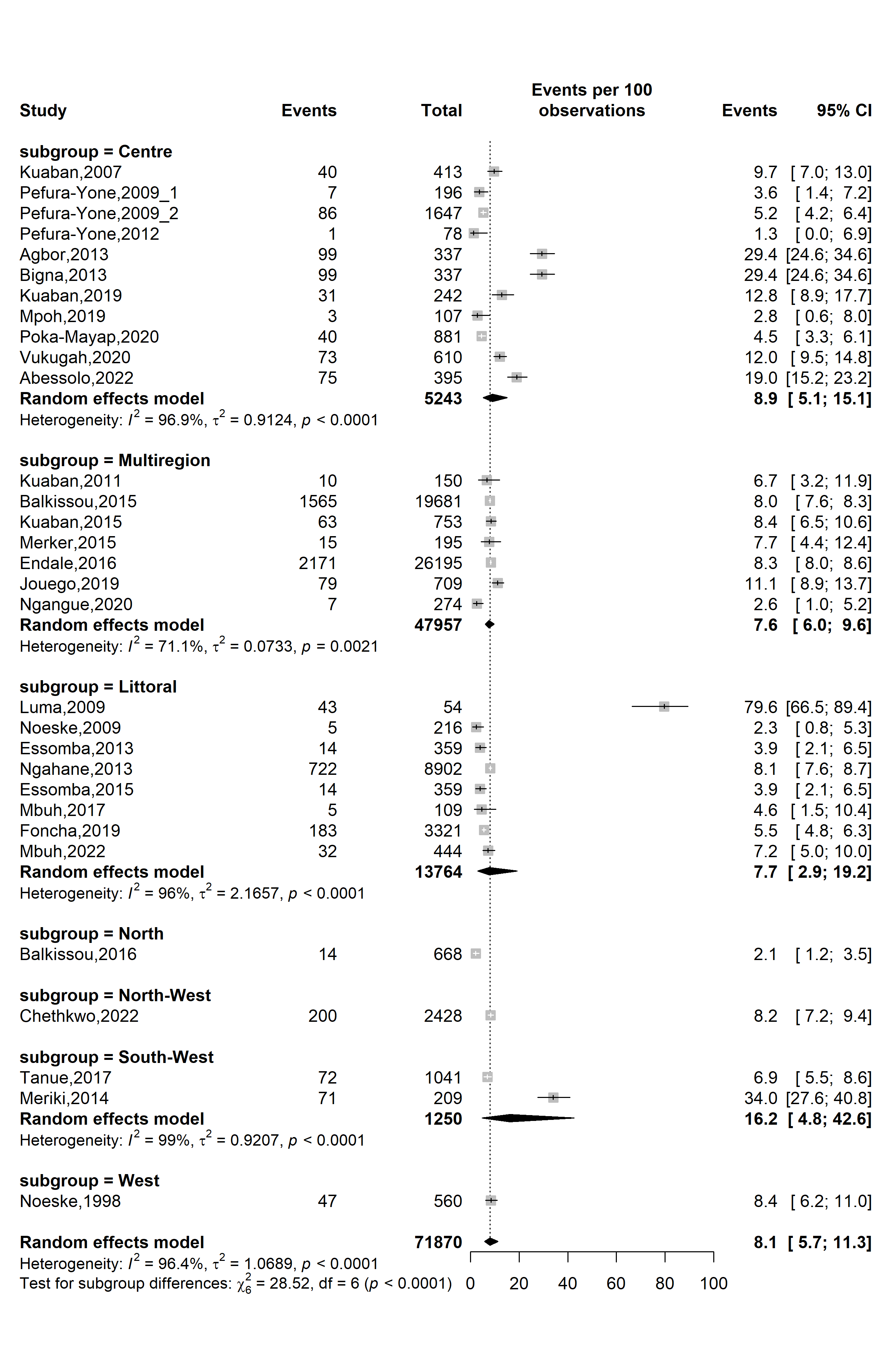

**Fig. S5** Forest plot displaying the subgroup meta-analysis of the pooled mortality rate among tuberculosis patients in Cameroon by study regions, 1998-2022

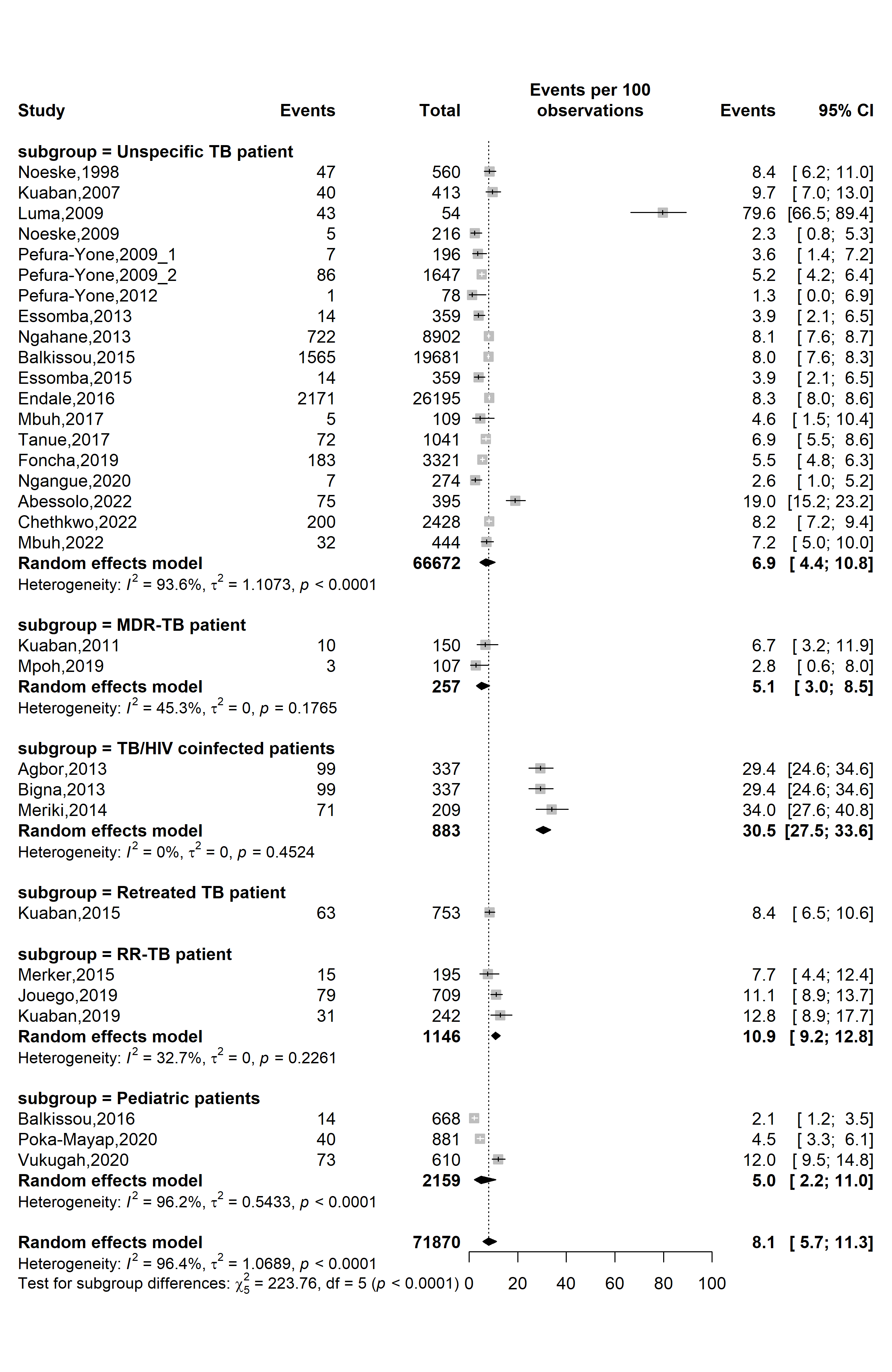

**Fig. S6** Forest plot displaying the subgroup meta-analysis of the pooled mortality rate among tuberculosis patients in Cameroon by types of patients, 1998-2022

*(TB: Mycobacterium tuberculosis infection; RR-TB: Rifampicin-resistant tuberculosis; MDR-TB: Multidrug-resistant tuberculosis; HIV: Human immunodeficiency virus)*

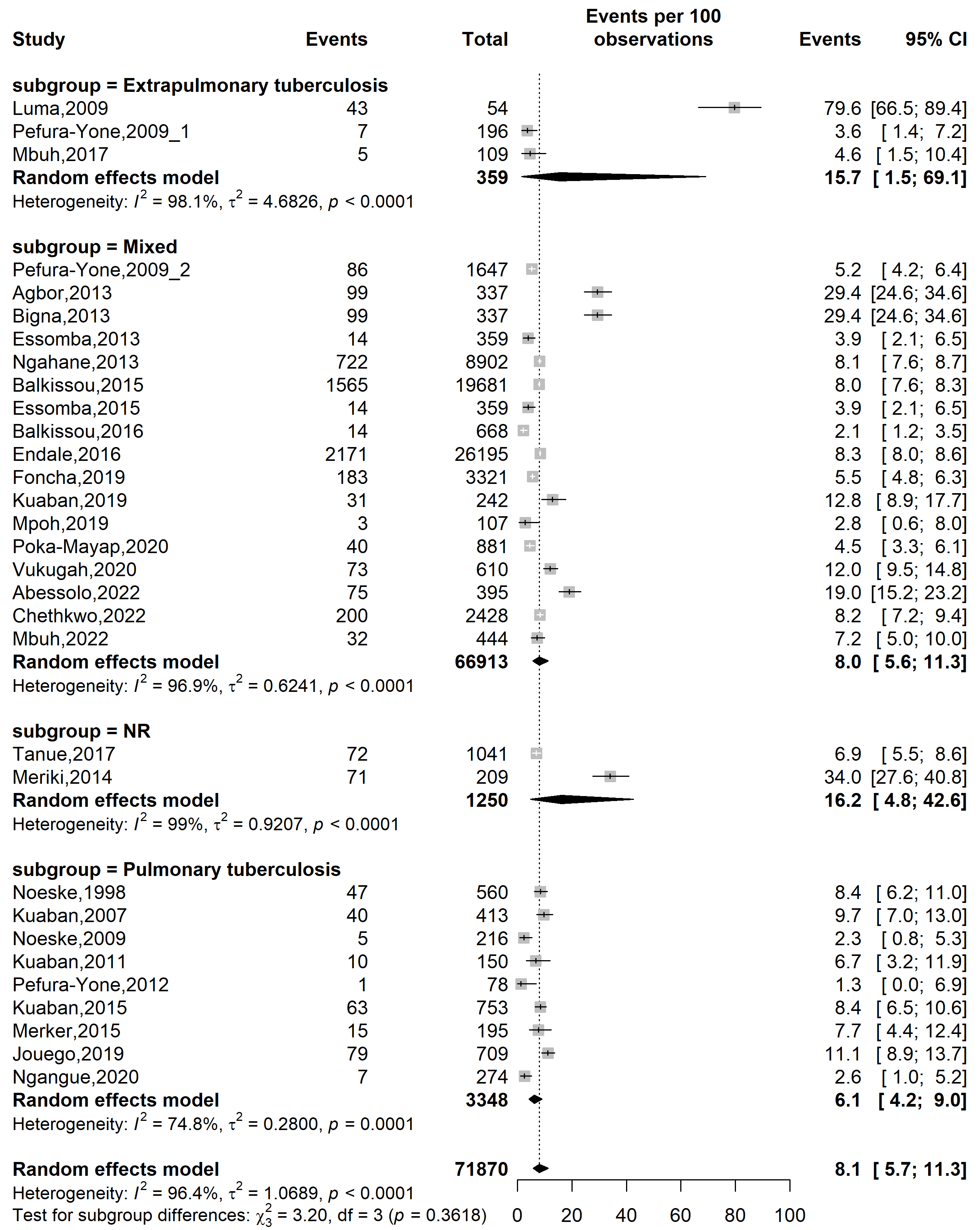

**Fig. S7** Forest plot displaying the subgroup meta-analysis of the pooled mortality rate among tuberculosis patients in Cameroon by tuberculosis disease localizations, 1998-2022

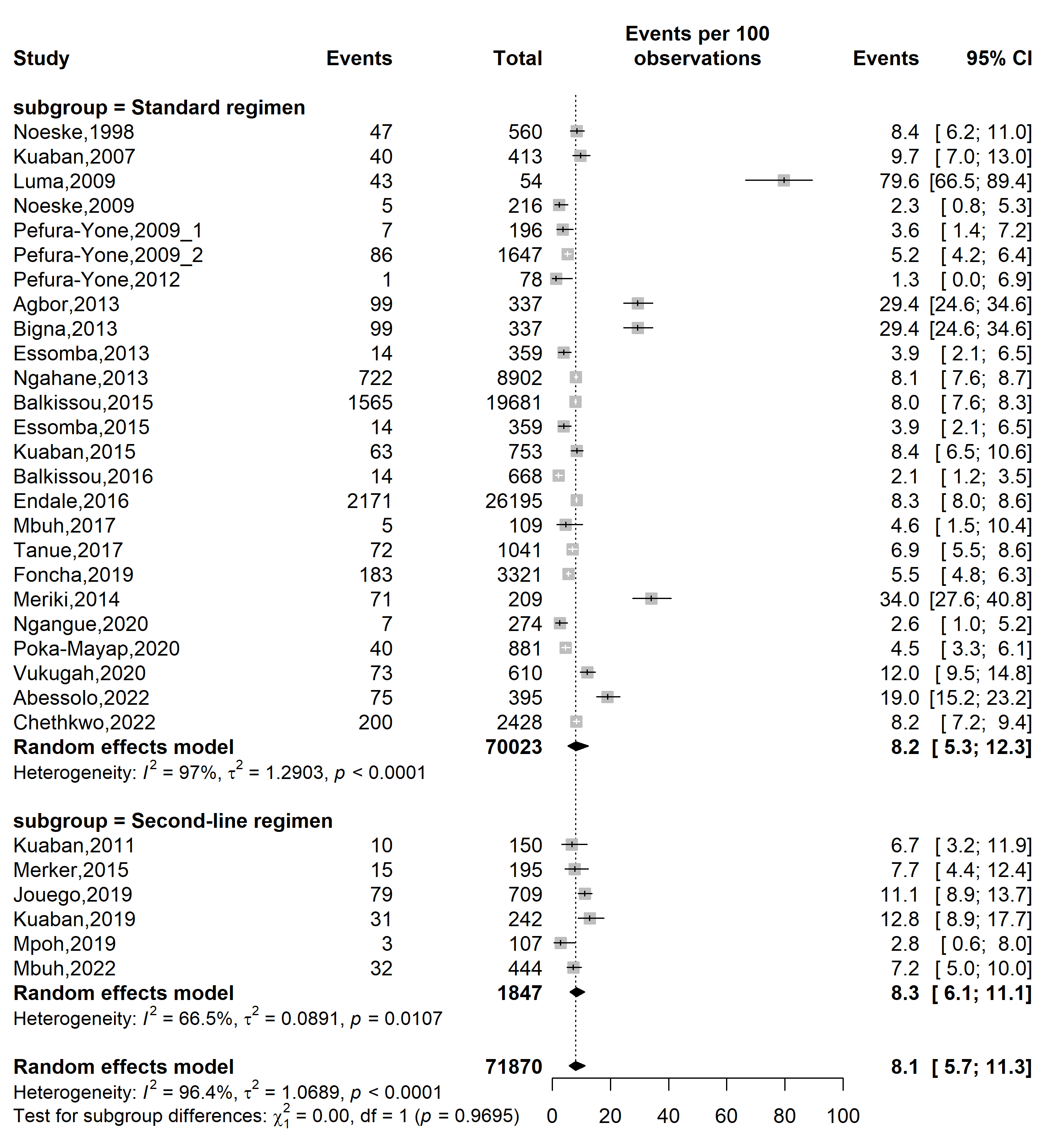

**Fig. S8** Forest plot displaying the subgroup meta-analysis of the pooled mortality rate among tuberculosis patients in Cameroon by types of tuberculosis treatment regimens, 1998-2022

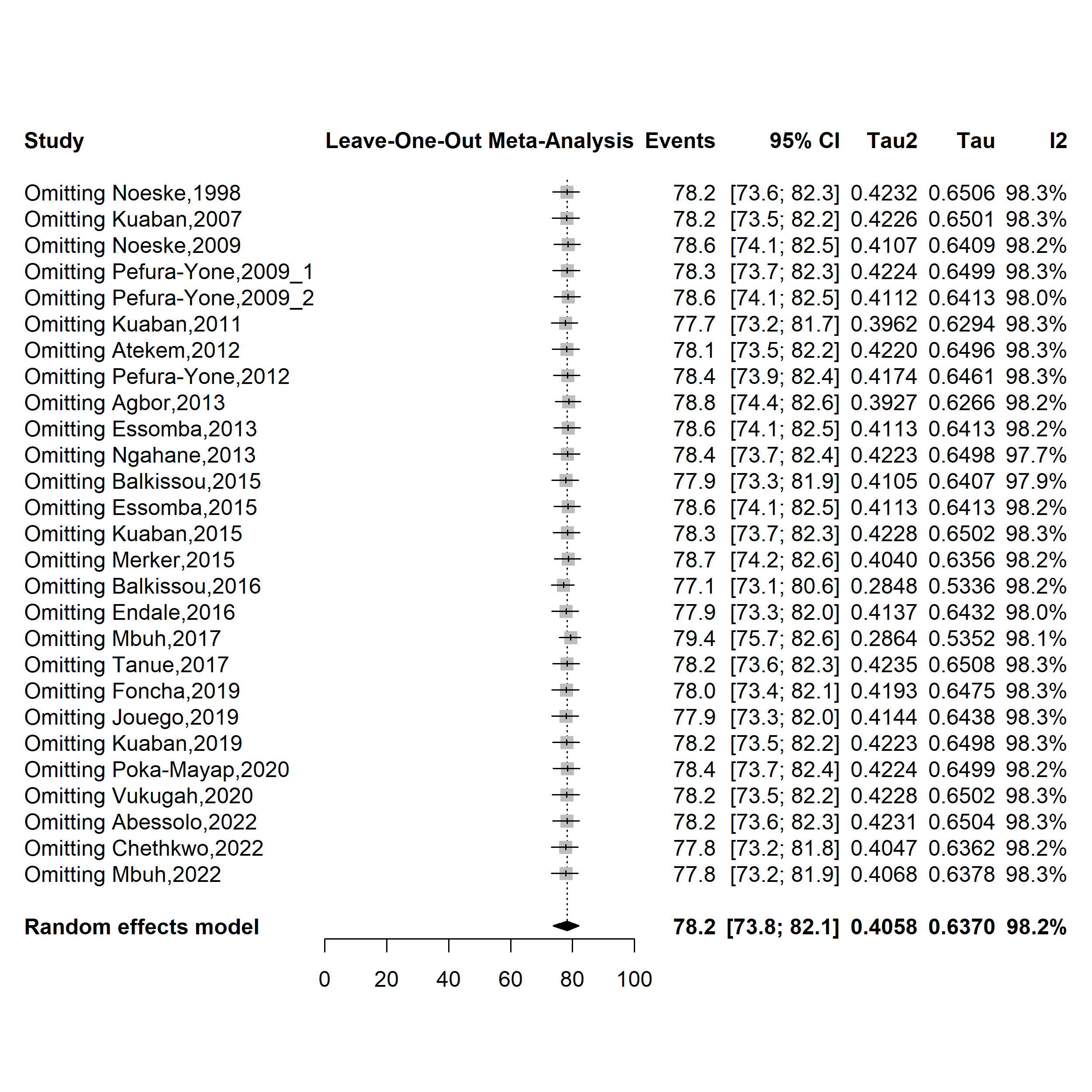

**Fig. S9** Forest plot displaying the sensitivity analysis of the pooled treatment success rate among tuberculosis patients in Cameroon, 1998-2022

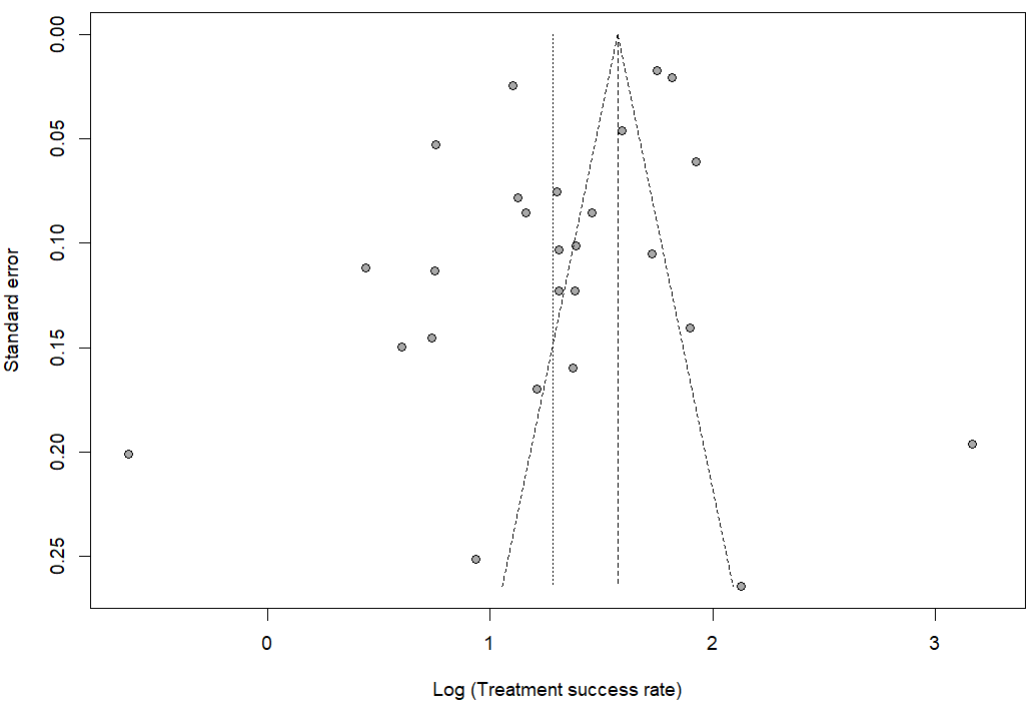

Egger’s test *p*-value = 0.036

Begg’s test *p*-value = 0.770

**Fig. S10** Funnel plot assessing publication bias among studies reporting treatment success rate among tuberculosis patients in Cameroon, 1998–2022

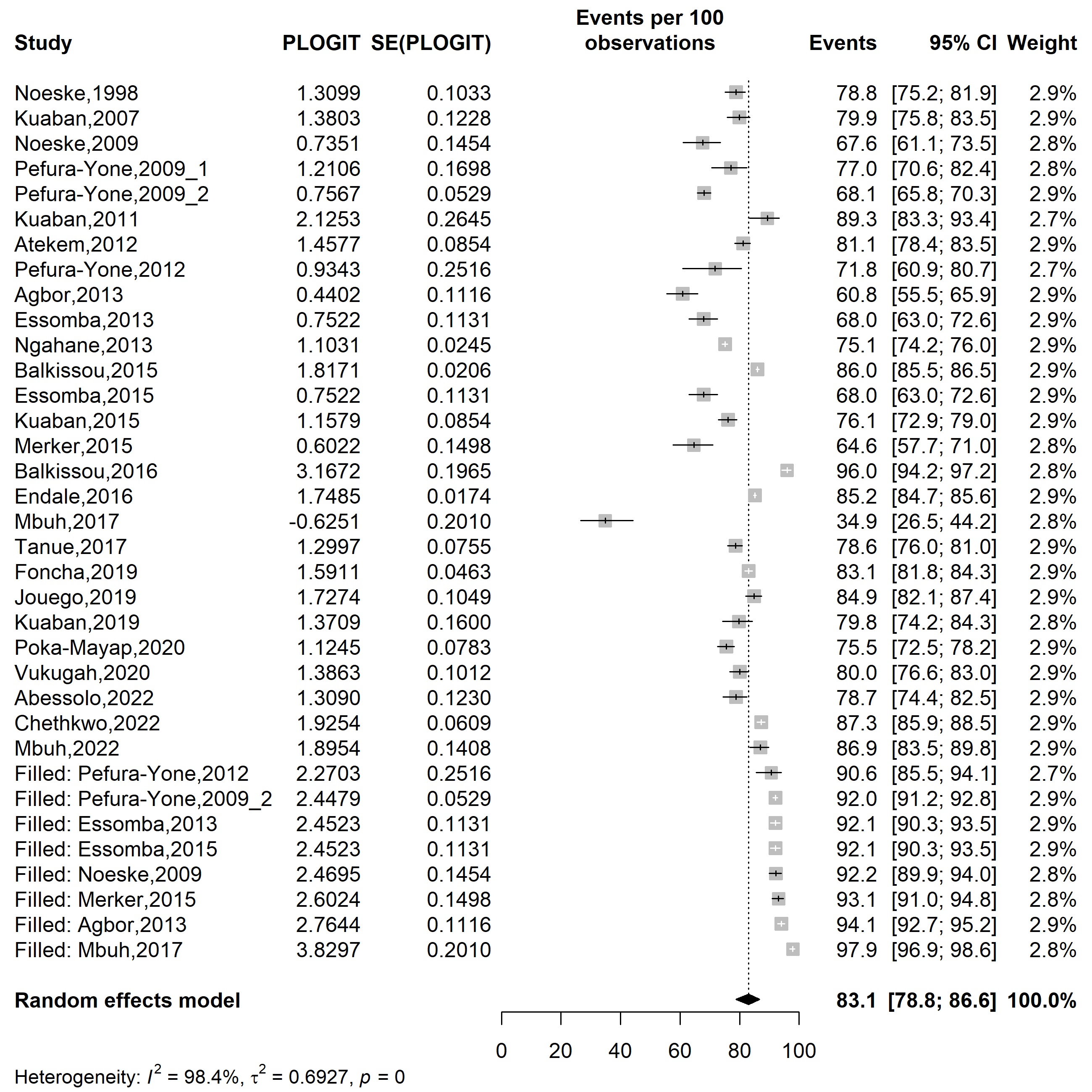

**Fig. S11** Trim and fill plot assessing publication bias among studies reporting treatment success rate among tuberculosis patients in Cameroon, 1998–2022

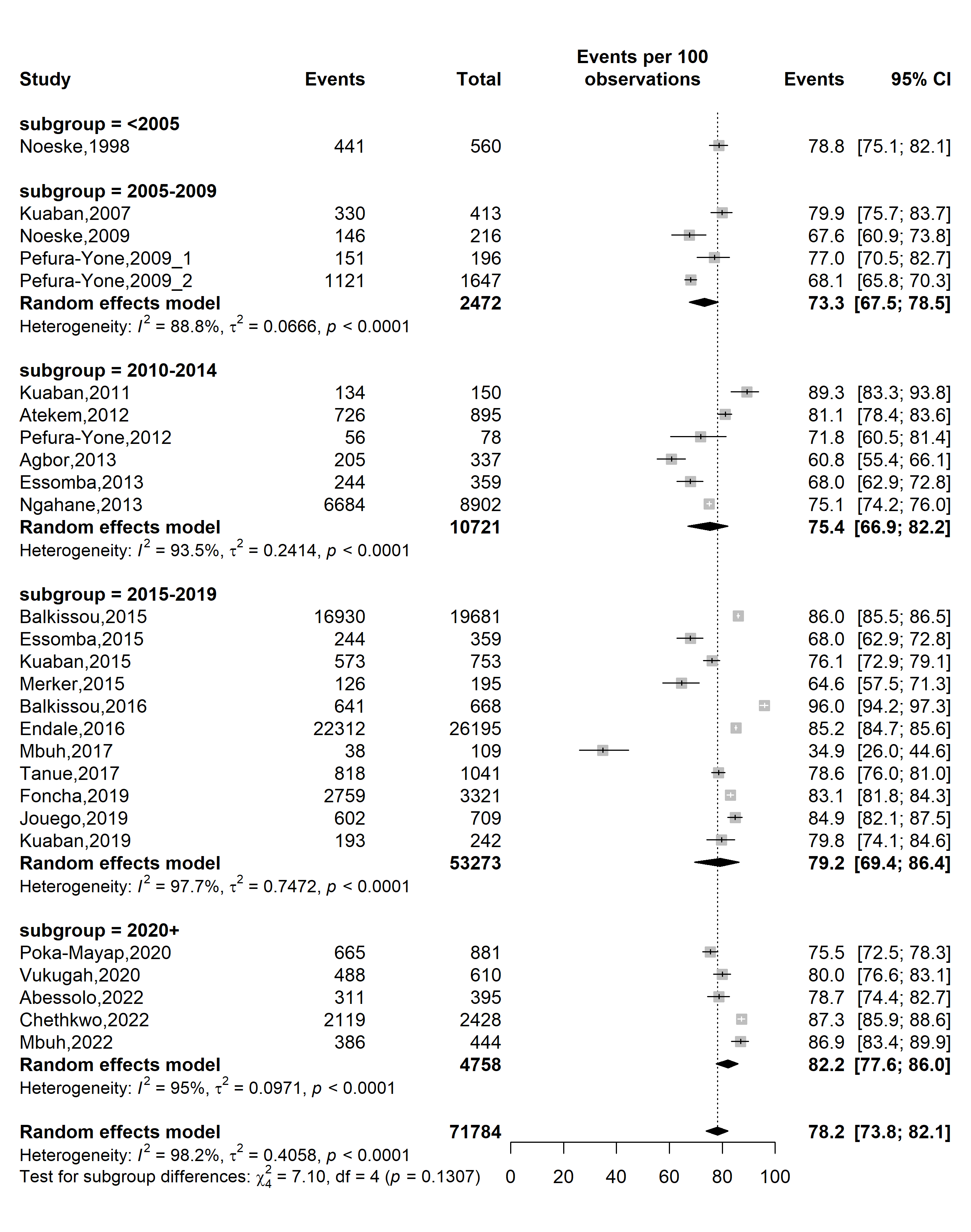

**Fig. S12** Forest plot displaying the subgroup meta-analysis of the pooled treatment success rate among tuberculosis patients in Cameroon by study timeframe, 1998-2022

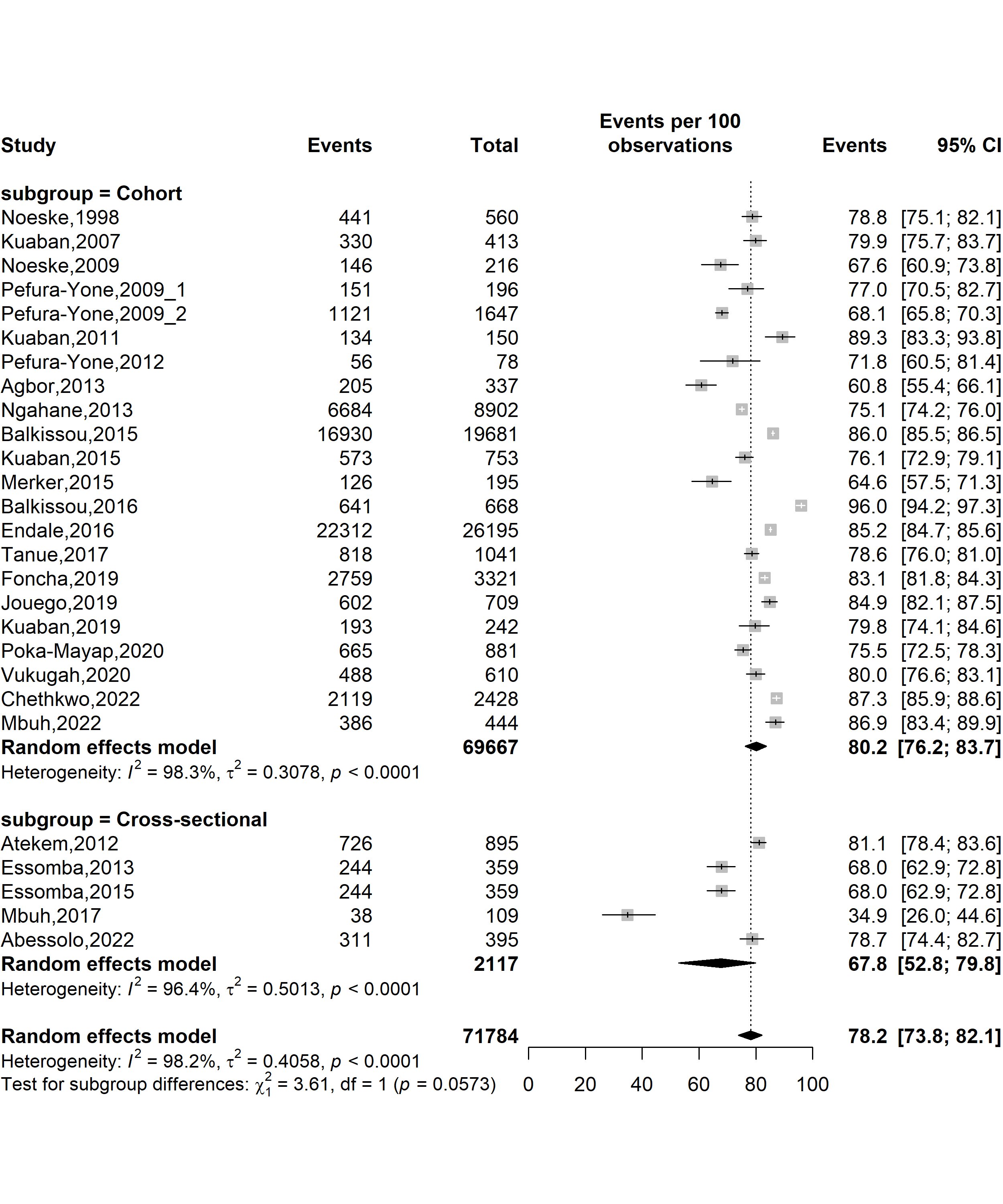

**Fig. S13** Forest plot displaying the subgroup meta-analysis of the pooled treatment success rate among tuberculosis patients in Cameroon by study designs, 1998-2022

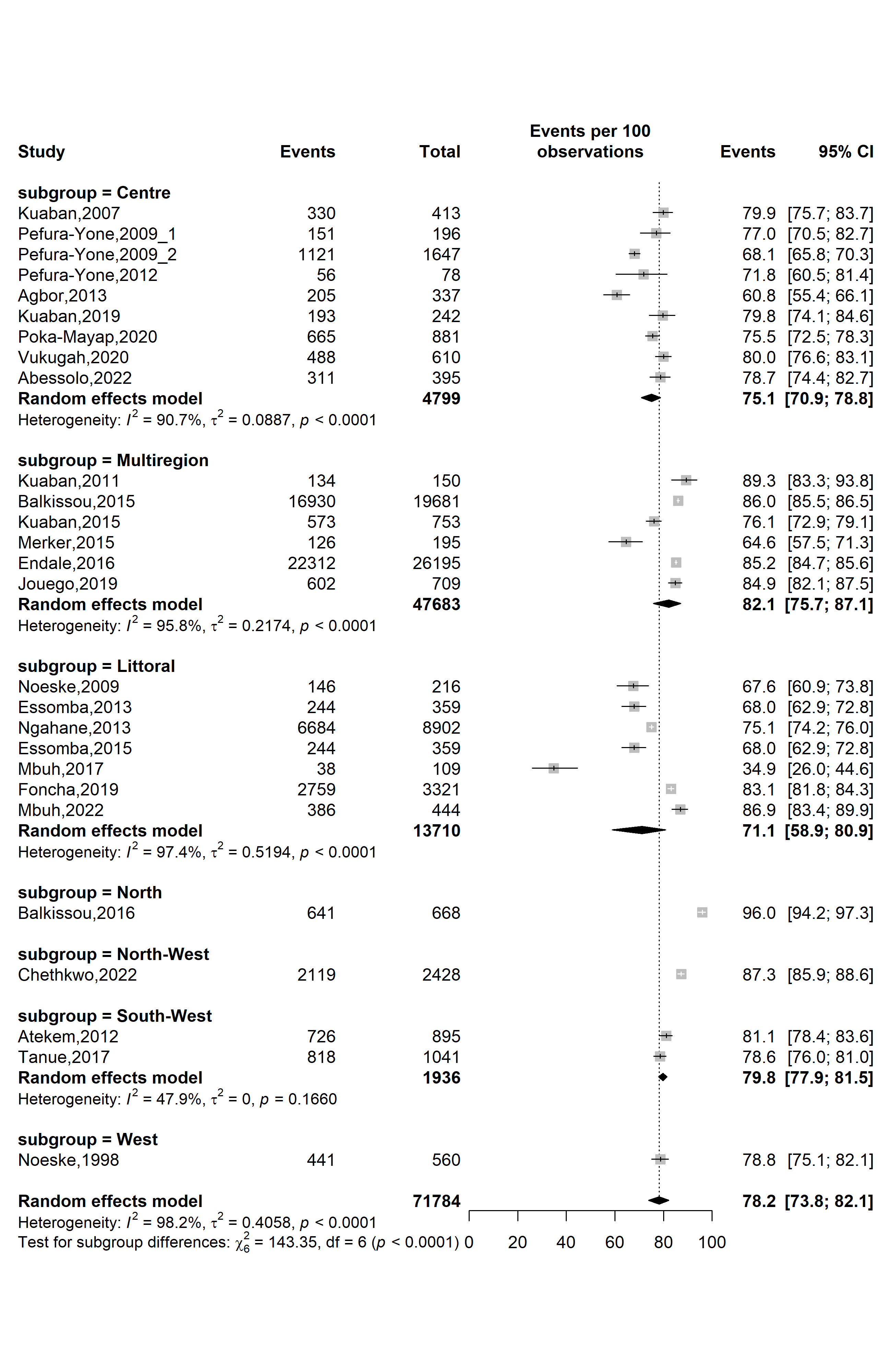

**Fig. S14** Forest plot displaying the subgroup meta-analysis of the pooled treatment success rate among tuberculosis patients in Cameroon by study regions, 1998-2022

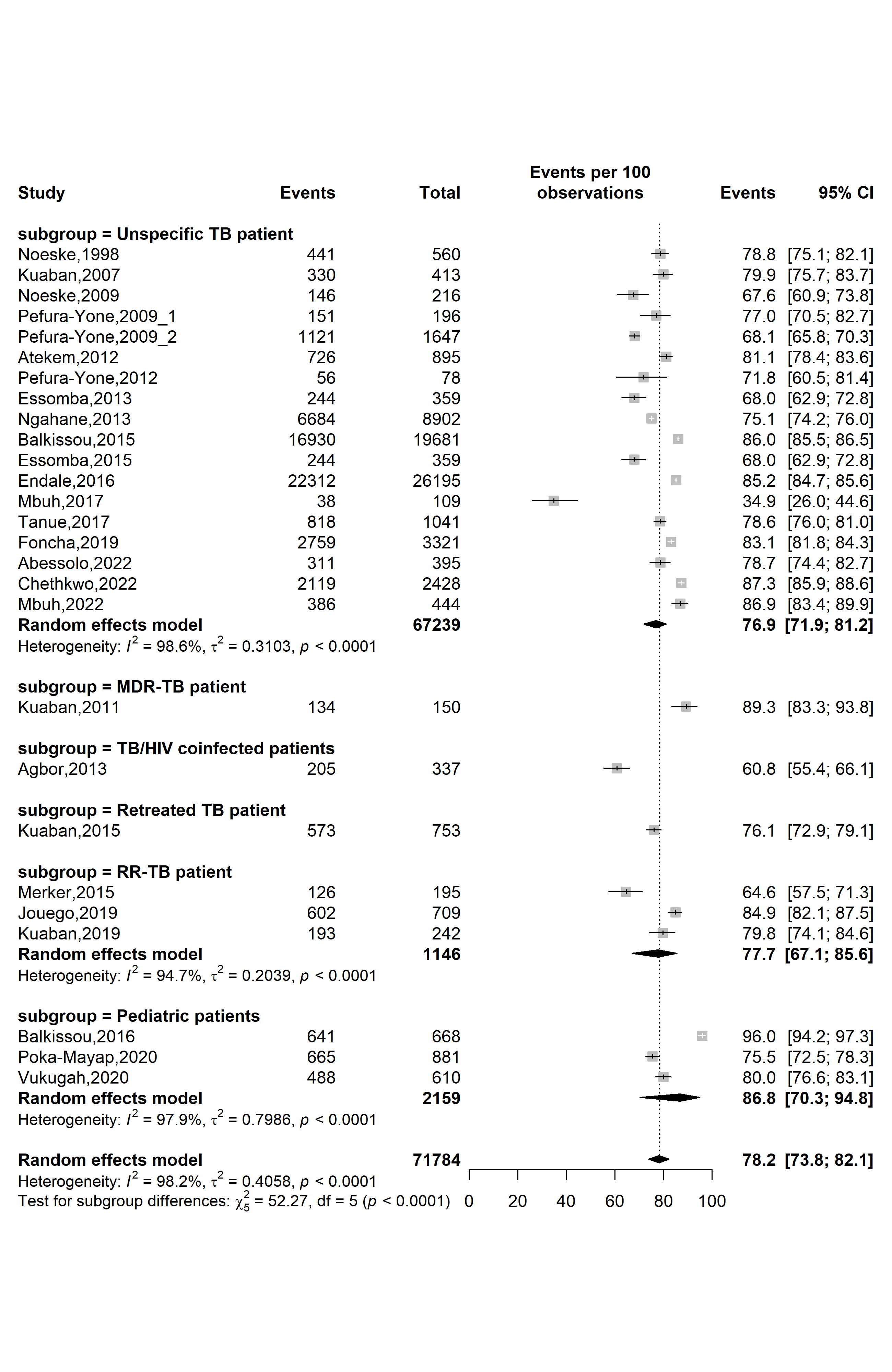

**Fig. S15** Forest plot displaying the subgroup meta-analysis of the pooled treatment success rate among tuberculosis patients in Cameroon by types of patients, 1998-2022

*(TB: Mycobacterium tuberculosis infection; RR-TB: Rifampicin-resistant tuberculosis; MDR-TB: Multidrug-resistant tuberculosis; HIV: Human immunodeficiency virus)*

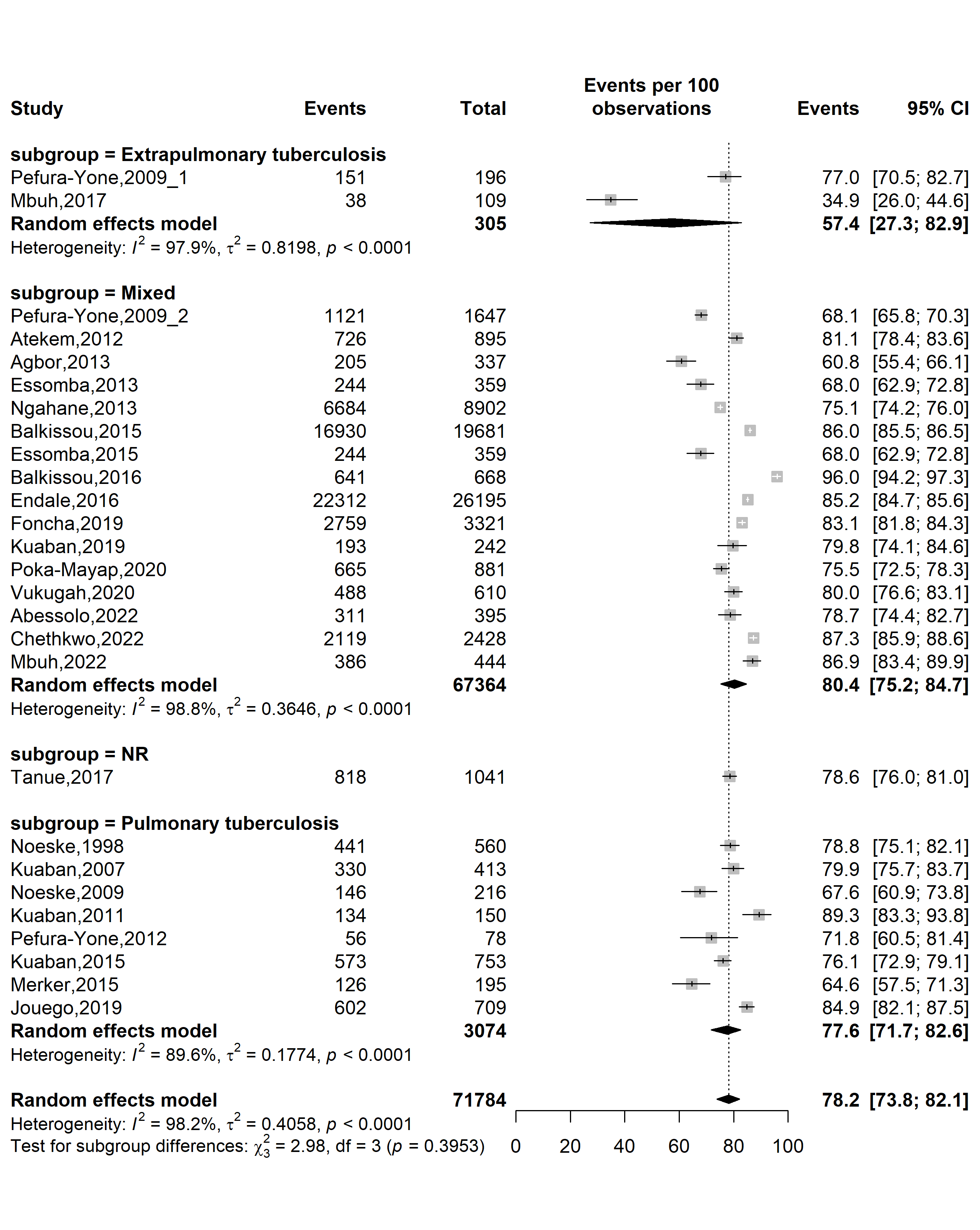

**Fig. S16** Forest plot displaying the subgroup meta-analysis of the pooled treatment success rate among tuberculosis patients in Cameroon by tuberculosis disease localizations, 1998-2022

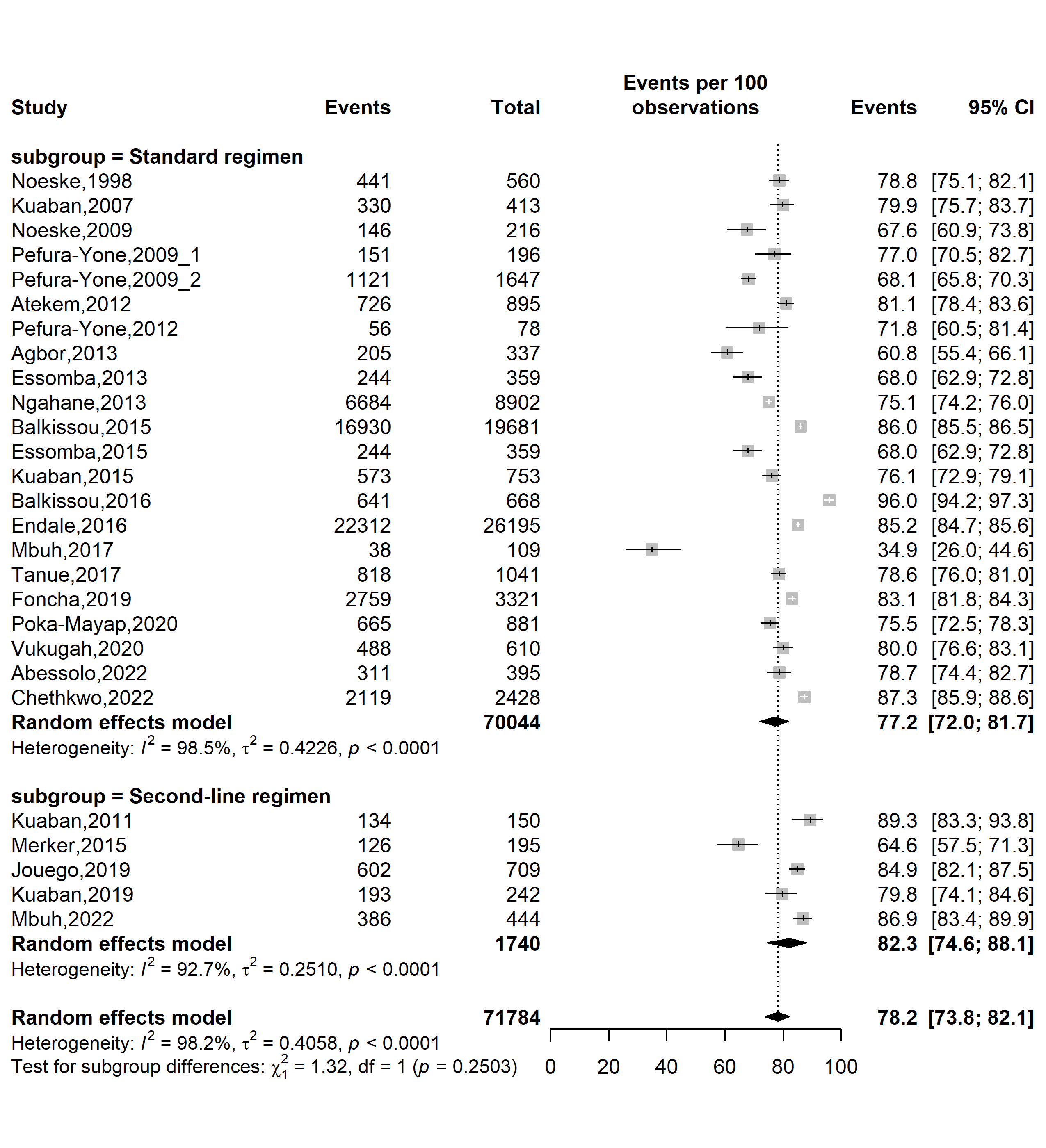

**Fig. S17** Forest plot displaying the subgroup meta-analysis of the pooled treatment success rate among tuberculosis patients in Cameroon by types of resistant tuberculosis regimens, 1998-2022

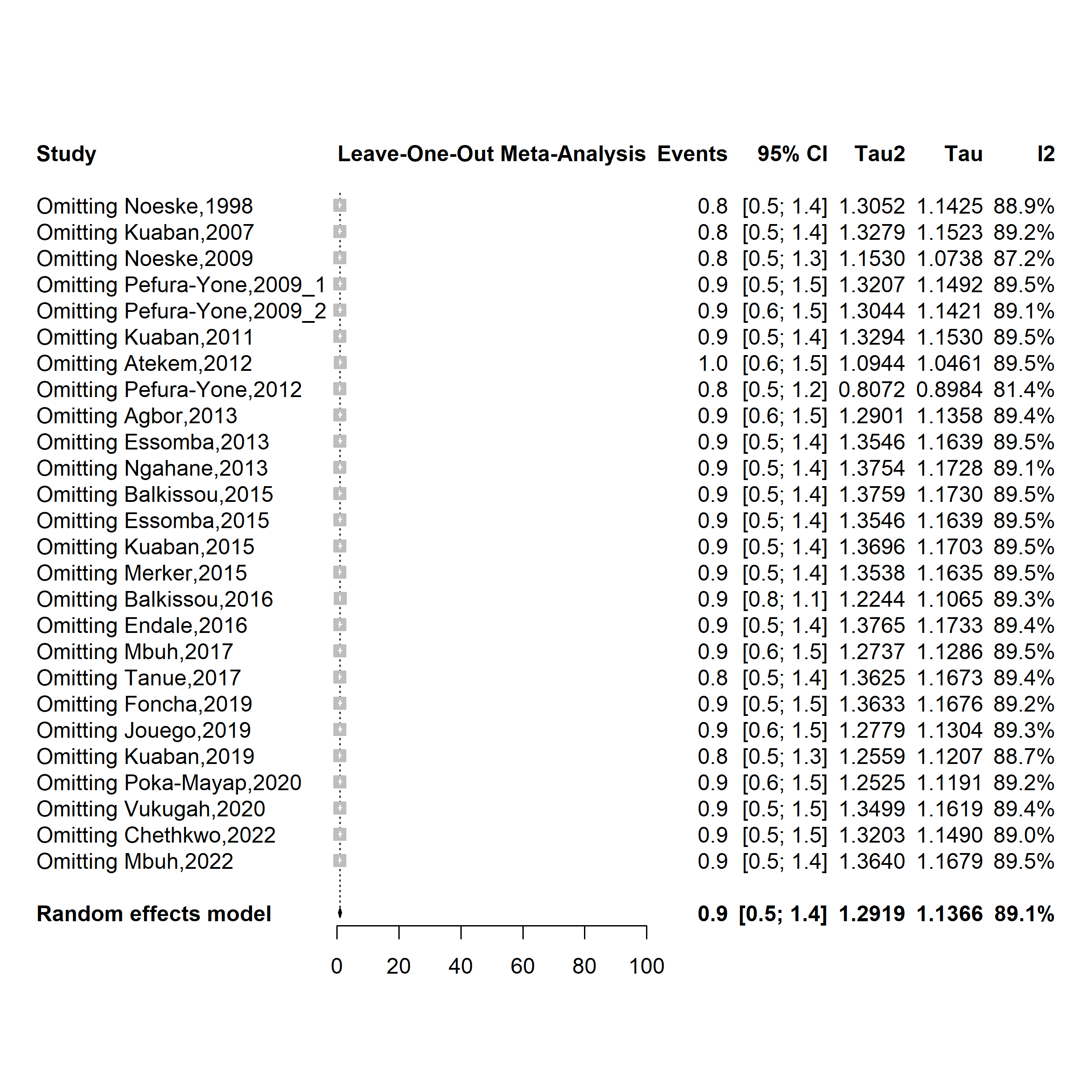

**Fig. S18** Forest plot displaying the sensitivity analysis of the pooled treatment failure rate among tuberculosis patients in Cameroon, 1998-2022

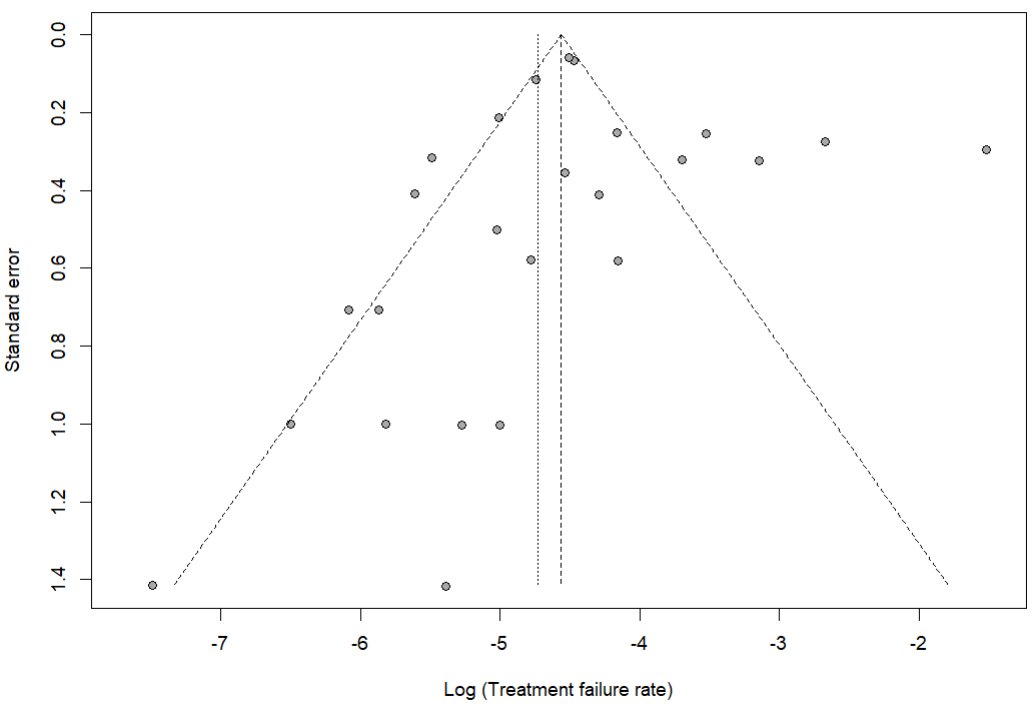

Egger’s test *p*-value = 0.957

Begg’s test *p*-value = 0.378

**Fig. S19** Funnel plot assessing publication bias among studies reporting treatment failure rate among tuberculosis patients in Cameroon, 1998–2022

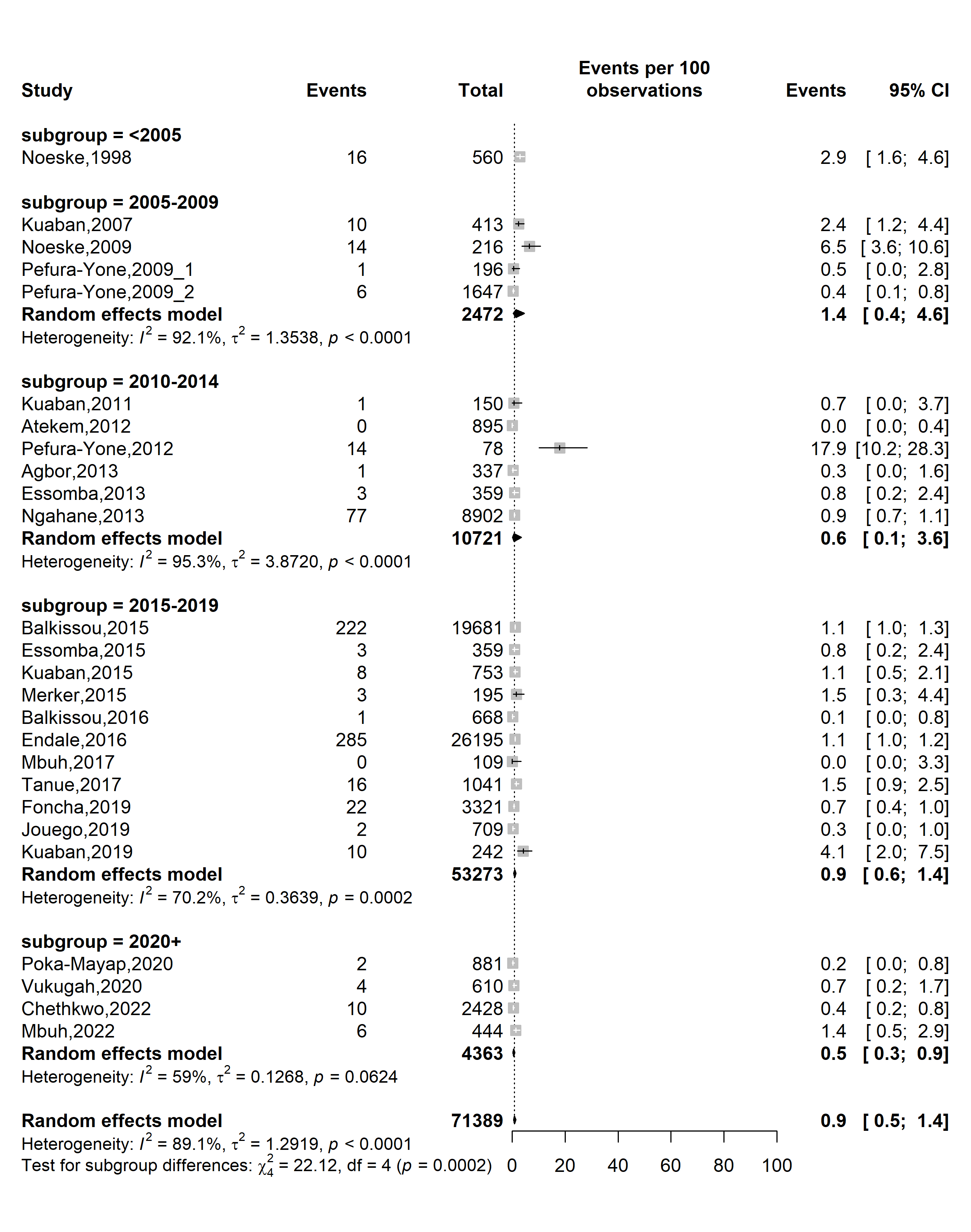

**Fig. S20** Forest plot displaying the subgroup meta-analysis of the pooled treatment failure rate among tuberculosis patients in Cameroon by study timeframe, 1998-2022

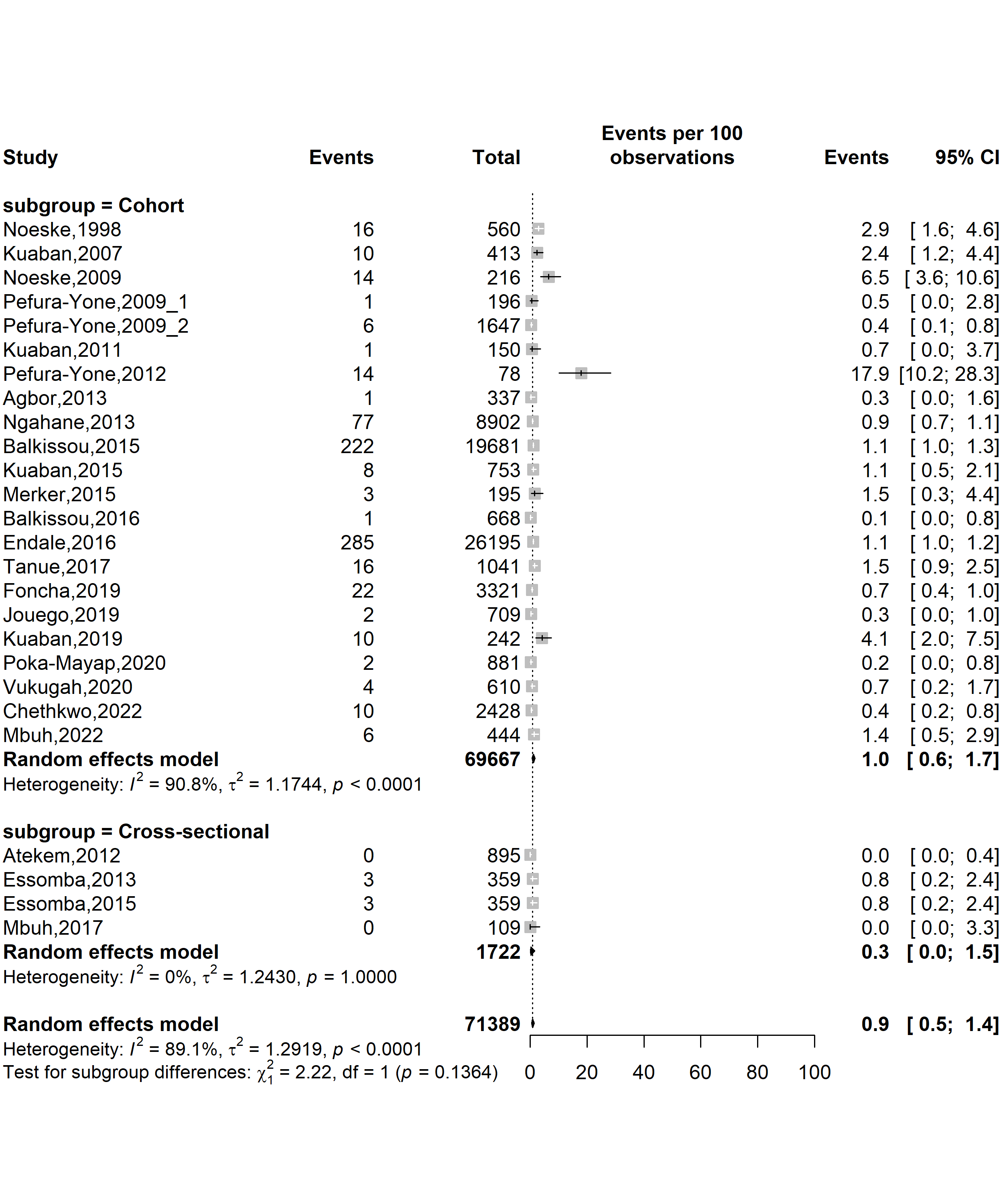

**Fig. S21** Forest plot displaying the subgroup meta-analysis of the pooled treatment failure rate among tuberculosis patients in Cameroon by study designs, 1998-2022

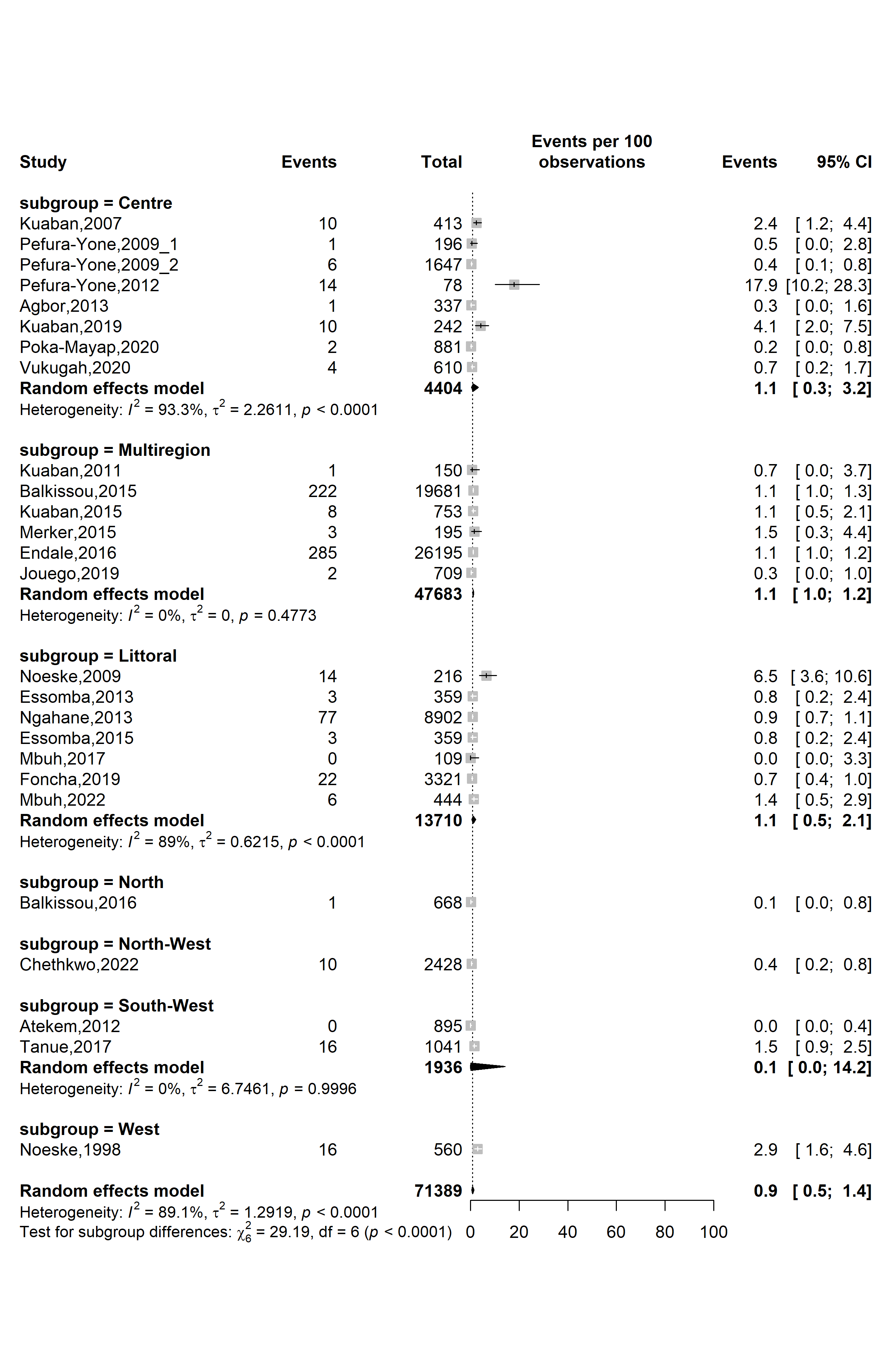

**Fig. S22** Forest plot displaying the subgroup meta-analysis of the pooled treatment failure rate among tuberculosis patients in Cameroon by study regions, 1998-2022

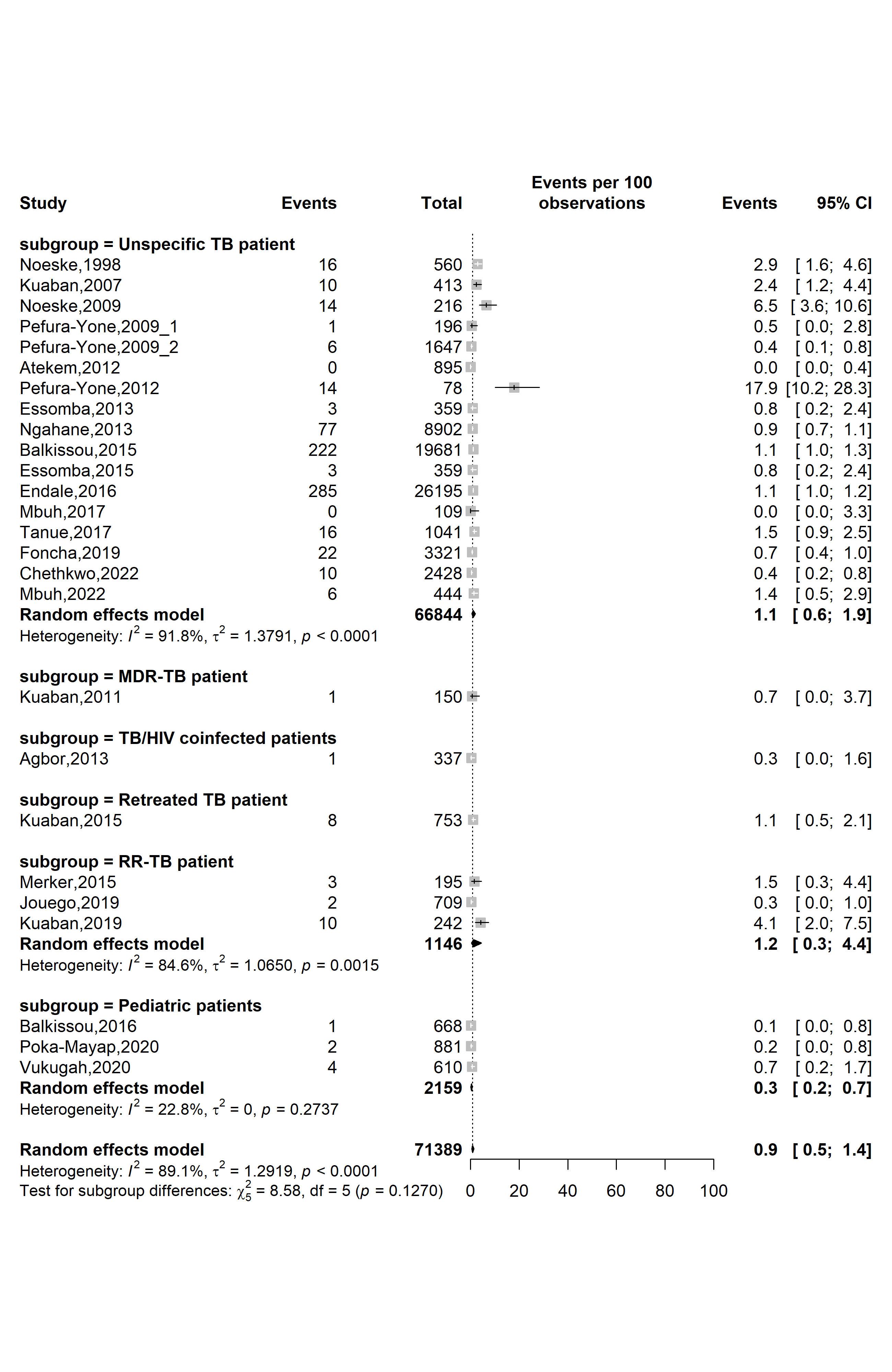

**Fig. S23** Forest plot displaying the subgroup meta-analysis of the pooled treatment failure rate among tuberculosis patients in Cameroon by types of patients, 1998-2022

*(TB: Mycobacterium tuberculosis infection; RR-TB: Rifampicin-resistant tuberculosis; MDR-TB: Multidrug-resistant tuberculosis; HIV: Human immunodeficiency virus)*

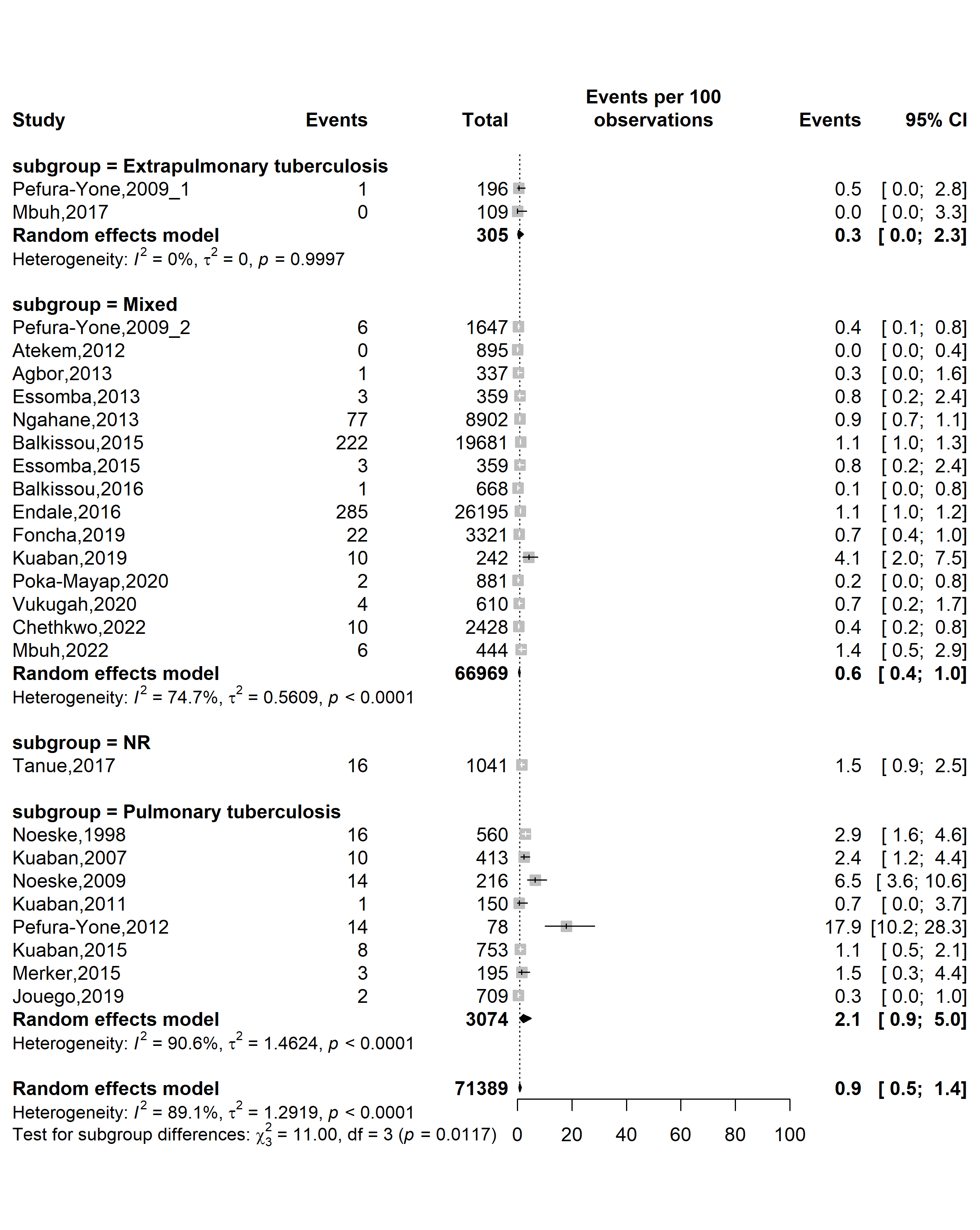

**Fig. S24** Forest plot displaying the subgroup meta-analysis of the pooled treatment failure rate among tuberculosis patients in Cameroon by tuberculosis disease localizations, 1998-2022

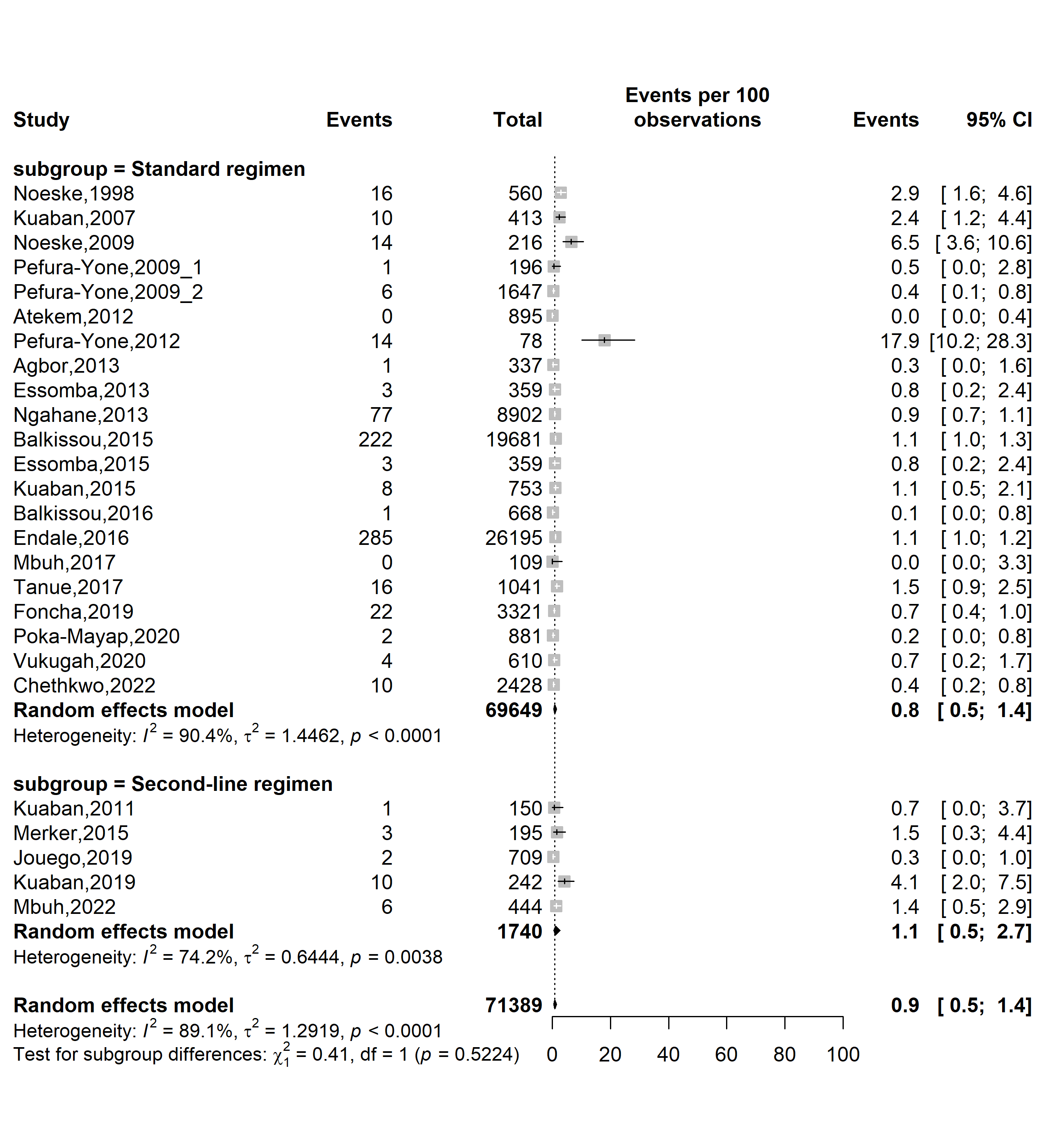

**Fig. S25** Forest plot displaying the subgroup meta-analysis of the pooled treatment failure rate among tuberculosis patients in Cameroon by types of resistant tuberculosis regimens, 1998-2022

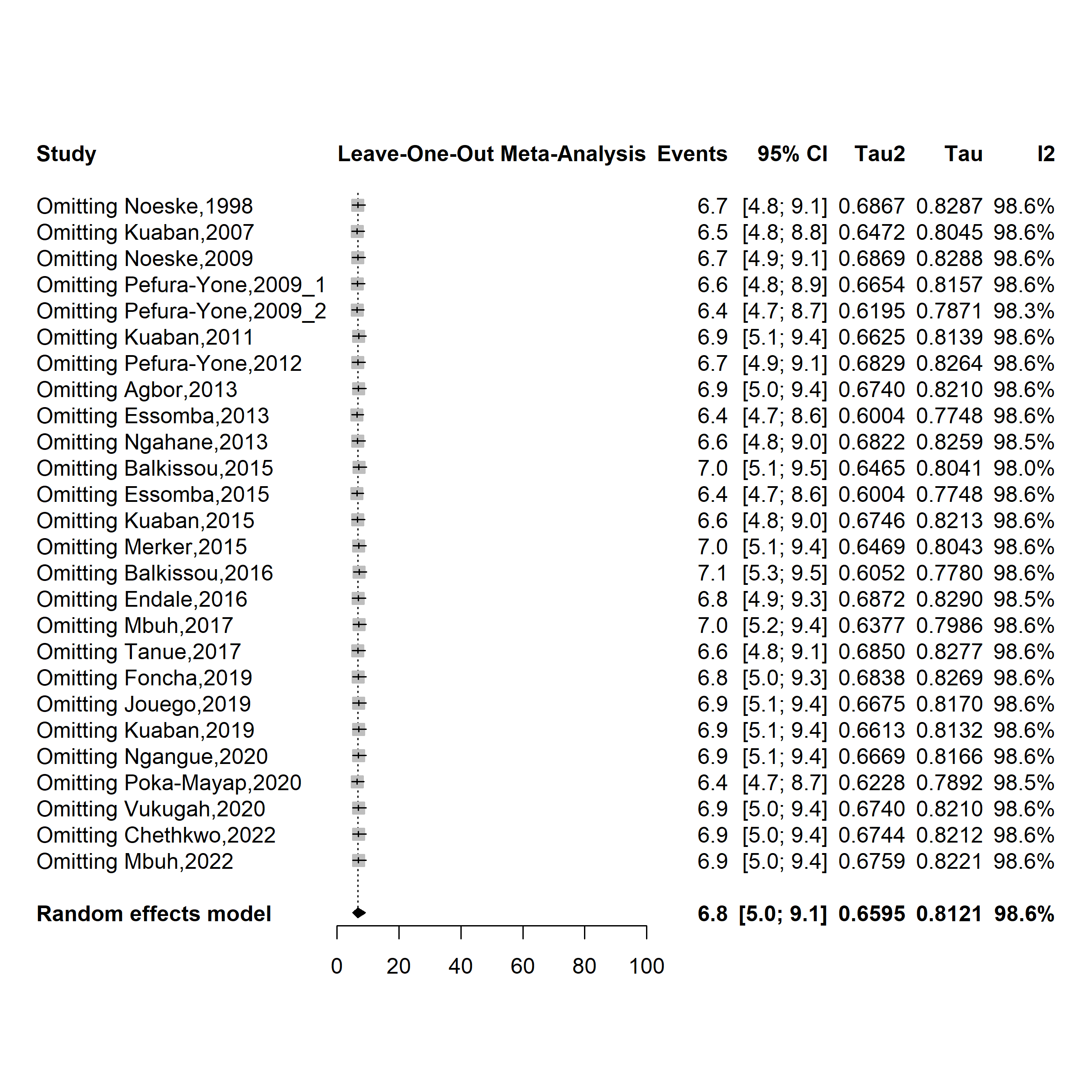

**Fig. S26** Forest plot displaying the sensitivity analysis of the pooled loss to follow-up rate among tuberculosis patients in Cameroon, 1998-2022

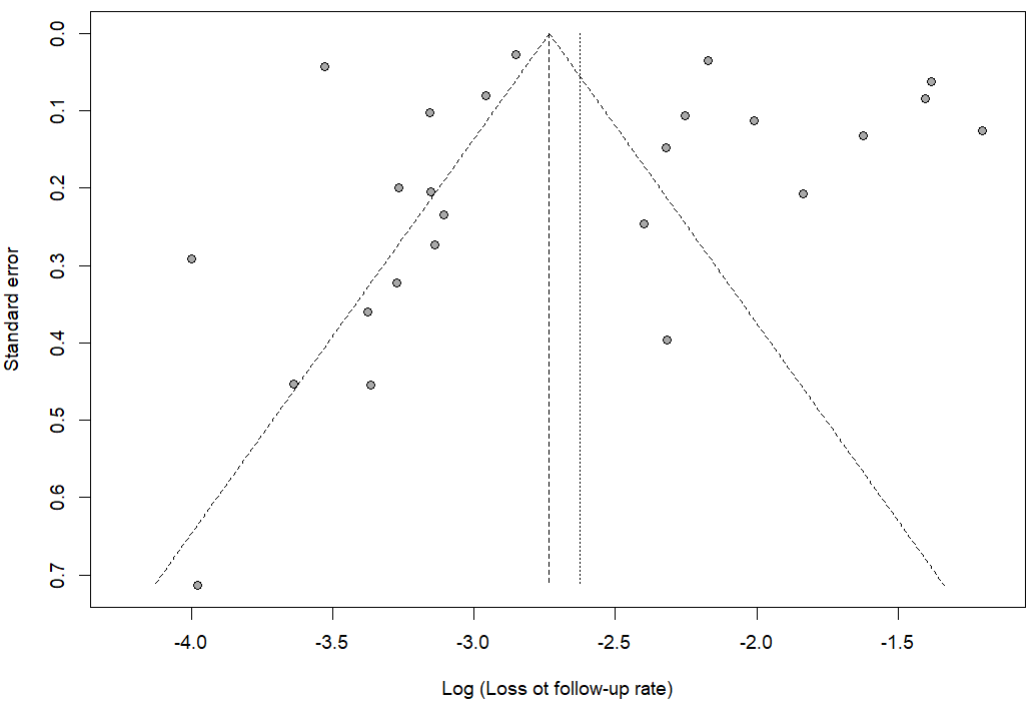

Egger’s test *p*-value = 0.575

Begg’s test *p*-value = 0.378

**Fig. S27** Funnel plot assessing publication bias among studies reporting loss to follow-up rate among tuberculosis patients in Cameroon, 1998–2022

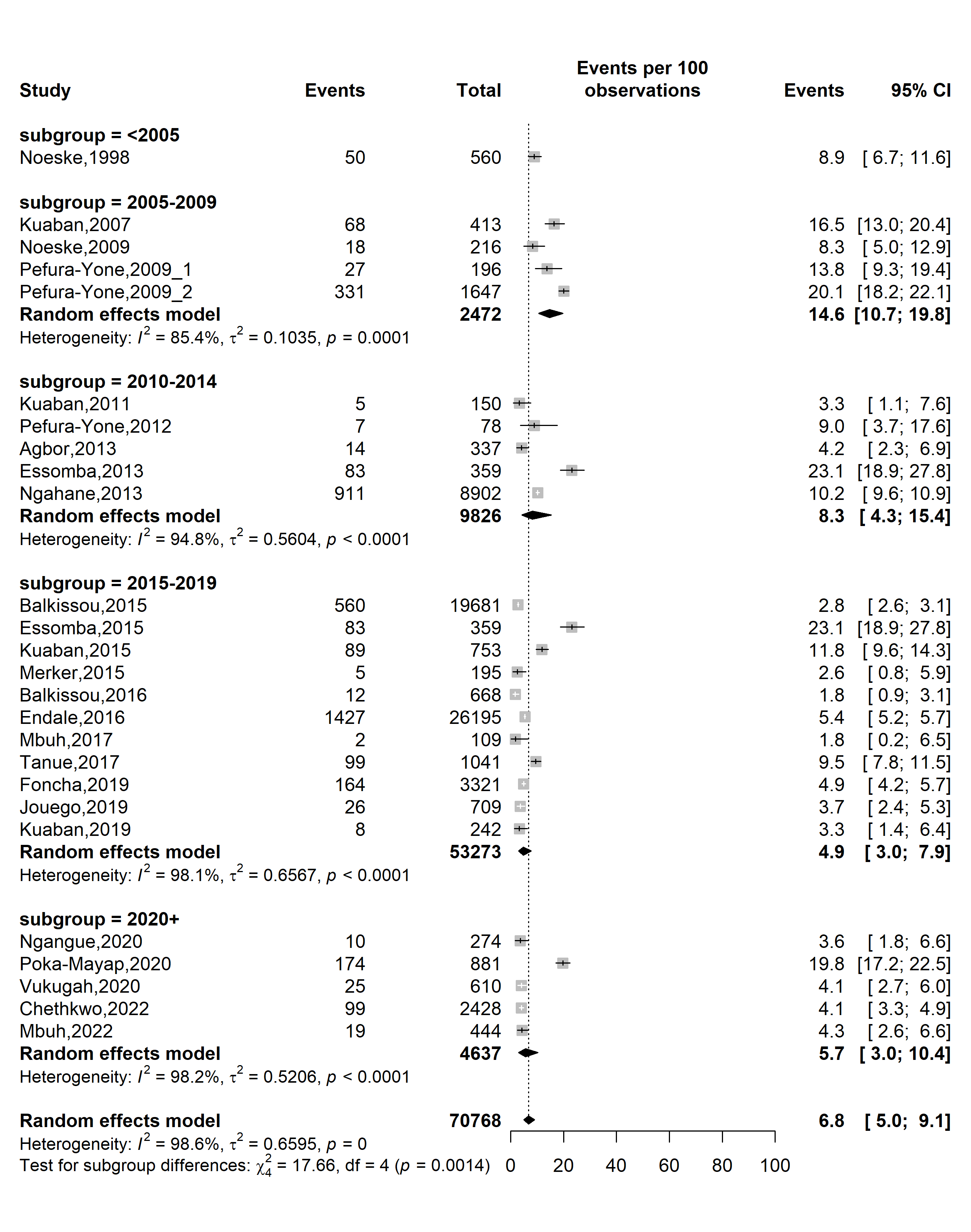

**Fig. S28** Forest plot displaying the subgroup meta-analysis of the pooled loss to follow-up rate among tuberculosis patients in Cameroon by study timeframe, 1998-2022

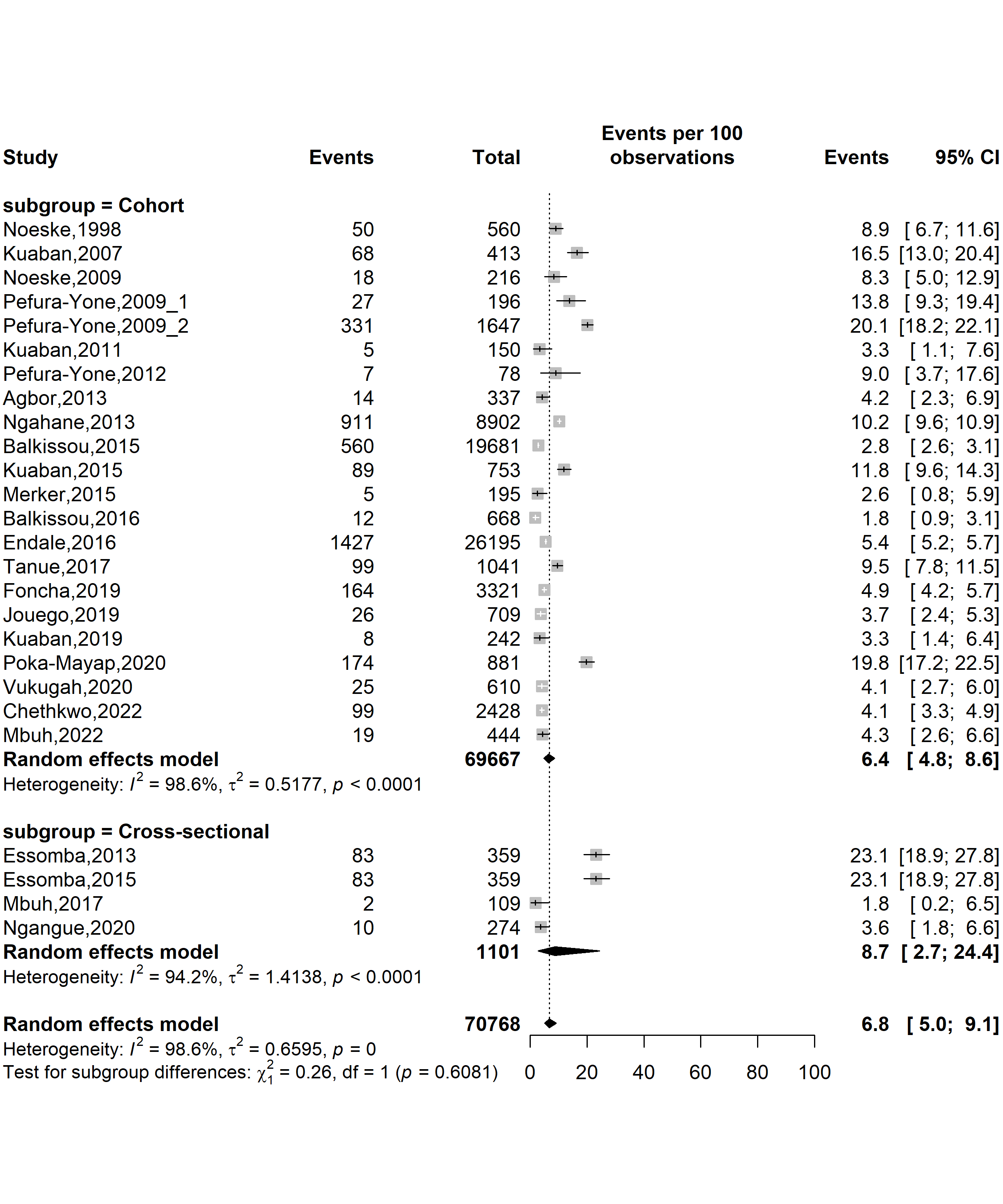

**Fig. S29** Forest plot displaying the subgroup meta-analysis of the pooled loss to follow-up rate among tuberculosis patients in Cameroon by study designs, 1998-2022

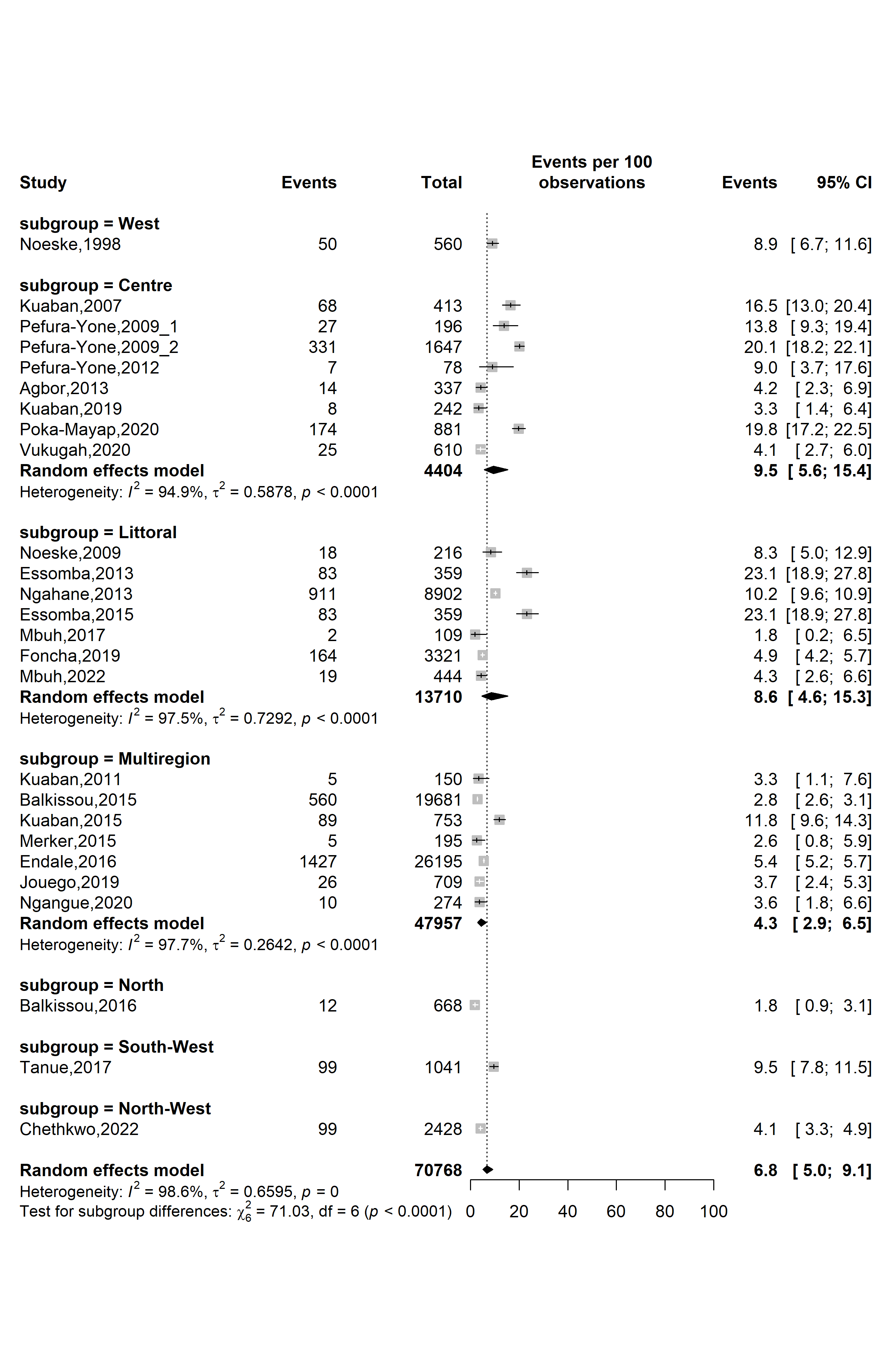

**Fig. S30** Forest plot displaying the subgroup meta-analysis of the pooled loss to follow-up rate among tuberculosis patients in Cameroon by study regions, 1998-2022

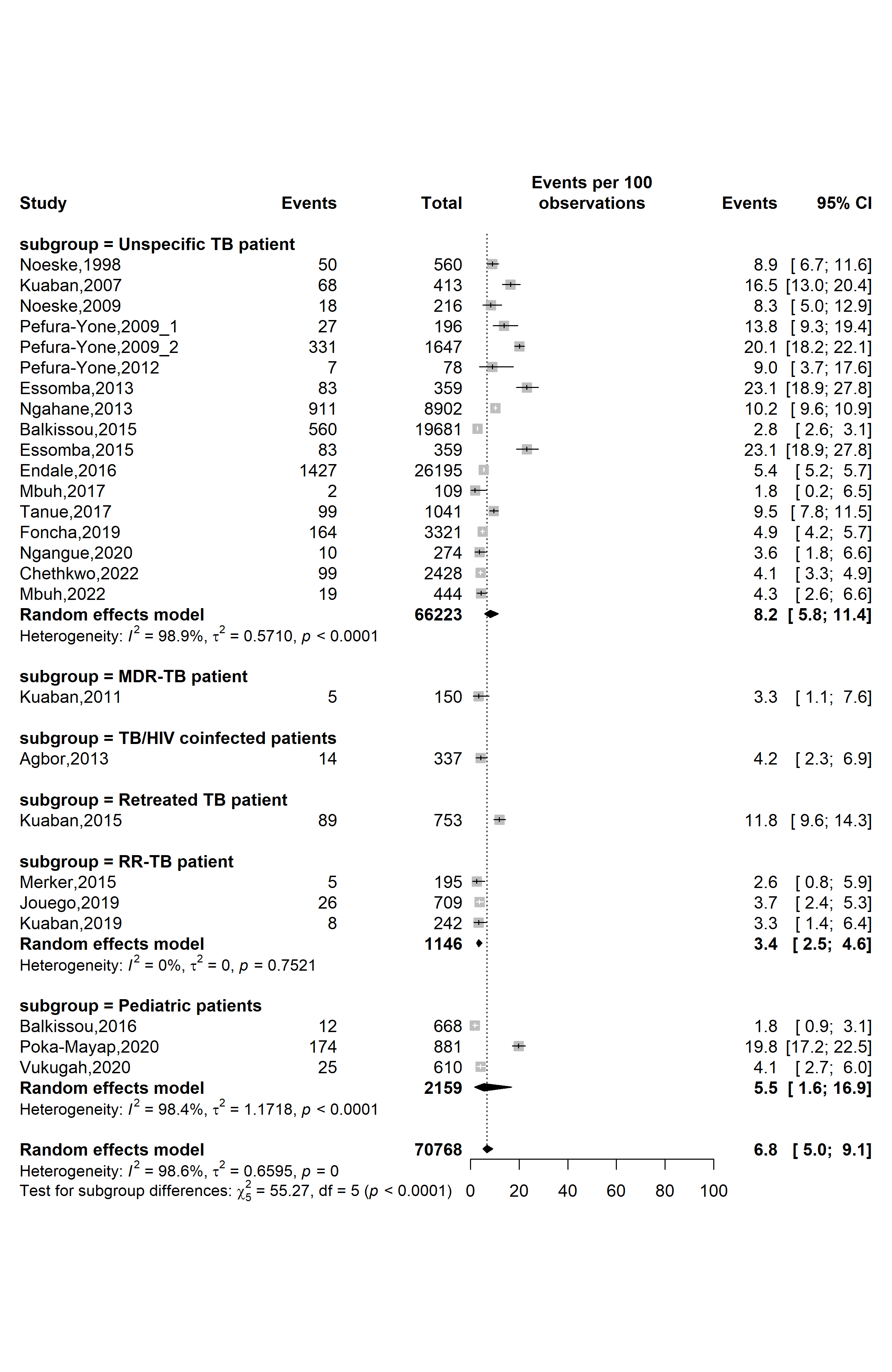

**Fig. S31** Forest plot displaying the subgroup meta-analysis of the pooled loss to follow-up rate among tuberculosis patients in Cameroon by types of patients, 1998-2022

*(TB: Mycobacterium tuberculosis infection; RR-TB: Rifampicin-resistant tuberculosis; MDR-TB: Multidrug-resistant tuberculosis; HIV: Human immunodeficiency virus)*

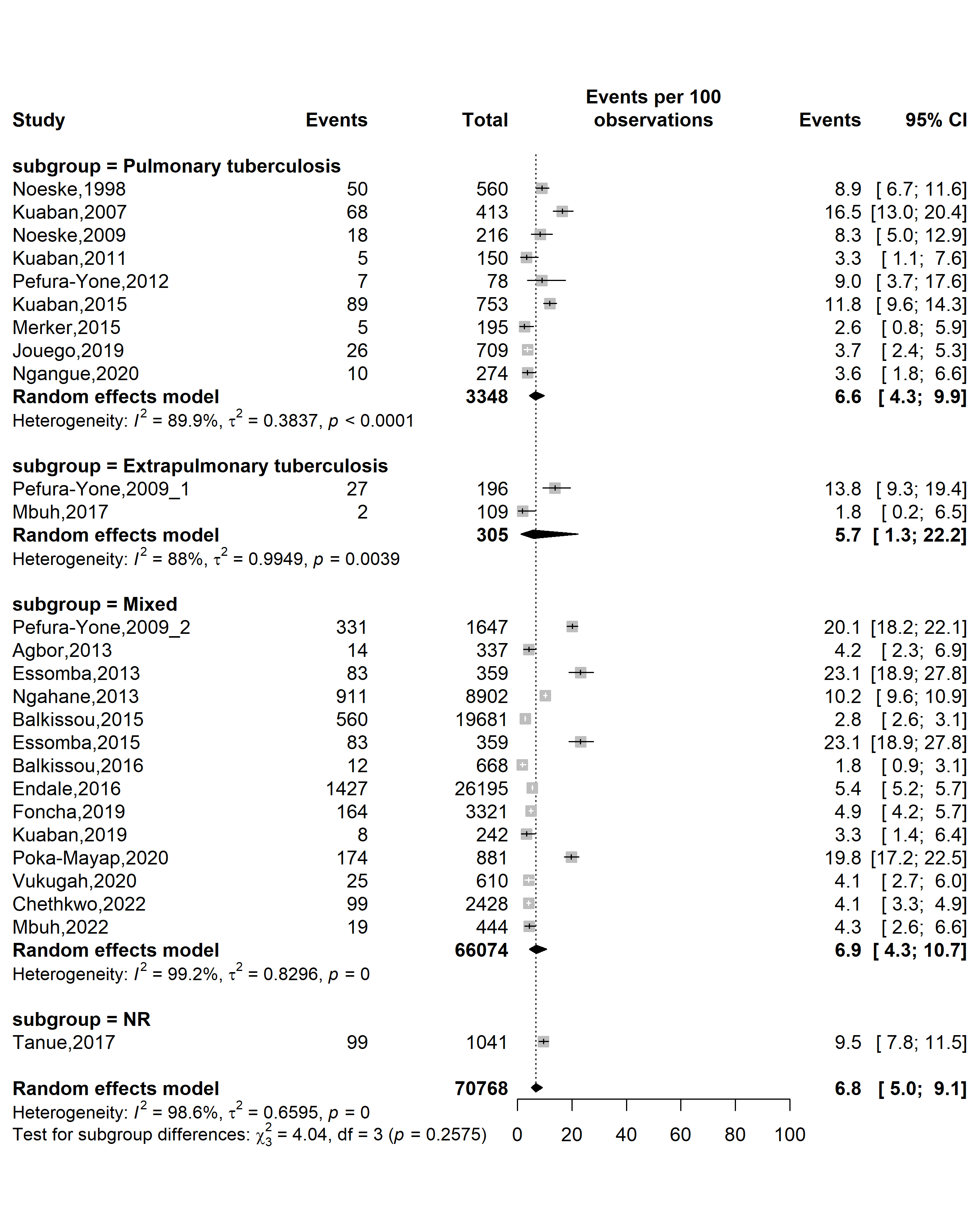

**Fig. S32** Forest plot displaying the subgroup meta-analysis of the pooled loss to follow-up rate among tuberculosis patients in Cameroon by tuberculosis disease localizations, 1998-2022

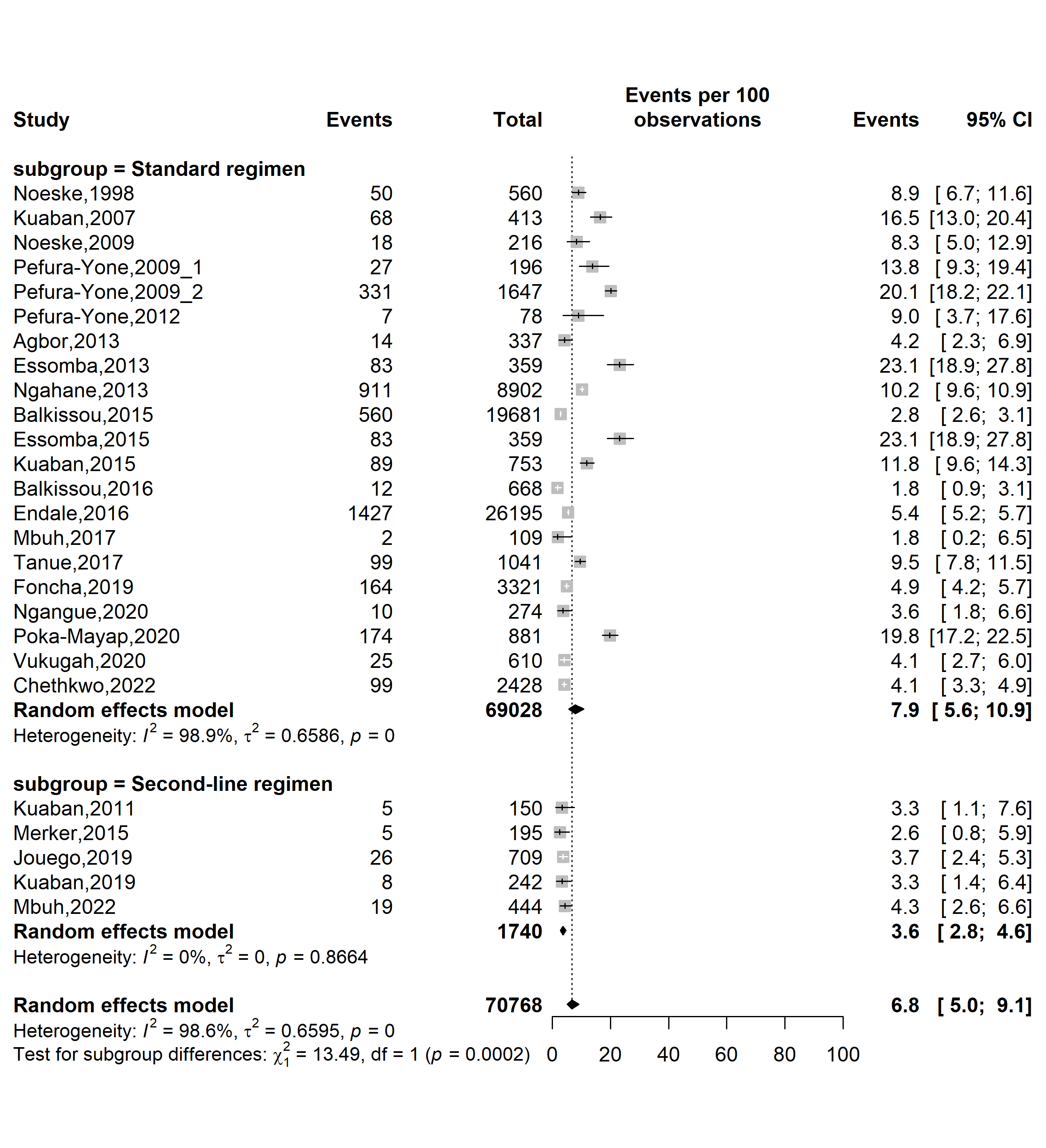

**Fig. S33** Forest plot displaying the subgroup meta-analysis of the pooled loss to follow-up rate among tuberculosis patients in Cameroon by types of resistant tuberculosis regimens, 1998-2022

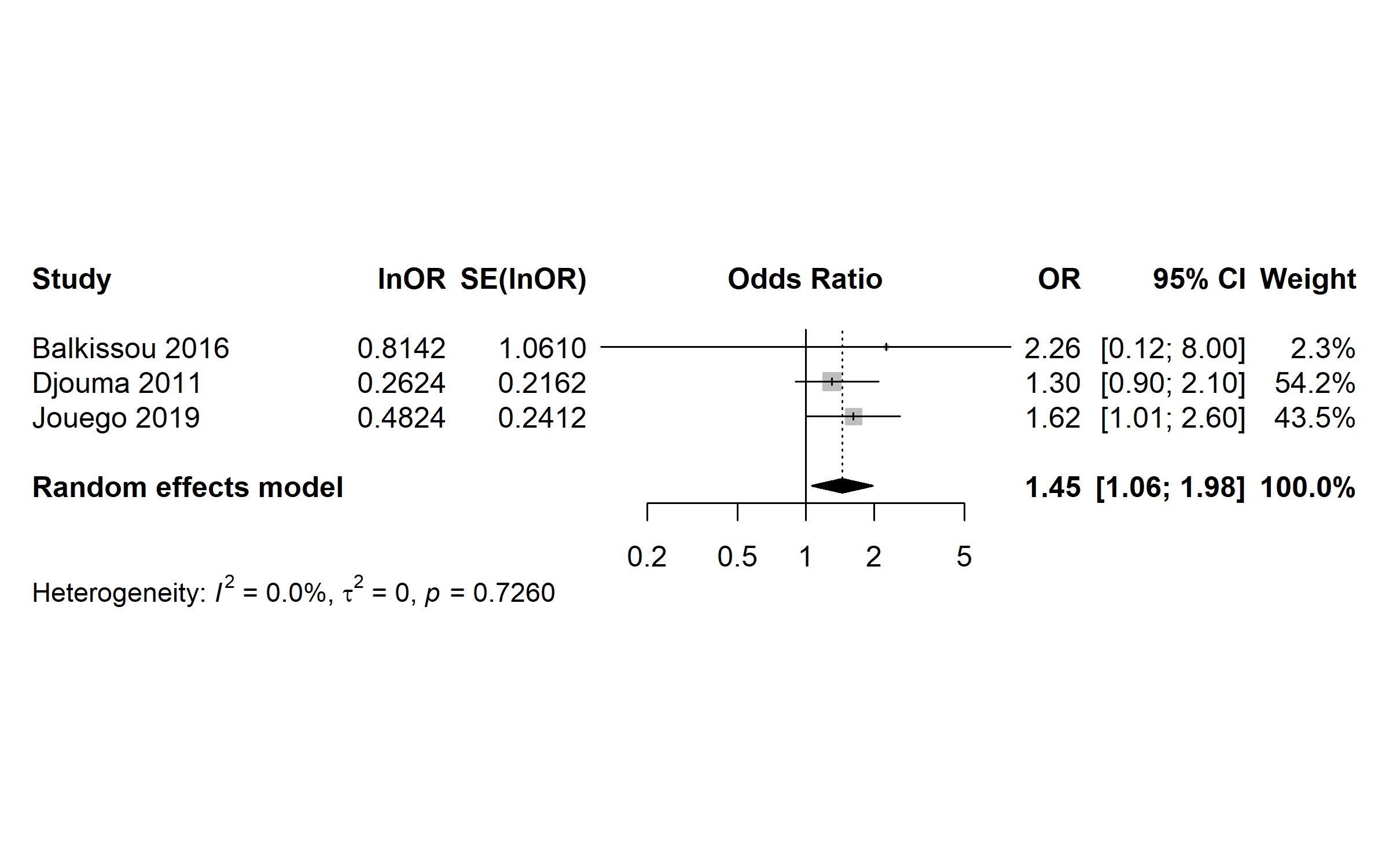

**Fig. S34** Pooled odds ratio of unfavorable outcome among patients treated for tuberculosis in Cameroon, 1998-2022 (Age: ≤ 40 years vs. ˃ 40 years)

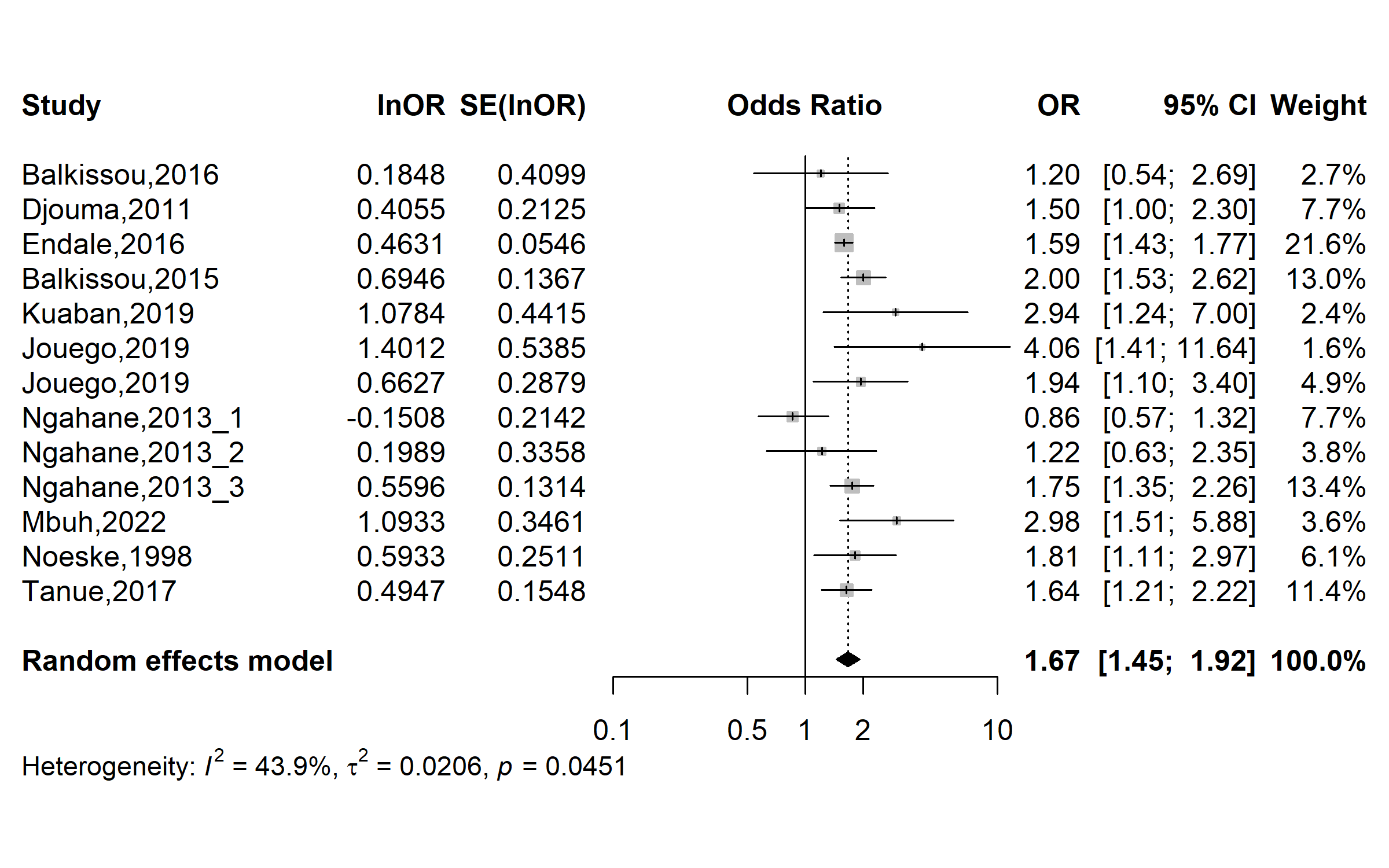

**Fig. S35** Pooled odds ratio of unfavorable outcome among patients treated for tuberculosis in Cameroon, 1998-2022 (Sex: Male vs. Female)

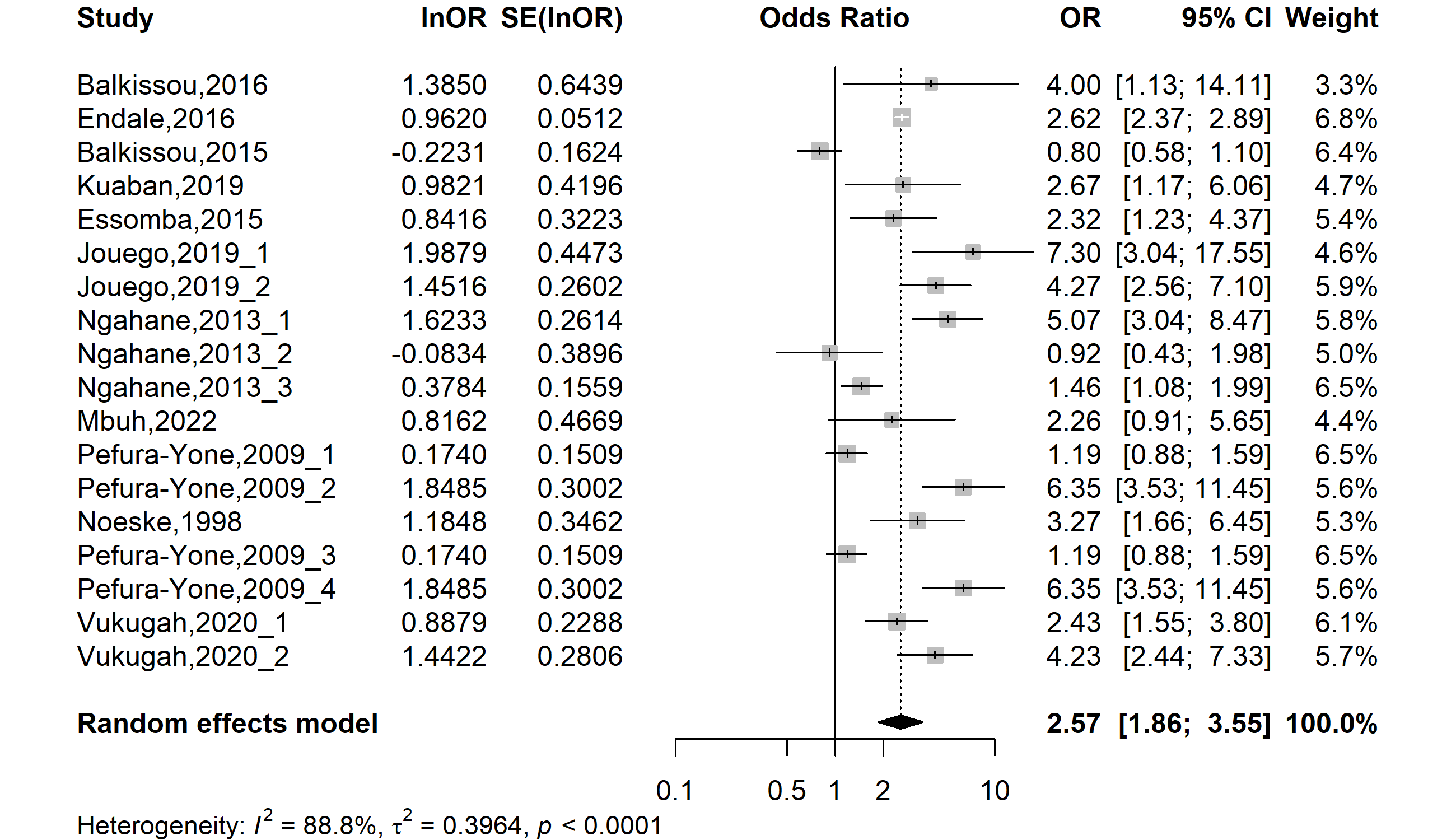

**Fig. S36** Pooled adjusted odds ratio of unfavorable outcome among patients treated for tuberculosis in Cameroon, 1998-2022 (Human immunodeficiency virus serological status: Positive vs. Negative)

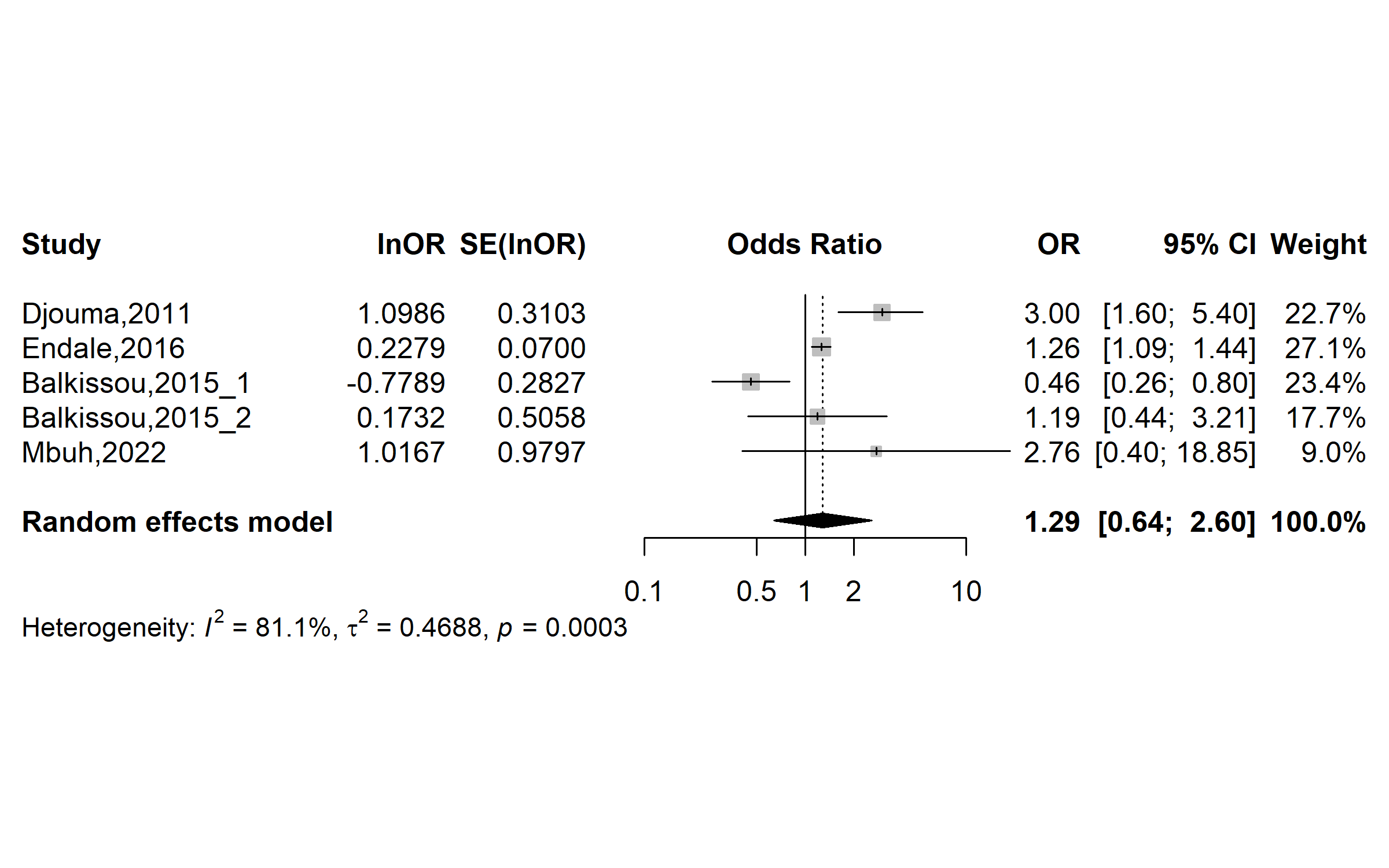

**Fig. S37** Pooled odds ratio of unfavorable outcome among patients treated for tuberculosis in Cameroon, 1998-2022 (Tuberculosis disease localization: Extrapulmonary vs. Pulmonary)

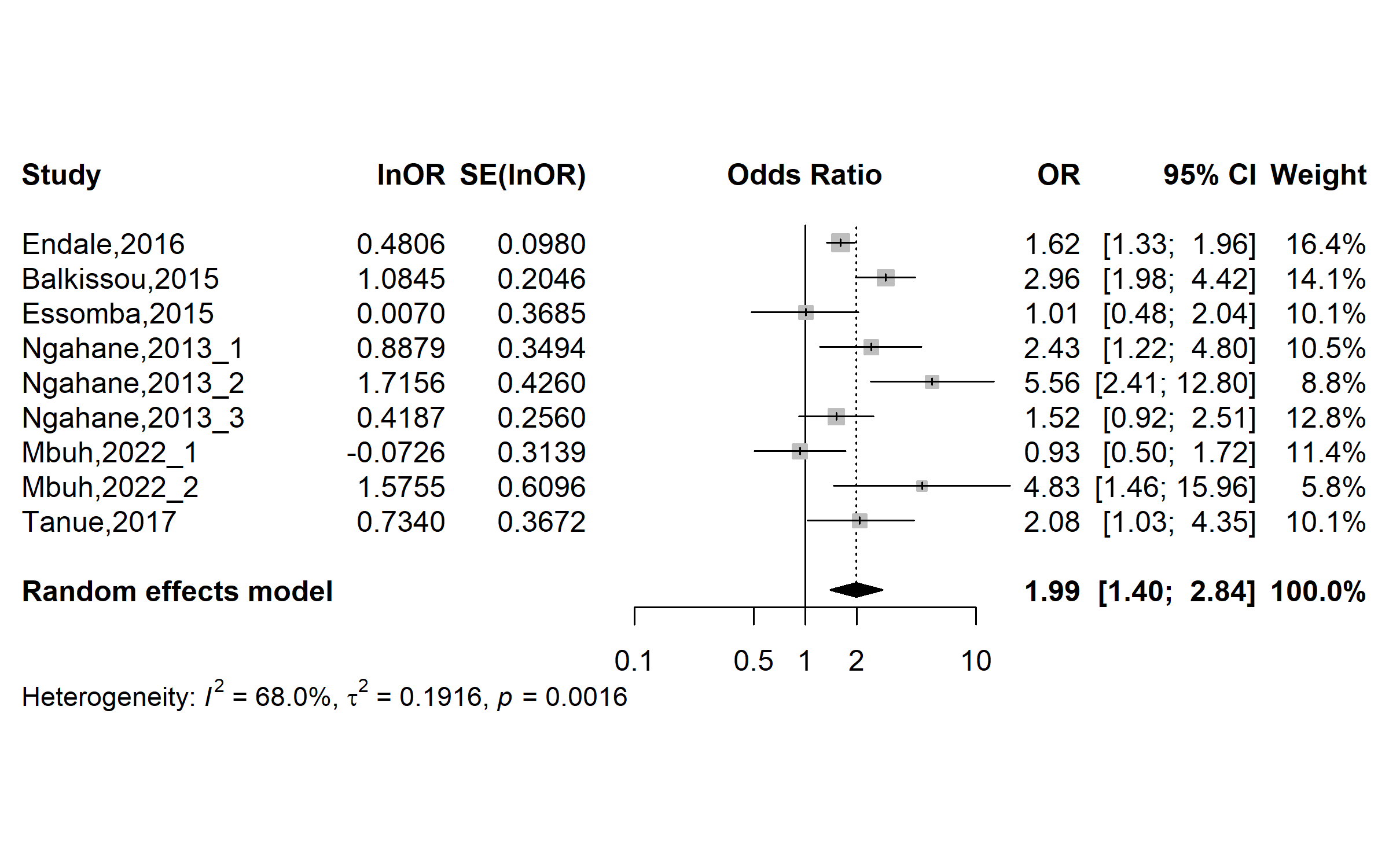

**Fig. S38** Pooled odds ratio of unfavorable outcome among patients treated for tuberculosis in Cameroon, 1998-2022 (Previously treated tuberculosis disease: Yes vs. No)

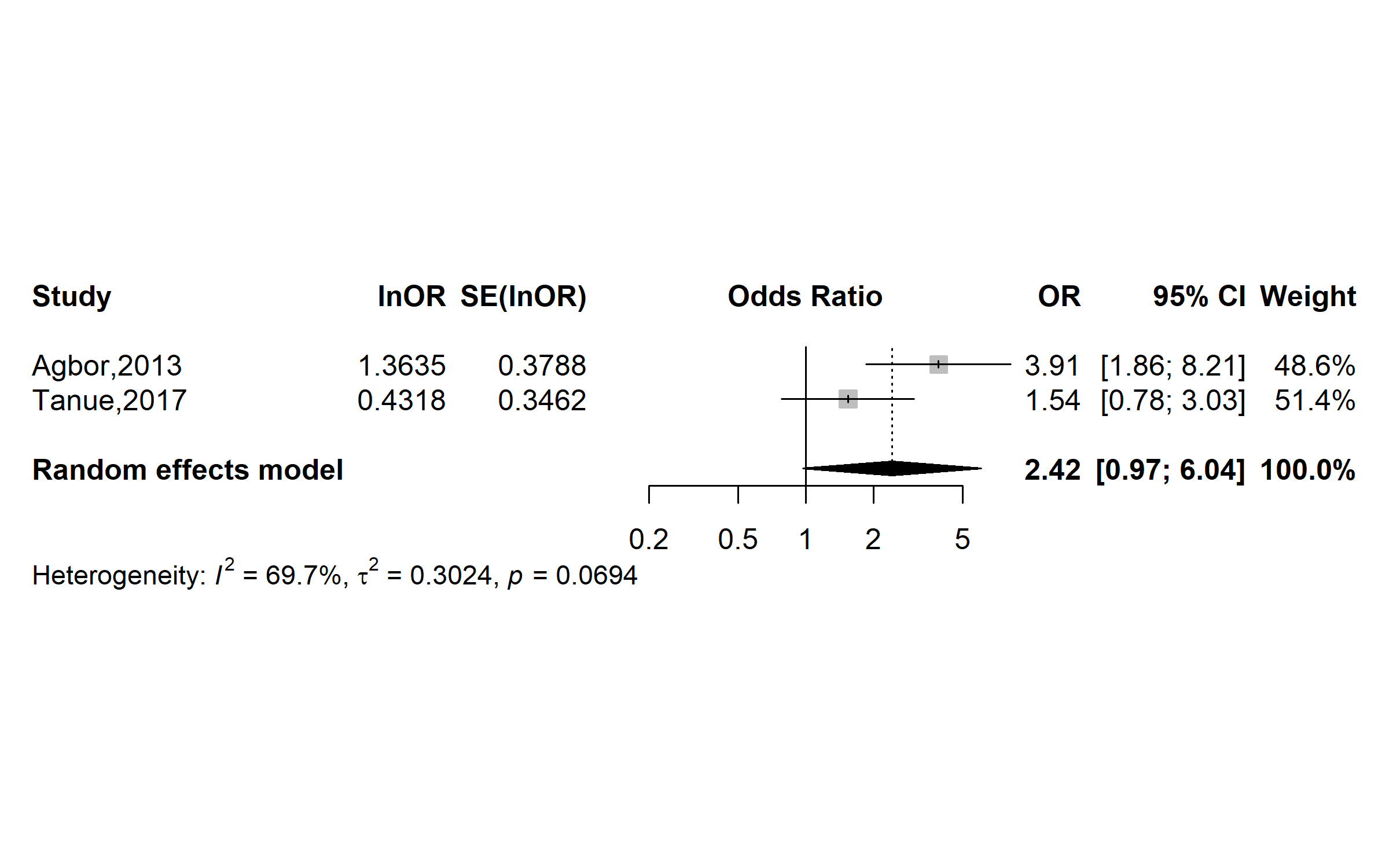

**Fig. S39** Pooled odds ratio of unfavorable outcome among patients treated for tuberculosis in Cameroon, 1998-2022 (Under co-trimoxazole chemoprophylaxis: No vs. Yes)

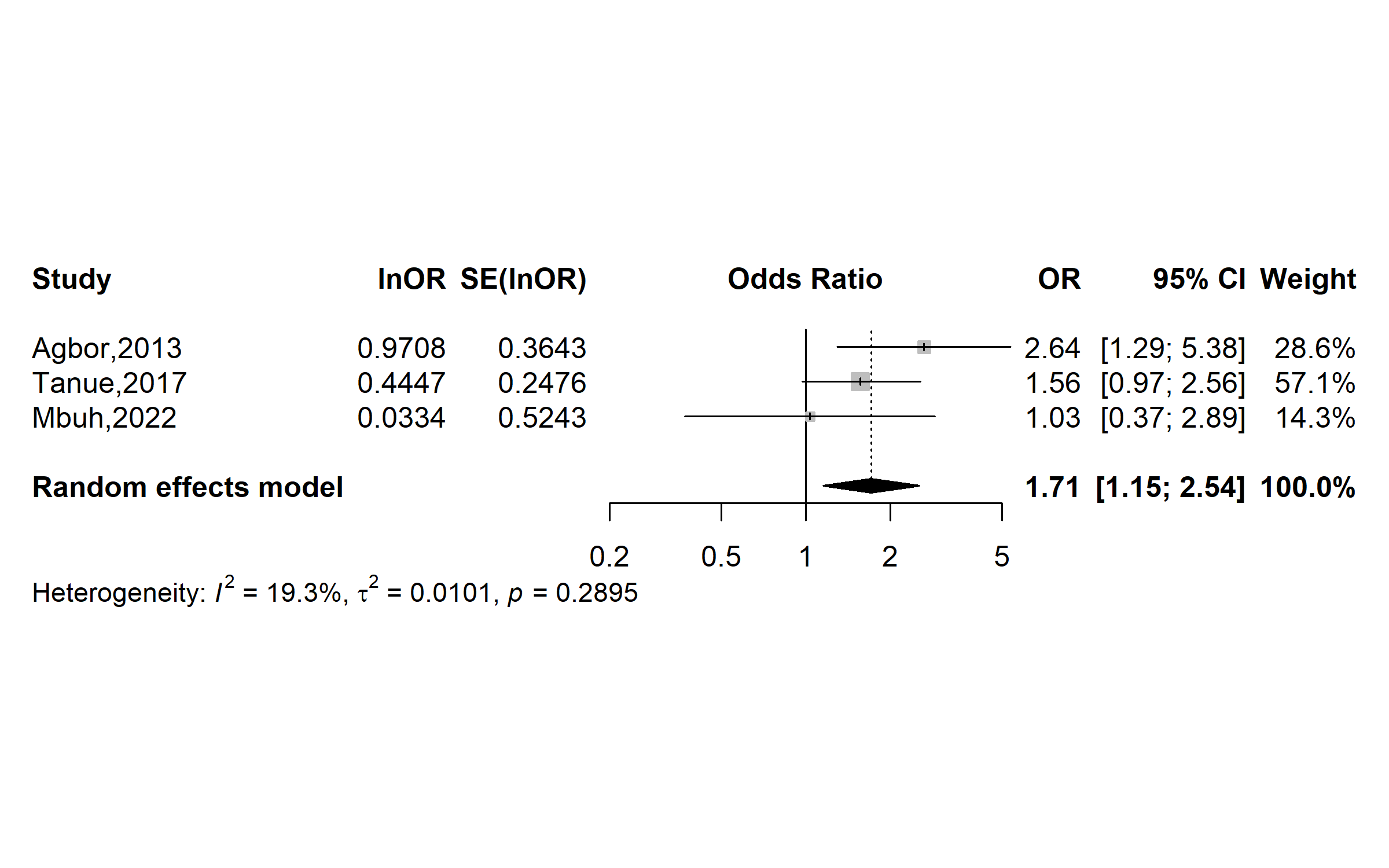

**Fig. S40** Pooled odds ratio of unfavorable outcome among patients treated for tuberculosis in Cameroon, 1998-2022 (Under antiretroviral therapy: No vs. Yes)
